# Plasma multi-omics analysis reveals very long chain ceramides as validated biomarkers of Friedreich’s ataxia

**DOI:** 10.1101/2022.09.27.22280432

**Authors:** Dezhen Wang, M. Grazia Cotticelli, Blanca E. Himes, David R. Lynch, Clementina Mesaros

## Abstract

Friedreich’s Ataxia (FRDA) is an autosomal neurodegenerative disease caused by the deficiency of the protein frataxin. Frataxin is a critical enzyme in the assembly of iron-sulfur clusters that are cofactors for several metabolic enzymes. To identify metabolic features that could be used as potential biomarkers for FRDA in plasma, we performed a multi-omics analysis using a discovery-validation cohort design. We combined metabolomics, lipidomics and proteomics from several liquid chromatography-high resolution mass spectrometry platforms. The analyses revealed that FRDA patients compared to healthy controls and unaffected carriers had dysregulated sphingolipids metabolism, phospholipid metabolism, citric acid cycle, amino acid metabolism, and apolipoprotein metabolism. Using an ROC, the decreased very long chain ceramides can distinguished FRDA patients from healthy controls with AUC from 0.75 to 0.85. Using induced pluripotent stem cell differentiated cardiomyocytes (iPSC-CMs), we demonstrated that frataxin deficiency preferentially affected ceramide synthase (CerS2), enriching long chain ceramides, and depleting very long chain ceramides. The ceramide metabolism was differentially regulated in two of the affected tissues in FRDA: heart and muscles. A machine-learning model improved the prediction of FRDA using the combination of three plasma metabolites (AUC > 0.9). In conclusion, decreased very long chain ceramides are reliable plasma biomarkers for FRDA patients.

**One Sentence Summary:** New plasma lipids biomarkers of Friedreich’s Ataxia (FRDA) were validated using a discovery-validation design with two independent cohorts.

## INTRODUCTION

Friedreich’s Ataxia (FRDA) is an autosomal recessive neurodegenerative disease characterized by progressive mobility loss, impairment of speech, ^1^ and eventual loss of vision ^2^ and hearing^3^. Individuals with FRDA exhibit clinical manifestations of ataxia, hypertrophic cardiomyopathy, scoliosis, and diabetes mellitus ^4–7^. Cardiac complications are the major cause of early death in FRDA ^8^. Most disease alleles in FRDA have a guanine-adenine-adenine (GAA) repeat expansion in the first intron of the disease gene *FXN*, this repeat leads to the silencing of the *FXN* gene and decreased frataxin protein levels ^9, 10^. Frataxin acts in the assembly of iron-sulfur (Fe-S) clusters, which are important for iron homeostasis and optimal functions of metabolic enzymes for citric acid cycle, electron chain transport, and fatty acid breakdown ^11–14^. Defective mitochondrial metabolism and mitochondrial functions are observed in different models of FRDA ^15–17^ including Fe–S containing aconitase and respiratory chain complexes ^18^ that result in a failure of energy (ATP) production ^19, 20^. Frataxin-deficient cardiomyocytes present altered thiol-redox state and a decrease content in pyruvate dehydrogenase (PDH A1) ^21^ and decreased pyruvate oxidation and increased fatty acid oxidation was observed in platelets from FRDA patients ^22^. Frataxin deficiency influences the peroxisome-proliferator activator receptor gamma (PPARγ)/PPARγ coactivator 1 alpha (Pgc1a) pathway in cellular and animal models, which is linked to mitochondrial biogenesis and lipid metabolism ^23^. Frataxin reduction induces iron accumulation and enhances the sphingolipid/PDK1/Mef2 pathway, which triggers neurodegeneration in fly and mouse models ^24, 25^. As discussed in a 2019 summary of FRDA biomarkers ^26^, metabolic dysfunctions are significant features of FRDA patients, but larger population studies are necessary to discover and validate actionable biomarkers. A recent study ^27^ uncovered dysregulation in the one carbon metabolism in plasma from FRDA patients. A common feature of all previous human studies is the limited number of FRDA patients and the lack of validation of the new finding in an independent cohort.

To understand the pathophysiology of FRDA, a high-throughput and large-scaled screening strategy could be employed to investigate the metabolic dysfunctions of FRDA and discover new biomarkers. Metabolomics and proteomics-based profiling provide a useful tool to unravel such biomarkers. Omics profiling have been widely used for biomarker discoveries, drug response prediction, and pathogenesis-related pathway revelation for other diseases ^28–30^. Recently, ceramide levels were shown to be accurate biomarkers of adverse cardiovascular disease outcomes in human serum ^31^.

To identify potential blood biomarkers in FRDA, we performed metabolomics, lipidomics, and proteomics analysis using discovery-validation design in two independent cohorts of fasting plasma samples. Our results showed that very long chain ceramides distinguish FRDA patients from healthy controls. We further investigated the ceramide metabolism in human induced pluripotent stem cell derived cardiomyocytes (iPSC-CMs) using high-resolution mass spectrometry (HRMS) lipidomics and isotope tracing assays, and consistently found that frataxin deficiency disrupted ceramide synthesis and preferentially enriched long chain ceramides but depleted very long chain ceramides. Using several cell lines from different affected tissues in FRDA patients, we demonstrated that ceramide metabolism was tissue specific differentially regulated. Our study provides new validated plasma biomarkers for FRDA patients, and these biomarkers could be potential therapeutic targets.

## RESULTS

### Experimental design and targeted multi-omics workflow

To identify metabolic features that can be used for potential biomarkers in FRDA plasma, we developed independent targeted metabolomics, lipidomics and proteomics workflows using ultra-high performance liquid chromatography-high resolution mass spectrometry (UHPLC-HRMS), and measured hundreds of metabolites, lipids, and proteins (Fig. 1A).

**Fig. 1.**
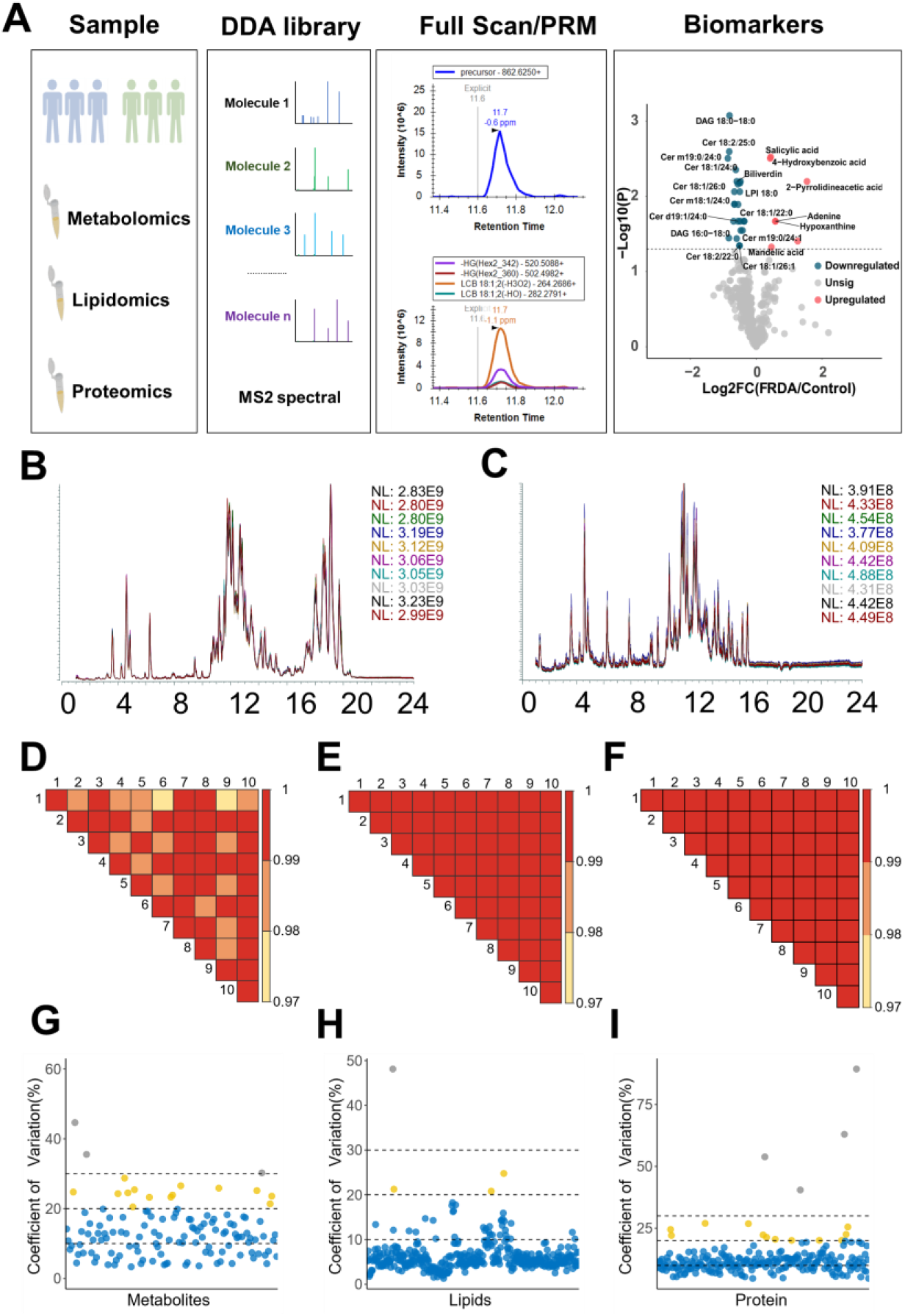
A multi-omics workflow to identify potential plasma biomarkers of FRDA. **A.** Multi-omics analysis workflow. Plasma samples from healthy controls and FRDA patients were pooled and processed in three different ways for metabolomics, lipidomics, or proteomics. Libraries were built by running the pooled plasma samples and authentic standards in a data-dependent acquisition (DDA) mode. Narrow scheduled chromatographic windows (1-2 min) were set based on DDA libraries to acquire MS2 data in a parrel reaction monitoring (PRM) mode, and full scan (MS1) data was also acquired simultaneously to have a better annotation. Univariate and multivariate statistical analysis were performed to identify potential biomarkers of FRDA. **B.** The overlapped total ion counts (TICs) from 10 technical workflow replicates for lipidomics in positive mode. **C.** Same as B but in negative mode. Those showed excellent chromatographic reproducibly. **D.** Color-coded R^2^ values for the Pearson correlation comparison of 10 technical workflow replicates for metabolomics. **E.** lipidomics **F.** proteomics. R^2^ values indicated high reproducibility. **G.** Intra-assay variability was evaluated by calculating the coefficient of variation (CV) for 10 technical workflow replicates for all quantified metabolites. **H.** lipids **I.** peptides; CV < 20% colored with blue, 20% < CV <30% colored with yellow, and CV > 30% colored with grey.

Using pooled plasma samples (Supplementary Table 1) or standard solutions, we build-up reliable MS^2^ spectral libraries by analyzing pooled samples in a data-dependent acquisition (DDA) mode. We included more than 50 internal standards (^13^C, ^2^H, or chain length analogs) (Supplementary Table 2) in combination with quality controls (QC) for locally estimated scatterplot smoothing (LOESS) to normalize for signal intensity drift. Next, well annotated (see experimental part), metabolites, lipids, and peptides were used to generate scheduled window-based inclusion lists for each individual omics experiment (Supplementary Table 3-5) for targeted analysis (Supplementary Fig. 1-3).

To validate the reproducibility and the robustness of our workflows, we separately processed and analyzed 10 technical replicates formed from pooled plasma samples for each of the workflow for metabolomics, lipidomics, and proteomics. We analyzed all the samples in full scan (FS) and parallel reaction monitoring (PRM) modes providing untargeted profiling in combination with more specific targeted quantification without significantly compromising the data acquisition speed. Excellent chromatographic reproducibility was observed for the positive (Fig. 1B) and negative (Fig. 1C) modes for the lipidomics workflow. For all the analyses, we combined the positive and negative mode data. Metabolomics analysis included 123 metabolites and Pearson correlation showed high coefficients (R^2^, 0.97-1) between 10 technical replicates for metabolomics (Fig. 1D). For lipidomics (Fig. 1E) and proteomics (Fig. 1F), even higher correlation coefficients (R^2^>0.99) were observed for the 10 technical replicates, indicating the high reproducibility of the workflow for these assays. To test the reproducibility of all the tested features, we calculated the coefficient of variance (CVs) for all quantified metabolites, lipids, and peptides. For metabolomics, three metabolites had CVs higher than 30%, and 14 metabolites had CVs higher than 20% (Fig. 1G). For lipidomics, out of 630 lipids, only four metabolites showed CVs higher than 20% (Fig. 1H). For proteomics, four peptides had CVs higher than 30%, and 12 peptides had CVs higher than 20% (Fig. 1I). All these preliminary technical data demonstrated that our approach using a targeted multi-omics workflow is a robust method for biomarker identification.

### Metabolomics and lipidomics analysis revealed dysregulated metabolism in FRDA

Using this targeted multi-omics workflow, we performed both metabolomics and lipidomics analysis using the discovery set of EDTA fasting plasma samples that included 15 controls and 33 FRDA patients (Supplementary Table 1). Those samples were collected between 2012-2019 as part of a Natural History study at the Children’s Hospital of Pennsylvania. The average age of the FRDA patients was 25.6±8.1 and the age of the controls was not significantly different 32.7±11.0. This discovery set also included 11 unaffected carriers, with a significantly different age, as the carriers are usually parents or relatives of the FRDA patients. All groups were balanced on sex as shown in Supplementary Table 1. We visualized the data distribution globally by principal component analysis (PCA). A clear but partial separation of the FRDA group (light blue) from the Control/Carrier groups was observed on the PC1 (Fig. 2A), indicating the metabolic features correlated with disease condition. Most small molecule analytes showed small CVs in our QCs (Supplementary Fig. 4). To identify significantly changed metabolites and lipids, we performed pairwise comparison using a *t*-test. There were no significantly different metabolites between the Control and Carrier groups in the volcano plot from Fig. 2B, which suggests that the Carriers are not metabolically different compared with controls. However, more than 30 metabolites or lipids had a false discovery rate (FDR) <0.05 and were dysregulated in the FRDA patients in comparison with healthy controls (Fig. 2C). We next applied multivariate statistical analysis method (least square discrimination analysis (PLS-DA)) to calculate the contribution of metabolites or lipids to model discrimination between the Control and FRDA groups (variable importance in projection-VIP). We found that all the metabolites having significant P values also showed higher VIP values (Fig. 2D). To visualize the distribution of the top 50 metabolites or lipids (FDR<0.1), we plotted these as a heatmap for carriers, controls, and FRDA patients (Fig. 2E). To detect the functional groups of metabolites that changed in FRDA patients, we performed correlation analysis on the amount of top 50 compounds (FDR < 0.1). The compounds can be categorized into several groups based on their structure and biological data. On the basis of partial correlation relationships, we build the metabolite correlation network (Fig. 2F and Supplementary Fig. 5) that showed three major clusters. The largest cluster was composed of multiple ceramides, and high intra-correlations were observed for these ceramides. The second largest cluster was the phospholipid cluster, composed of phosphocholines (PC), lyso-PC (LPC), phosphatidic acids (PA), phosphoinositols (PI), and lyso-PI (LPI).

**Fig. 2.**
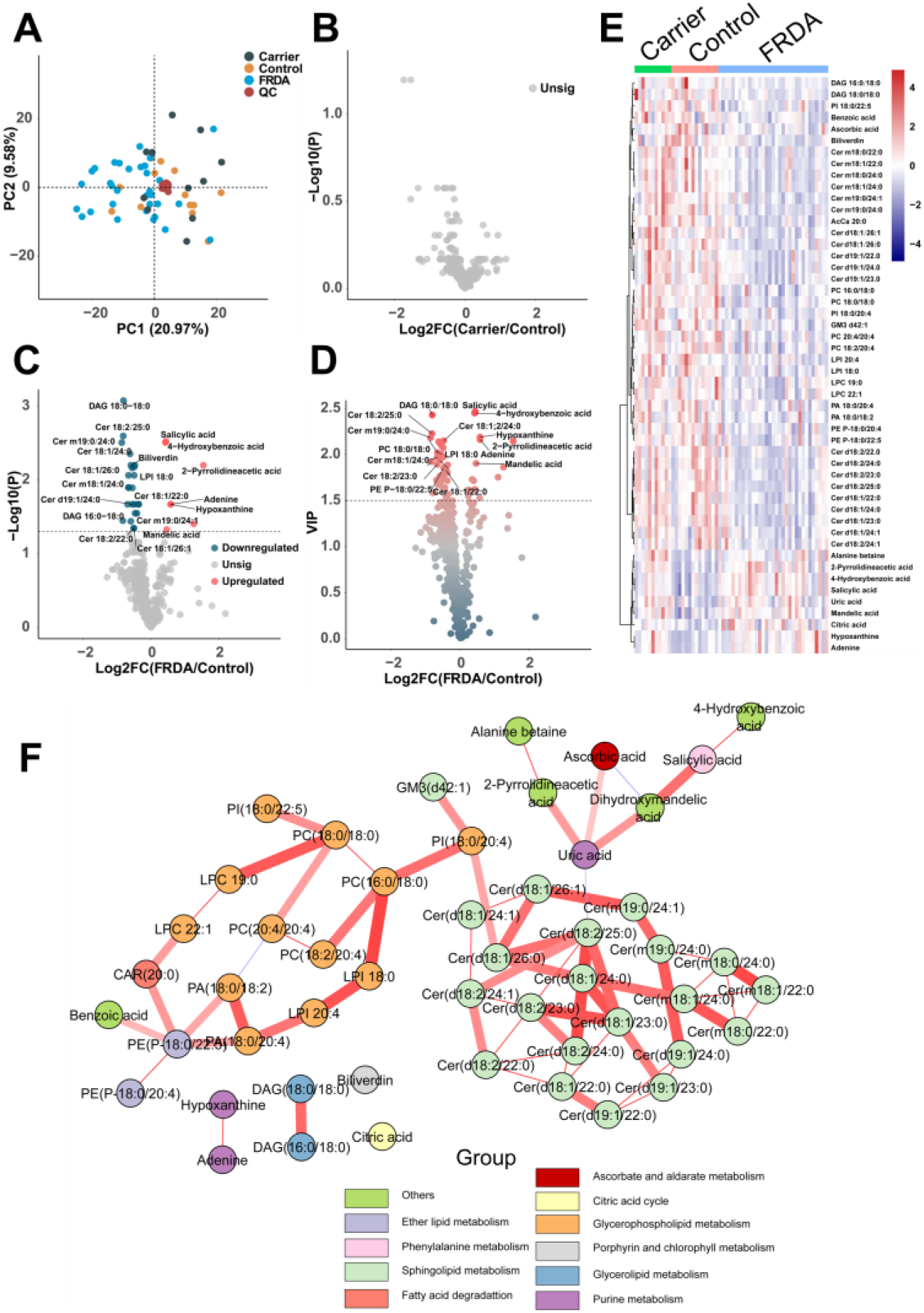
Metabolomics and lipidomics analysis revealed dysregulated metabolism in FRDA plasma. **A.** PCA score plot shows the plasma clustering according to disease conditions. Quality controls (QCs) were run every ten samples. The internal standard and QC-LOESS were combined to be used for signal drift correction. The closed clustering of the QC samples shows the robust methodology. **B.** Volcano plot of Carrier samples versus Control samples showed no statistical significance (-Log_10_P, two-sided *t*-test) against log2-fold change between the means. **C.** Volcano plot of statistical significance (-Log_10_P, two-sided *t*-test) against log2-fold change between the means of FRDA patients versus Controls. Upregulated metabolites (lipids) were colored with red, and downregulated ones with blue. **D.** Volcano plot of statistical significance (VIP, PLS-DA) against log2-fold change between the means of FRDA patients versus Controls. Metabolites (lipids) were colored according to VIPs from blue (low) to red (high). **E.** Heatmap showing relative abundances of top 50 metabolites or lipids (P < 0.05 and FDR < 0.1) in Control, Carrier, and FRDA groups. The abundances of metabolites (lipids) were mean-centered and unit-scaled. **F.** Metabolite correlation network using MetScape and Cytoscape. Each node represented one metabolite (lipid), and nodes were colored according to metabolite (lipid) pathways. The edge colored with red indicated positive correlation, while the edge with blue indicated negative correlation. The edge width represented the strength of partial correlation coefficient.

### Pathway analysis revealed abnormal sphingolipid metabolism in FRDA

We further examined the pathway changes by performing Metabolite Set Enrichment Analysis (MSEA). For lipids, ceramides, PCs, and PIs had the highest enrichment ratio (Fig. 3A). For metabolites, the most enriched ones were categorized into purine metabolism, citric acid cycle, and amino acid metabolism (Fig. 3B). To quantify the metabolic pathway activity, we performed PCA for each mapped KEGG pathway, and calculated PC1 score as the proxy of pathway activity (Fig.3C). Hierarchical clustering showed three main clusters of metabolic pathways. The downregulated pathways (cluster 1) in FRDA patients were mainly lipid metabolism related pathways, including sphingolipid metabolism, glycerophospholipid metabolism, and glycerolipid metabolism. The upregulated pathway (cluster 3) in FRDA patients included the citric acid cycle, amino acid metabolism, and fatty acid degradation. We then performed statistical analysis for several top altered pathway using pathway activity score (scaled PC1 score). Consistent with the results from MSEA, sphingolipid metabolism and glycerophospholipid metabolism decreased in plasma from FRDA patients, while citric acid cycle and amino acid metabolism were increased Fig. 3D).

**Fig. 3.**
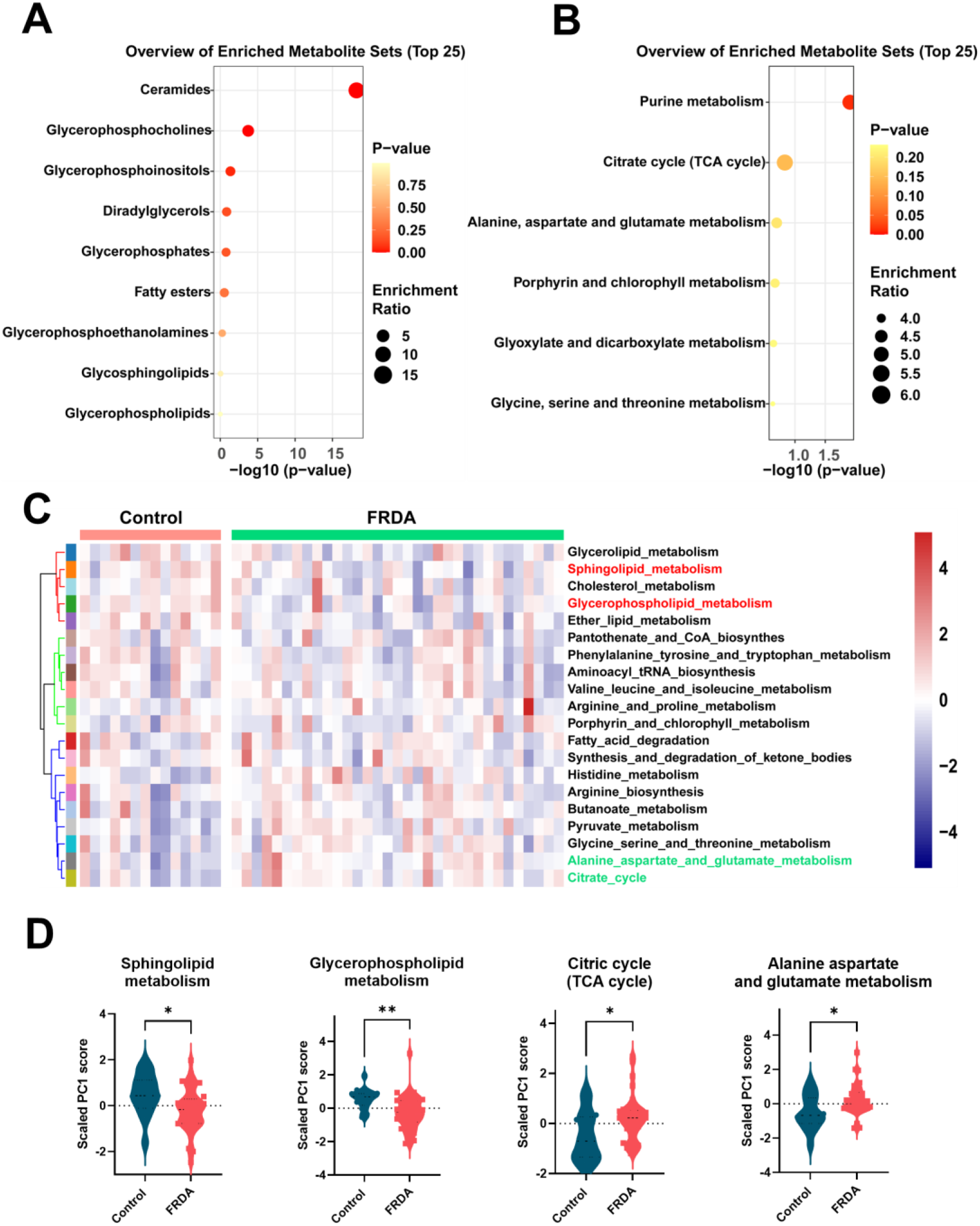
Pathway analysis indicated abnormal sphingolipid metabolism in FRDA. **A.** Lipid set enrichment analysis of significantly changed lipids (FDR adjusted P value less than 0.1). Enrichment Ratio is computed by Hits / Expected. **B.** Metabolite set enrichment analysis of significantly changed metabolites (FDR adjusted P value less than 0.1). Enrichment Ratio is computed by Hits / Expected. **C.** Metabolic pathway activity inferred by pathway score (PC1 scores) between healthy controls and FRDA patients. Hierarchical clustering of pathway scores were colored with pathway names labeled. **D.** Significant pathway scores changes of several metabolic pathways between FRDA patients and Controls. * p<0.05, ** p<0.005.

### Decreased very long chain ceramides distinguished FRDA patients from healthy controls

Ceramides were the most affected metabolites (lipids) in FRDA patients in this first discovery experiment. The long chain ceramides (C16 and C18 fatty acyl chain) showed no significant differences between FRDA patients and healthy controls (Fig. 4A, top left), while very long chain ceramide (C22 - C26 fatty acyl chain) were significantly decreased in FRDA patients (Fig. 4A). These differential regulations resulted in the increased ratio of long chain ceramide to very long chain ceramides (C16/C24 and C16/C26) (Fig. 4B), being an indicator of higher cardiometabolic risk as previously shown in non-FRDA patients ^32, 33^. To test the very long chain ceramides abilities to distinguish FRDA patients from healthy controls, we plotted the receiver operating characteristic (ROCs) analysis using different ceramides as predictor. All very long chain ceramides showed good predictive capacities with area under the curve (AUC) values from 0.75 to 0.85 (Fig. 4C).

**Fig. 4.**
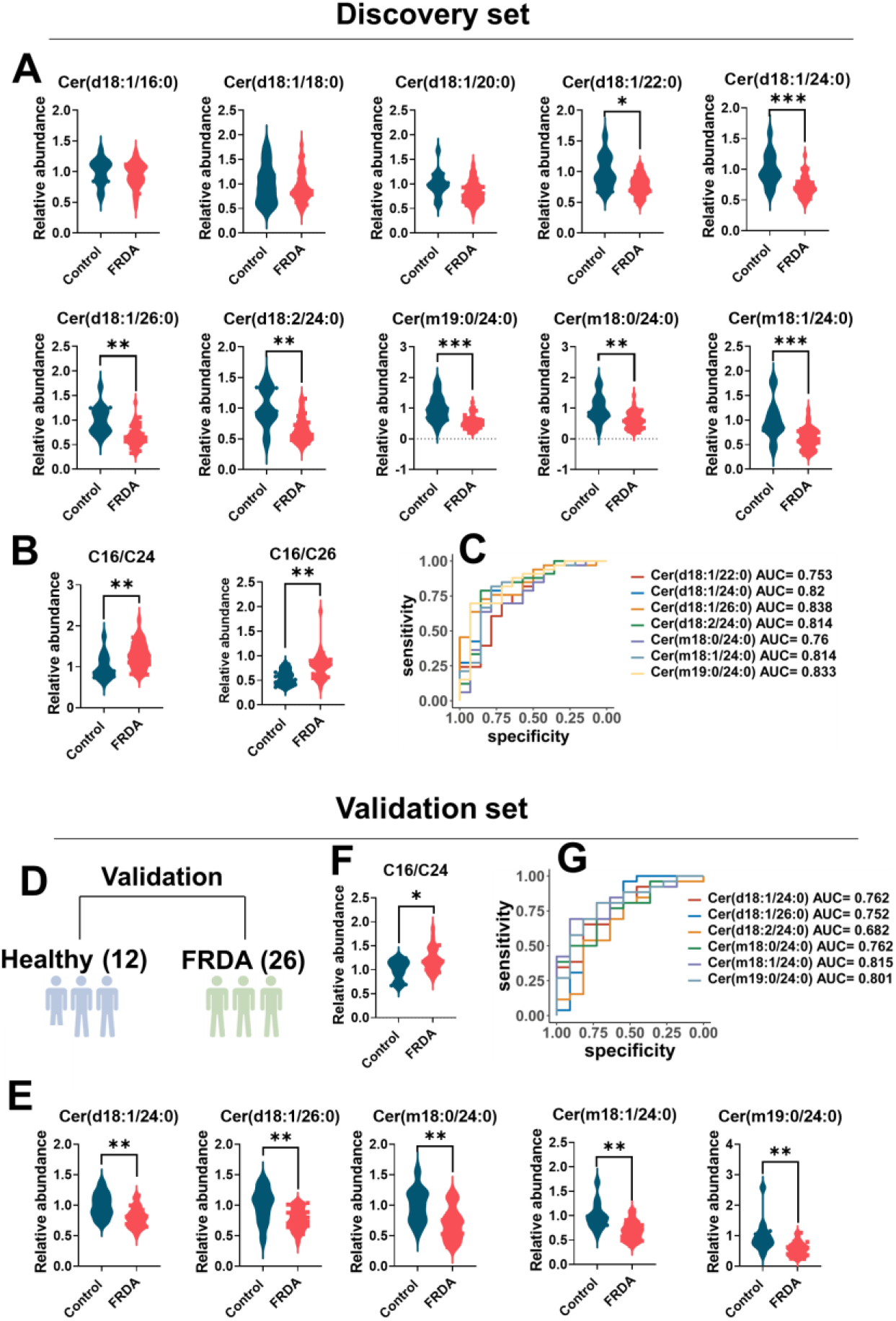
Decreased very long chain ceramides distinguished FRDA patients from healthy controls in two independent cohorts. **A.** Relative abundances of ceramides including long chain ceramides and very long chain ceramides between Controls and FRDA patients in the discovery set. **B.** The ratio of long chain ceramide (Cer(d18:1/16:0)) to very long chain ceramide (Cer(d18:1/24:0)) and (Cer(d18:1/26:0)) between Controls and FRDA patients in the discovery set. **C.** The receiver operating characteristic (ROC) curves for prediction of FRDA versus healthy controls using different ceramides, showed AUC values higher than 0.75. **D. Independently obtained plasma samples have been include in the** Validation set to verify the outcomes from the discovery set. **E.** Relative abundances of very long chain ceramides between heathy controls and FRDA patients in the validation set. **F.** The ratio of long chain ceramide (Cer(d18:1/16:0)) to very long chain ceramide (Cer(d18:1/24:0)) between heathy controls and FRDA patients in the validation set. **G.** The receiver operating characteristic (ROC) curves for prediction of FRDA versus healthy controls using different ceramides in the validation set, and AUC values were calculated and stated in plot. * p<0.05, ** p<0.005, *** p<0.005.

### Validation of very long chain ceramides as FRDA biomarkers in plasma

To validate the discovery results an independent cohort of EDTA fasting plasma was collected. The new cohort was collected between 2020-2022with plasma isolated within 1 h after draw in a research lab (as opposed to the clinical lab that did the isolation for the discovery cohort with no known timing associated between draw and isolation). This validation set did not sample from carriers as we detected no metabolic biomarkers that differentiated Carriers from the Controls (Fig. 2B). The validation set had similar numbers of controls and FRDA patients (Fig. 4D) as the discovery set (Supplementary Table 1). In the validation set, we found that very long chain ceramides were also significantly decreased in the FRDA patients (Fig. 4E and Supplementary Fig. 6A). The validation set also showed the ratio ceramides C16/C24 significantly increased (Fig. 4F) predicting a higher cardiometabolic risk ^31, 32^. The predictive powers from the ROC (Fig. 4G) remained similarly strong compared to the discovery set with AUC from 0.7 to 0.82, suggesting very long chain ceramides as plasma biomarkers of FRDA.

### Quantification of upstream and downstream metabolites from known deficient enzymes

Because frataxin is directly involved in the regulation of citric acid enzyme aconitase, we quantified the levels of upstream and downstream metabolites of this enzyme. The upstream metabolites citric acid and aconitic acid increased in both the discovery and validation sets (raw P value < 0.05), while the downstream metabolite iso-citric acid was not significantly different (Supplementary Fig. 6B,C). The alteration of citric acid intermediates indicated that enzyme activity is directly reflected in the plasma concentration.

### Unbalanced ceramide synthesis in frataxin-deficient cardiomyocytes

To investigate if the alterations of ceramides in FRDA plasma reflects the dysfunction of clinically affected tissues, we used siRNA to knockdown FXN in human induced pluripotent stem cell-derived cardiomyocytes (iPSC-CMs) and then examined ceramide metabolism (Fig. 5A). We used two independently derived iPSC-CM lines. Line SV20 was obtained the Penn iPSC Core Facility and line 11713 was obtained from Fujifilm Cellular Dynamics (see Materials and Methods). FXN knockdown using two different siRNA, dramatically decreased the frataxin protein levels in both cell lines (Fig. 5B,C and Supplemental Fig. 7A, using an additional cell line). Lipidomics analysis showed that both lines of frataxin-deficient cardiomyocytes had significantly decreased very long chain ceramides (Fig. 5D,E). For line 11713, higher scalability allowed us to perform lipidomics analysis with two siFRX KDs that demonstrated the same trends versus control (Fig. 5E,G). The unbalanced alterations of ceramides increased the ratios of long chain to very long chain ceramides (Fig. 5F,G), similar to prior observations in the fibroblast cells from FRDA patients ^34^. The *de novo* synthesis of long chain ceramides, and very long chain ceramides is regulated by different ceramide synthase (CerS1-6) that each have tissue and acyl-CoA chain length specificity. Ceramide synthase 2 (CerS2 or LASS2) preferentially select very long chain fatty acyl-CoA (C20-26) as the substrate ^35^. Consistent with the lipidomics results, CerS2 protein levels decreased in iPSC-CMs after FXN knockdown in comparison with control (Fig. 5H and Supplementary Fig.. 7 B,C).

**Fig. 5.**
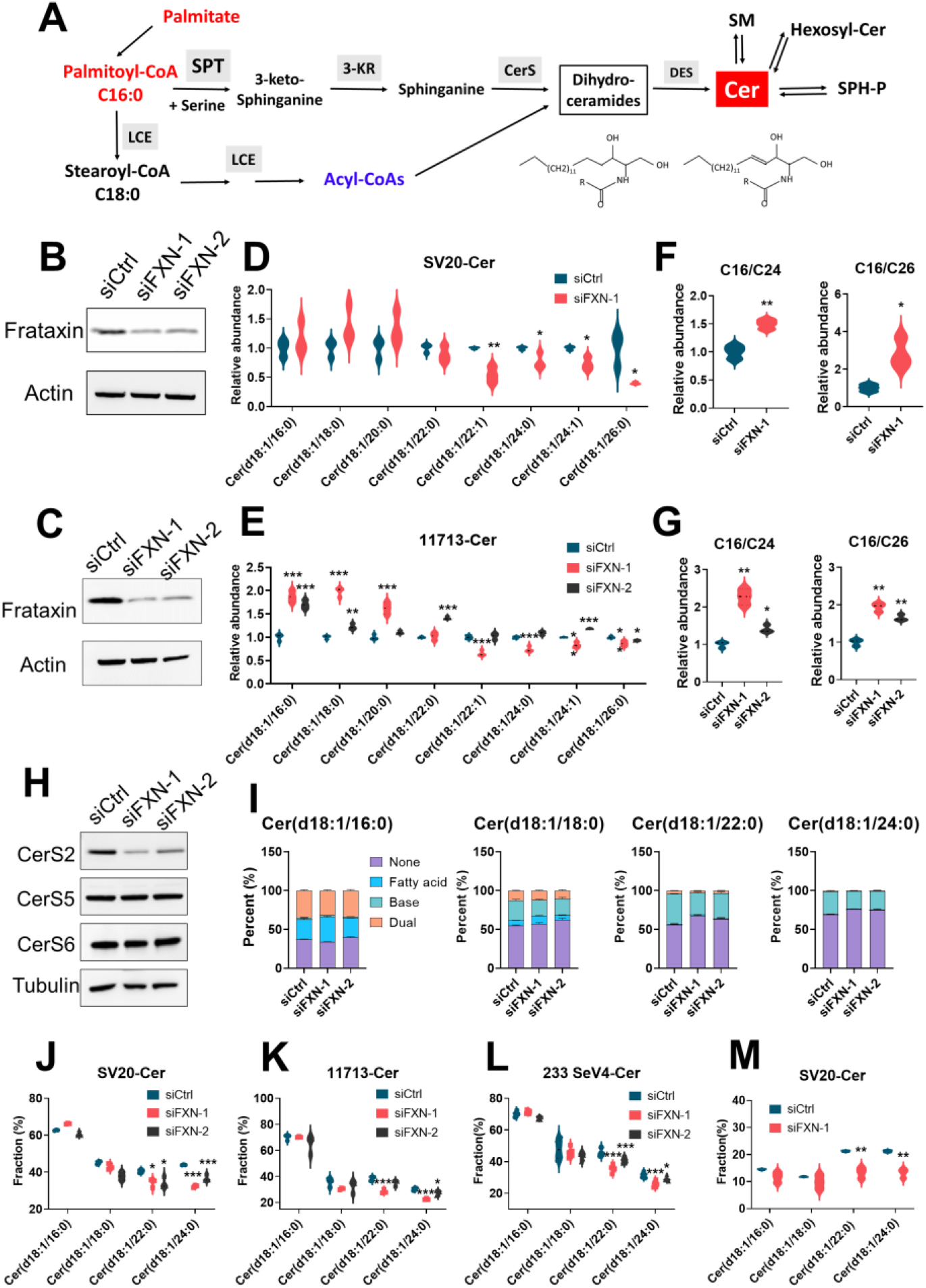
Unbalanced ceramide synthesis in several lines of frataxin-deficient cardiomyocytes. **A.** Schematic depiction of *de nova* ceramide synthesis **B.** Western blots showing the frataxin levels in FXN-knockdown cardiomyocyte SV20 **C.** same as B but for a different cell line, 11713. **D.** The relative abundances of ceramides in cardiomyocyte SV20 and **E.** 11713 (after siRNA-mediated FXN knockdown. **F.** The ratios of long chain ceramide (C16) to very long chain ceramide (C24 and C26) in cardiomyocyte SV20 and **G**. in cardiomyocytes 11713, both after FXN-knockdown. **H.** Western blots showing the protein levels of ceramide synthase (CerS2, CerS5, and CerS6) in cardiomyocyte SV20 after siRNA-mediated FXN knockdown **I.** The mass isotopomer distribution of ceramides from FXN-knockdown cardiomyocyte SV20 **J.** The isotopic labeling fractions of ceramides in FXN-KD cardiomyocytes SV20 **K.** in cardiomyocytes 11713 and **L**. cardiomyocytes 233-SeV4, all after ^13^C_16_-palmitate labeling. **K.** The isotopic labeling fractions of ceramides in FXN-KD cardiomyocytes SV20 after ^13^C_3_,^15^N-serine labeling. * p<0.05, ** p<0.005, *** p<0.005.

To further test unbalanced ceramide regulation in frataxin-deficient cardiomyocytes, we performed ^13^C_16_-palmitate based isotope tracing experiments using a sensitive LC-HRMS targeted method ^34^ to quantify the mass isotopomer distribution of ceramides. Long chain ceramides (C16,C18) were not significantly changed after FXN knockdown, while very long chain ceramides (C22 and C24) had lower fractions of the labeled form in frataxin-knockdown cardiomyocytes (Fig.. 5I and Supplemental Fig. 7D,E). The isotopic labeling fractions of ceramides in FXN-KD cardiomyocytes had similar patterns in the three different cell lines: SV20 (Fig. 5J), 11713 (Fig. 5K), and an additional line from the Penn core, 233 SeV4 (Fig. 5L) All cell lines showed around 40% incorporation from ^13^C_16_-palmitate in the very long chain ceramides and all were significantly reduced in 12h (Fig. 5J-L). As the precursors in ceramides biosynthesis are palmitate and serine (Fig. 5A), we also used ^13^C_3_,^15^N-serine tracing in one of the cell lines (SV20) and similar results were observed after frataxin knockdown, with decreased labeling fractions of very long chain ceramides (Fig. 5M). All these data indicated that frataxin deficiency in cardiomyocytes led to unbalanced ceramide biosynthesis, with CerS2 being reduced at protein levels and enzymatic activity.

### Ceramides were differentially regulated in primary skeletal muscle cells

To examined if, ceramide dysfunction was a ubiquitous outcome of frataxin deficiency in another affected FRDA tissues, we chose human primary skeletal muscle cells (HSkMC). We used three different siRNA-mediated FXN knockdown (Fig. 6A) and performed the lipidomics and ^13^C_16_-palmitate based isotope-tracing experiments. For HSkMC, FXN knockdown significantly increased the levels of ceramides, including both long chain and very long chain ceramides, with long chain ceramides increasing more significantly than very long chain ceramides (Fig. 6B). The ratios of long chain to very long chain ceramides increased in FXN-knockdown HSkMC (Fig. 6C), similar to results observed in frataxin-deficient cardiomyocytes, but the increase was most likely driven by the large increase in the long chain ceramides. For isotopic labeling experiments, FXN knockdown increased the fraction of isotope labeled forms (fatty acid chain, sphingosine base, and dual) for most of ceramides based on mass isotopomer distribution data (Fig. 6D). The isotopic labeling fractions were dramatically increased in FXN-KD cells for all ceramides except Cer(d18:1/24:0) in HSkMCs (Fig. 6E), indicating that the FXN knockdown differentially regulated ceramides in cells originating from different affected tissues.

**Fig. 6.**
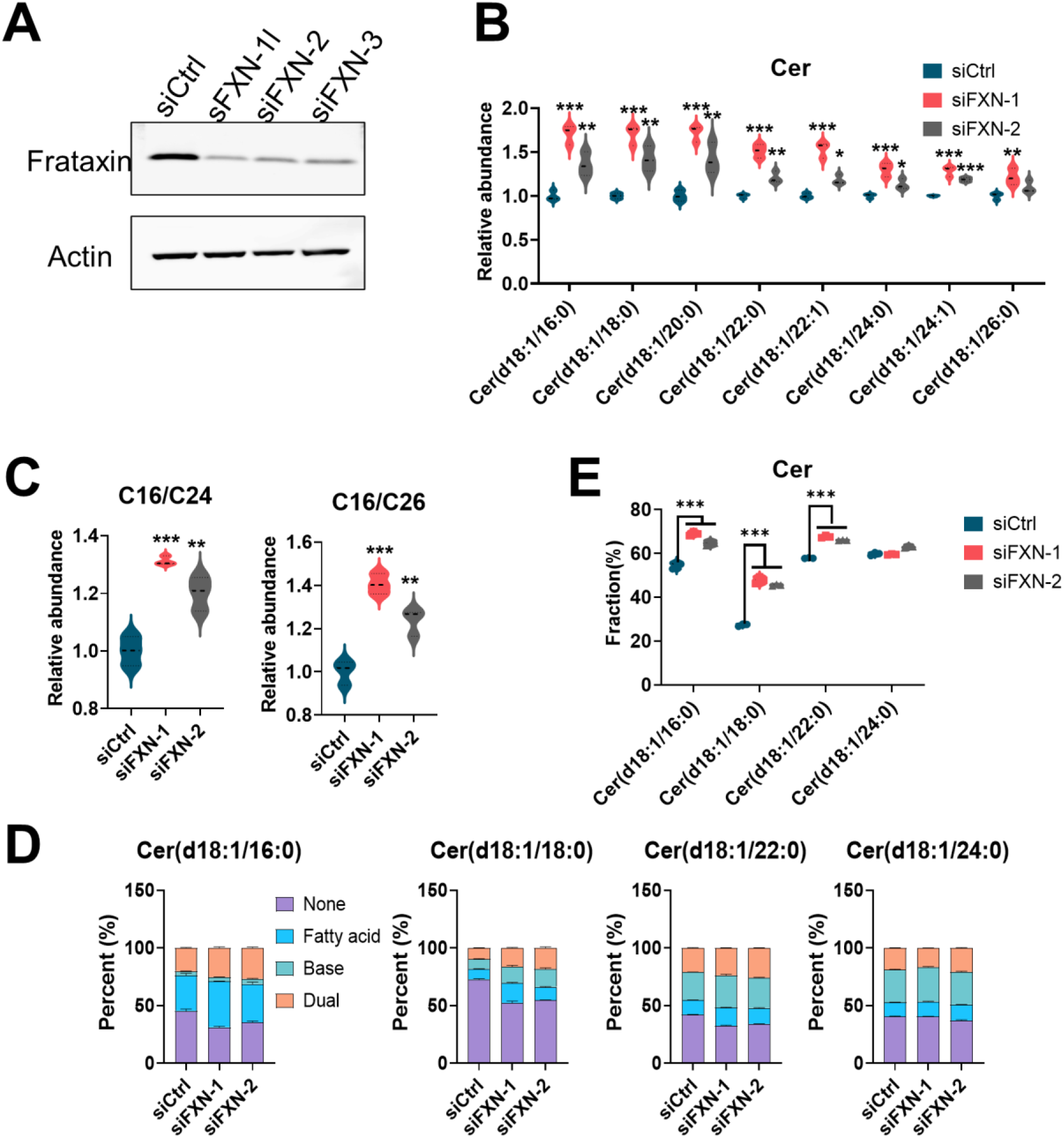
Frataxin deficiency had different effect on the ceramide metabolism in primary skeletal muscle cells. **A.** Western blots showing the frataxin levels in human primary skeletal muscle cells (HSkMC) after siRNA-mediated FXN knockdown **B.** The relative abundances of ceramides in FXN-knockdown HSkMC. **C.** The ratios of long chain ceramide (C16) to very long chain ceramide (C24 and C26) in FXN-knockdown HSkMC **D.** The mass isotopomer distribution of ceramides from control (siCtrl) and FXN-Knockdown HSkMCs. **E.** The isotopic labeling fractions of ceramides in FXN-knockdown HSkMC. * p<0.05, ** p<0.005, *** p<0.005.

### Proteomics revealed decreased apolipoprotein levels

Using a similar targeted omics workflow, we performed proteomics analysis for the plasma discovery set. By inspecting for contamination biomarkers for coagulation, erythrocyte, and platelet ^36^, our samples appeared to reflect high quality plasma, except for two samples contaminated by platelets (Supplementary Fig. 8). When the quantified proteins were plotted in a heatmap (Fig. 7A), the plasma proteins measured cannot distinguish FRDA patients from healthy controls in the discovery set (Fig. 7B). Statistical analysis revealed 9 proteins that were significantly altered in FRDA patients in comparison with healthy controls, with 8 decreased and 1 increased (FDR < 0.05) in FRDA patients (Fig. 7C). Most of these dysregulated proteins belonged to the apolipoprotein family (Fig. 7D). However, only one apolipoprotein (APOD) could be verified in the validation set (Fig. 7E). A previous report using a stable isotope dilution quantification of APOA1 in FRDA serum samples also observed the decrease of APOA1 ^37^. Considering the label-free quantification strategy and the smaller number of samples in the validation set a more targeted method with a larger number of samples could help the validation of additional apolipoproteins in plasma similar to the reduced levels of ApoA in FRDA serum^37^.

**Fig. 7.**
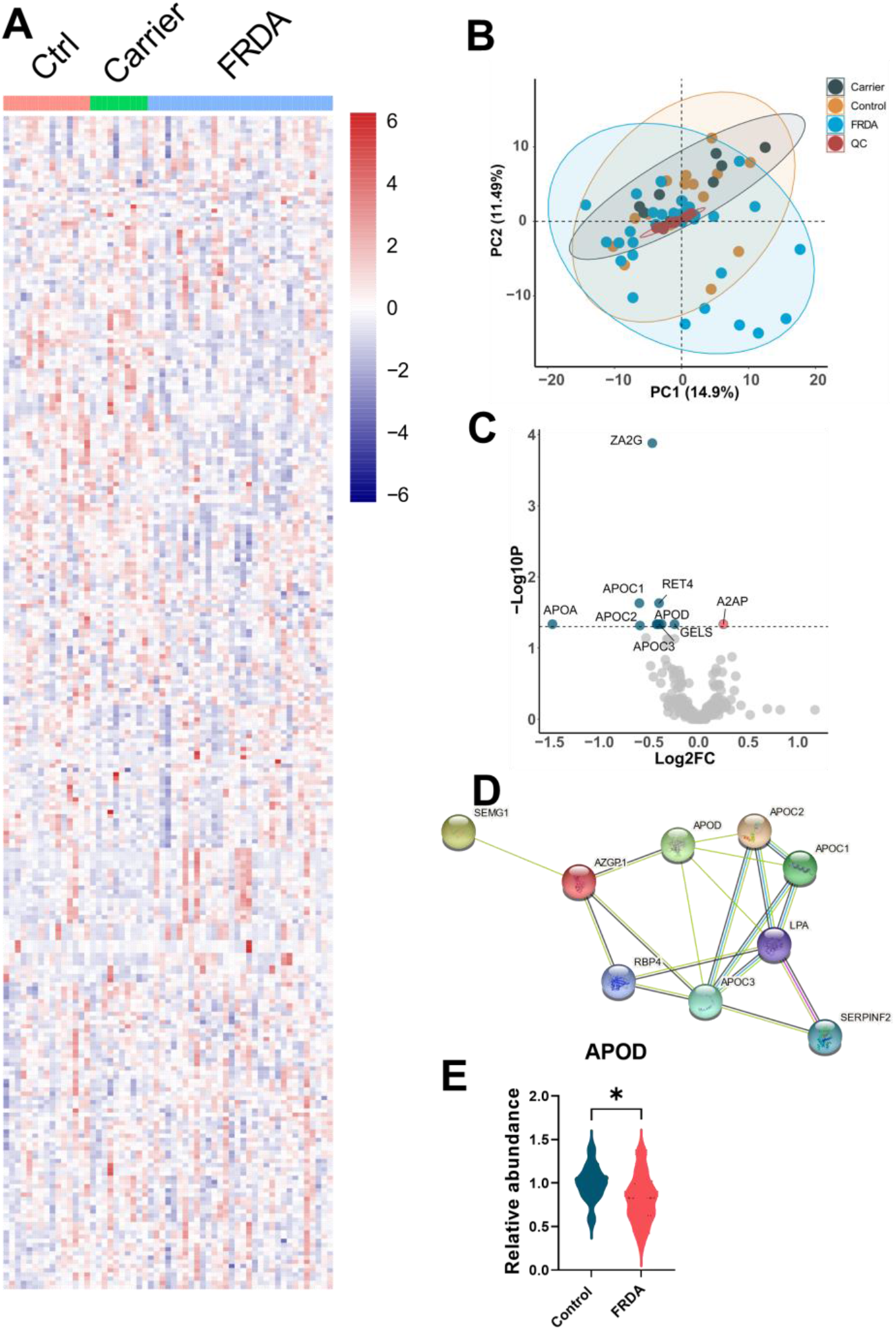
Proteomics analysis showed decreased apolipoprotein levels in FRDA plasma. **A.** Heatmap showing hierarchical clustering of peptides (proteins) in Control, Carrier, and FRDA groups. **B.** PCA score plot showing the plasma clustering according to disease conditions, and quality controls (QCs) were run every ten samples. The QC-LOESS were used for signal drift correction. **C.** Volcano plot of statistical significance (-Log10P, two-sided t-test) against log2-fold change between the means of FRDA patients versus healthy controls. Upregulated proteins were colored with red, and downregulated ones with blue. **D.** Protein-Protein Interaction Networks of significantly changed proteins (FDR corrected P value less than 0.05) using String. **E.** APOD was the only significantly decreased protein in FRDA patients versus healthy controls in the validation set. * p<0.05.

### Machine learning identified metabolic features for FRDA prediction

To improve the predictive powers of ceramides as biomarkers for FRDA, we performed machine learning based optimization (Fig. 8A). First, we included 26 metabolites that were robustly altered in FRDA patients in both, discovery set, and validation set. Next, we chose the top 4 metabolites by linear support vector machine (SVM) based feature selection strategy, namely Cer(m19:0/24:0), 3-methoxytyrosine, Cer(m18:1/22:0), and putrescine (Fig. 8B). To reduce the redundancy, a k-means clustering (KM) was performed to exclude Cer(m18:1/22:0) that showed similar behavior with Cer(m19:0/24:0), creating a final model with three metabolites (Cer(m19:0/24:0), 3-methoxytyrosine, and putrescine). Using three metabolites, we optimized linear SVM models using the discovery set and verified the model in the validation set. The ROC curves showed that these three-metabolite combination improved the predictive capability, with AUC as high as 0.924 (0.850-0.998) in the discovery set (Fig. 8C) and 0.923 (0.842-1.000) in the validation set (Fig. 8D).

**Fig. 8.**
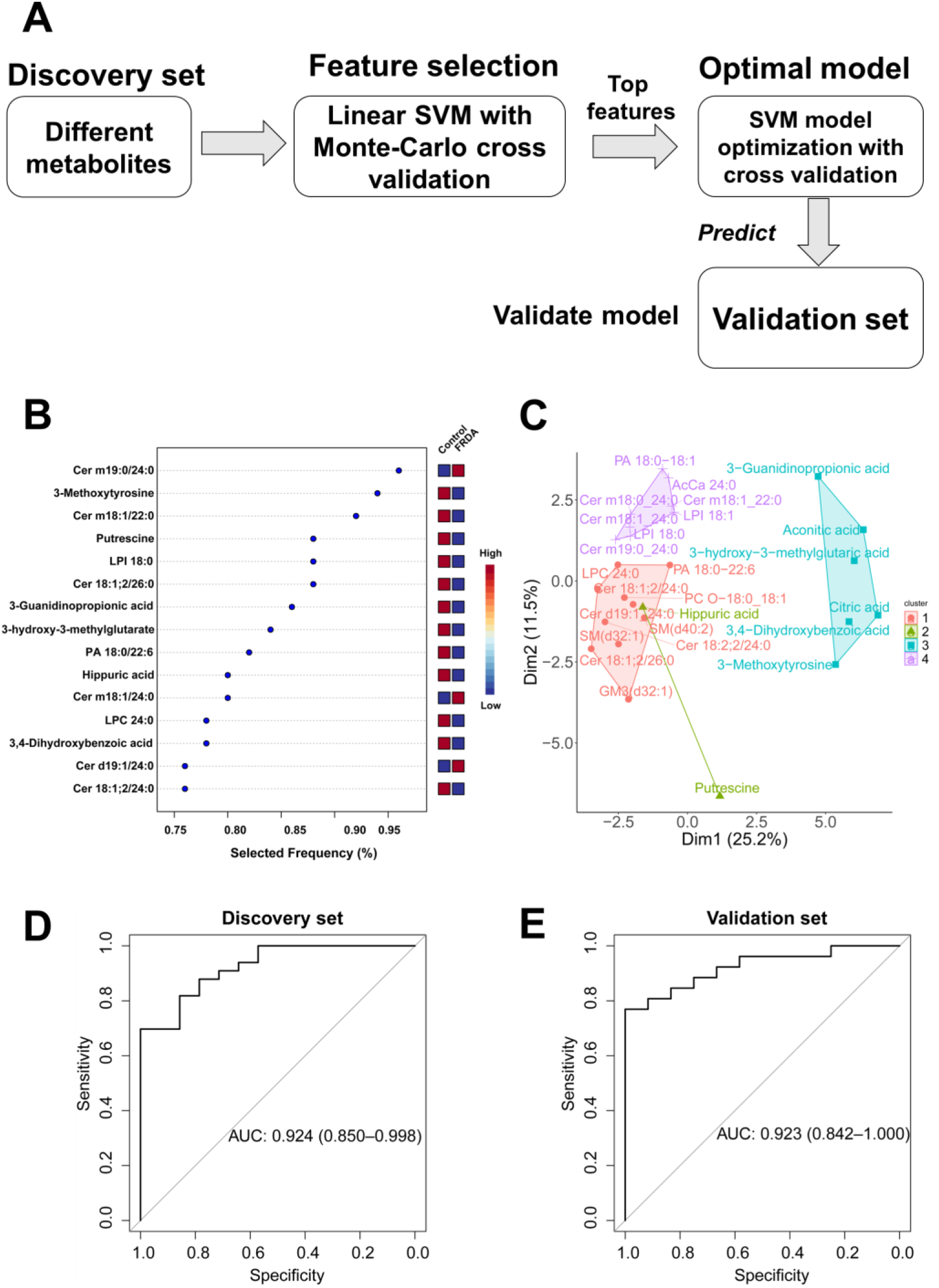
Machine learning improves the prediction of FRDA. **A.** Machine learning workflow for prediction of FRDA using metabolic features. **B.** Top 15 selected metabolites from Linear support vector machine (SVM) model. **C**. K-Means Clustering revealed different 4 clusters for these metabolites. **D.** The receiver operating characteristic (ROC) curves using three metabolites [Cer(m19:0/24:0), 3-methoxytyrosine, and putrescine] based SVM model in the discovery set **E**. same as D but in the validation set.

## DISCUSSION

FRDA is a multi-systemic mitochondrial disease caused by the frataxin deficiency. The important role of frataxin in the iron-sulfur cluster assembly results in multiple altered metabolic pathways in FRDA. However, there is no readily available metabolic biomarker adequately validated for use in human blood samples. To identify metabolic features as potential biomarkers, we performed multi-omics profiling and evaluated systematic metabolic changes in FRDA fasting plasma samples compared with healthy controls. Multiple metabolic pathways, including sphingolipid metabolism, phospholipid metabolism, citric acid cycle, and apolipoprotein metabolism were consistently affected in FRDA plasma samples (Fig. 2F).

In our lipidomics analysis, long chain ceramides were unchanged in FRDA plasma (C16-20), while very long chain ceramides (C22-26) were significantly decreased (Fig. 4A,F), resulting in the increased ratio of long chain ceramide to very long chain ceramides (Fig. 4B,G). The decreased very long chain ceramides were validated in an independent cohort, indicating the robust results of our multi-omics studies. An increasing number of clinical studies are reporting associations between ceramides and cardiovascular outcomes, to the extent that the Mayo Clinic is marketing diagnostic tests to measure ceramides ^32^. Of note, unbalanced ceramide levels, with preferentially decreased very long chain ceramide, were first observed in human heart tissue collected post-mortem from FRDA patients, but the number of tissue samples used was very low (n=6) and this tissue is obviously not routinely accessible ^25^.

The preferential depletion of very long chain ceramide along with enrichment of long chain ceramides were replicated in frataxin-deficient human cardiomyocytes (Fig. 5D,E) from two independent sources. Furthermore, frataxin deficient cardiomyocytes consistently showed the enrichment of long chain ceramide and depletion of very long chain ceramide. The biosynthesis enzyme CerS2 that is responsible for preferential synthesis of very long chain ceramide was decreased after FRX KD (Fig. 5H). Long-chain ceramide (C16) is usually thought to be deleterious, whereas very-long-chain ceramides (such as C24:0) are considered to be benign or protective ^38–41^. Ceramides act as a second messenger in the apoptotic cascade, with long chain ceramides (C16 and C18) being the main ceramides activators of apoptosis. CerS5 or CerS6 (long chain ceramide synthesis) overexpression leads to more cells undergoing apoptosis whereas CerS2 overexpression protected cells from death ^42, 43^. Heart-specific Sptlc2-deficient (hSptlc2 KO) mice showed decreased C18:0 and very long chain ceramides but led to cardiac dysfunction ^44^. Therefore, balanced ceramide metabolism is important for normal functions; thus, unbalanced ceramides in FRDA may also contribute to the pathophysiology of cardiac hypertrophy or heart failure in FRDA. Further studies testing the effects of modulating different ceramides on cardiac functions of FRDA could answer this question. Very recently, a potent N-acetylgalactosamine-conjugated antisense oligonucleotide engineered to silence CerS2 in hepatocytes *in vivo*, prompts the modulation of blood plasma ceramides ^45^. This recent study points that circulating ceramides can be modulated by ceramide biosynthesis in hepatocytes.

Our studies provide very long chain ceramides as robust biomarkers for FRDA patients. Ceramide dysfunctions are involved in many neurodegenerative and cardiovascular diseases ^46–49^. Targeting ceramide metabolism can ameliorate neurological symptoms and cardiac hypertrophy and heart failure ^50, 51^. In FRDA, ceramides accumulated in a fly model and were associated with neurodegeneration in FRDA ^24, 25^, so we did not test the effect of frataxin knockdown in neurons due to these robust pre-existing data.

However, ceramide metabolism was differentially regulated in human primary skeletal muscle cells in response to frataxin knockdown (Fig. 6A,B), with the skeletal muscles being one of the first affected tissues in FRDA. In contrary to the finding in the cardiomyocytes, where only long chain ceramide increased after frataxin deficiency, most of ceramides were increased in the skeletal muscle cells. Noteworthy, C18 ceramide, which is mediated by CerS1- an isoform highly expressed in muscles, was the most increased ceramide followed by C16 ceramides. In skeletal muscle, elevated levels of long chains ceramides (such as C16:0 or C18:0) was demonstrated to induce insulin resistance and hepatic steatosis, whereas very long chain ceramides (for instance, C24:0 or C24:1) do not ^39, 40, 52^. The dramatic increase in C16 and C18 long chain ceramides in skeletal muscles cells, if replicated in the muscles, could add to the increased risk of diabetes and insulin resistance in FRDA patients, also driven by the lack of beta cells in FRDA. The tissue-specific alterations in FRDA were also revealed in a functional genomic analysis of heart and muscle tissues in FRDA mice models ^23^. Therefore, tissue-specific intervention strategy should be considered for targeted therapy. Muscles tissues analysis would be easier available through muscles biopsy, than heart or brain tissue.

Frataxin deficiency directly affected Fe–S containing enzyme aconitase and respiratory chain complexes. Decreased aconitase activity was observed in FRDA heart tissues and yeast models ^18^. However, blood cells (lymphocytes, platelets, and peripheral blood mononuclear cell) from FRDA patients failed to replicate this finding though frataxin protein levels were significantly decreased in whole blood ^53^ or different subtypes of blood cells ^54, 55^. In our metabolomics analysis, we expectedly observed the increase of citric acid and aconitic acid, the upstream metabolites of aconitase in FRDA plasma, indicating defective activity of this enzyme. The alteration of citric acid cycle pathway activity was also demonstrated in our metabolomics analysis (Supplementary Fig. 6). These metabolic differences validate that plasma samples are relevant to the metabolic conditions of FRDA patients.

There are several limitations in this study, mostly deriving from the relatively small number of subjects in both cohorts. Although these small cohort samples showed robust metabolic and lipidomic alterations in FRDA patients, the numbers were not enough to reveal the proteomic changes. This may be due to the fact that no internal standards were used for the proteomics, workflow, but at least one internal standard per chemical class was used for the small molecule omics. Decreased levels of several apolipoproteins from our discovery cohort failed to validate in the validation cohort (8 out 9 could not be validated). By comparing the APOA1 levels between FRDA patients and healthy control in a larger cohort ^37^, APOA1 levels were significantly decreased in FRDA serum samples. It appears that a larger cohort is necessary for robust proteomics biomarkers, due to the inherent challenges in measuring plasma proteins. This also seems to support the utility of small molecules biomarkers for clinical studies. We did demonstrated decreased long chain ceramides as robust biomarker for FRDA patients and replicated this finding in frataxin-deficient cardiomyocytes, consistent with of the reduced long chain ceramides in FRDA heart tissue ^25^ but direct correlation between plasma ceramide levels and heart ceramide levels in human subjects is not readily achievable. Further studies should be done to clarify if dysregulated ceramide levels can be potential biomarkers of cardiomyopathy in FRDA patients. Hammand et al ^56^ showed that the C24 ceramides are not affected by the fasting state, so the decreased ceramides biomarkers observed here open the possibility to utilize larger cohorts of archived non-fasting plasma samples, and evaluate their levels longitudinally. Due to their high blood concentrations (around 3 µM), excellent ionization, and specific fragmentation pattern in collision induced dissociation, very long chain ceramides are feasible for translation into a routine lab assay in clinical settings.

In summary, by performing multi-omics profiling in fasting plasma samples, we identified and validated the very long chain ceramides as robust metabolic biomarkers for FRDA patients. We also demonstrated the clear reduction in the ceramide synthase 2 in frataxin-deficient cardiomyocytes that lead to the depletion of very long chain ceramides but observed different ceramide alterations in human primary skeletal muscle cells. Tissue-specific ceramide alterations should be considered when testing therapeutic approaches. Moreover, we applied machine learning based optimization, indicating that very long chain ceramide in combination with other two metabolites can improve the prediction of FRDA.

## MATERIALS AND METHODS

### Human blood sample collection

Plasma samples were collected in two different cohorts all enrolled in parallel in an ongoing natural history study at the Children’s Hospital of Philadelphia. The blood samples were collected after overnight fasting, all before 10 AM the next morning. Written informed consent was obtained from each donor participating in the study and possible consequences of the studies were explained. If subjects were under the age of 18, written informed consent was obtained from a parent and/or legal guardian. The study was approved by the Institutional Review Board (IRB) of the Children Hospital of Philadelphia (IRB Protocol # 01–002609). Venous blood was drawn in 8.5 mL purple cap Vacutainer EDTA tubes and gently invert to mix. Plasma was collected after centrifugation at 1000xg for 15 min. All samples were immediately aliquoted to Eppendorf tubes and frozen at −80 °C until analysis. The discovery cohort was collected between 2012-2019 (Table S1). The validation cohort was collected between 2020-2022 (Table S1).

### Chemicals and reagents

Optima LC/MS-grade methanol (A456-4), water (W6500), acetonitrile (A955-4), isopropanol (A461-4), formic Acid (A117-50), and ammonium formate (A11550) were purchased from fisher scientific (Waltham, MA, USA). HPLC-grade methyl tert-butyl ether (MTBE, E127-4) and 1-Butanol (A383SK-1) were purchased from Fisher Scientific (Waltham, MA, USA). Labeled internal standards were obtained as showed in Supplementary Table 2. Bovine Serum Albumin (fatty acid free, low endotoxin, lyophilized powder) (A8806), 2-Iodoacetamide (IAA, 8047440100), urea (U0631), ammonium bicarbonate (A6141), and DL-Dithiothreitol (DTT, D0632) were purchased from Sigma-Aldrich (St. Louis, MO, USA). Pierce™ Trypsin Protease, MS Grade (90058) was purchased from Thermo Scientific (Waltham, MA, USA).

#### Human induced pluripotent stem cell differentiated cardiomyocytes (iPSC-CMs)

The iPSC-CMs line SV20 and 233 SeV4 were obtained from the Penn iPSC Core Facility. The iPSC-CMs (SV20 and 233 SeV4) were replated into 12 well plates (0.1% gelatin coated) in RPMI/20%FBS/1µM thiazovivin overnight and replaced with RPMI/B27+ for 2 days to recover. All the experiments for these two lines were conducted in RPMI/B27 medium. The iCell Cardiomyocytes 11713 (R1106) was ordered from Fujifilm Cellular Dynamics and maintained in iCell Cardiomyocyte Maintenance Medium (M1003) for all the experiments. We received the cell lines from the Penn iPSC and Fujifilm several times over 6 months period for repeated experiments, and each experiment had at least 3 technical replicates.

#### Human primary skeletal muscle cell culture

Human skeletal muscle cells (HSkMC, PCS-950-010) were ordered from ATCC (Manassas, VA, USA) and maintained in Mesenchymal Stem Cell Basal Medium (PCS-500-030, ATCC) supplemented with Primary Skeletal Muscle Growth Kit (PCS-950-040, ATCC).

### siRNA knockdown

Silencer® Select siRNAs (s5361-siFXN-1, s530978-siFXN-2, and s5362-siFXN-3) targeting gene FXN and Silencer™ Negative Control No. 1 (4404021-siCtrl) were purchased from Thermo Scientific (Waltham, MA, USA). FXN siRNA and negative control siRNA were transfected into targeted cells using Lipofectamine™ RNAiMAX Transfection Reagent (13778075, Invitrogen™) at concentrations of 10-50 nM according to the manufacturer’s instructions. The concentrations of siRNAs were optimized to bring down the frataxin protein levels to less than 50% of negative control cells. The knockdown efficiency was verified by western blot.

### Stable isotope tracing and cell lipidomics

iPSC-CMs and HSkMCs were cultured in their respective fresh medium containing 100 μM ^13^C_16_-palmitate (conjunction with fatty acid free BSA with ratio 2:1) for 6 h or 24 h. Then cells were collected into 1 mL cold 80% methanol after washing 2x with 5 mL PBS. 2.4 mL MTBE and 0.6 mL water were sequentially added and vortex for 1 min. The phase separation was achieved after centrifugation at 3000x*g* for 10 min. The top organic phase was collected, dried under nitrogen, and reconstituted in MTBE/methanol (1:3, v/v) with 10 mM ammonium formate for LC-HRMS analysis. A parallel reactive monitoring (PRM) targeted method was used to quantify the mass isotopomer distributions of ceramides according to our previous method ^34^.

For cell lipidomics analysis, the same protocol was used for sample preparation (iPSC-CMs and HSkMCs) after cells being spiked with the SPLASH® LIPIDOMIX® Mass Spec Standard (330707) and Cer/Sph Mixture I (LM6002). Data acquisition was performed using the sample method described in Lipidomics Section except the full scan mode was used for ceramides quantification.

### Western blot

Cells were lysed in RIPA buffer (150 mM NaCl, 1% NP-40, 20 mM Tris-HCl pH 7.5, 1 mM EGTA, 1% sodium deoxycholate, 1 mM Na_2_EDTA, 2.5 mM sodium pyrophosphate, 1mM beta-glycerophosphate, 1 mM Na_3_VO_4_, 1μg/ml leupeptin) containing protease inhibitors (A32963, Thermo Scientific). Total protein content was determined using the Pierce^TM^ 660nm protein assay reagent (22660, Thermo Scientific). 20 μg of total protein (3:1 protein/loading buffer) was boiled at 95 °C for 5 min and then loaded onto NuPAGE 10% or 12% Bis-Tris gel. Following SDS-PAGE, proteins were transferred onto nitrocellulose membrane, blocked with 5% dry milk and incubated with primary antibodies against frataxin (1:500) and other target proteins (1:500-1:2000) overnight at 4 °C. Blots were then incubated with appropriate horseradish peroxidase-conjugated secondary antibodies and developed using ultra-sensitive enhanced chemiluminescent SuperSignal West Femto (34094, Thermo Scientific). Lass2 polyclonal antibody (PA5-115496), Lass5 polyclonal antibody (PA5-95899), Lass6 polyclonal antibody (PA5-20648), beta Tubulin Monoclonal Antibody (MA5-16308), and Actin monoclonal antibody (MA1-744) were purchased from Thermo Scientific (Waltham, MA, USA). FXN Polyclonal antibody (14147-1-AP) was purchased from Proteintech (Rosemont, IL, USA). Full Wester Blots are in Supplementary Fig. 9.

### Flow cytometry

iPSC-CMs was dissociated with Trypsin-EDTA, and fixed in 4% PFA for 10 min, and permeabilized in PBS containing 0.3% Tween 20 for 1h. Permeabilized cardiomyocytes were stained with a primary mouse anti-Troponin antibody (Abcam Ab8295) at dilution 1:200 overnight followed by goat anti mouse-Alexa Fluor 488 conjugated (Invitrogen 11001) at dilution 1:1000 for 1h. Data were acquired on an Amnis Image Stream and analyzed using open source Flowing Software (Turku Centre for Biotechnology). We routinely used iPSC-CMs that were 80% or above c-troponin positive. Nuclei were stained with DAPI (#P36935, Thermo Fisher Scientific). Not stained cells were used for negative control.

### Metabolomics

20 μL plasma, spiked with 30 μL internal standard (Supplementary Table 2), was vortex mixed with 200 μL methanol for 10 min, and incubated at −20°C for 1h. After 10 min centrifugation at 14000*g* at 4 °C, the supernatant was transferred to a new tube, dried under nitrogen, and re-suspended in 50 μL acetonitrile/water (75/25, v/v). 5 μL of each sample was combined to make a quality control (QC) metabolomics sample and ran every ten samples in the sequence to monitor for retention time and signal intensity drift. The remaining samples (30 μL) were transferred into injection vials for metabolomic analysis. Samples were randomized before analysis for all the LC-HRMS runs.

Metabolites were separated using an Ascentis^®^ Express HILIC HPLC column (2.1 mm x 150 mm, 2.7 µm particle size, #53946-U, SUPELCO Analytical) on an UltiMate 3000 quaternary UHPLC (Thermo Scientific, Waltham, MA) equipped with a refrigerated autosampler (4°C) and column heater (35°C). Solvent A (95% acetonitrile 10 mM ammonium acetate 0.1% acetic acid) and solvent B (5% acetonitrile with same modifiers as A) were used to elute the metabolites with a 26 min gradient, as follows: 10% B at 2 min, 30% B at 10 min, 100 % B at 15 min, 100 % B at 18 min, and back to 10 % B at 19 min. The flow rate was 0.25 ml/min. Samples were analyzed using a Q Exactive HF (QE-HF) (Thermo Scientific, Waltham, MA) equipped with a heated electro-spray ionization (HESI) source operated in both positive and negative ion mode. To build up metabolite spectral library for targeted analysis, we ran a plasma pooled sample extracted in 10x technical replicates in full scan/ddMS2 mode (untargeted metabolomics). The Full Scan settings were as follows: 120,000 resolution, AGC target, 1e6; Maximum IT, 200 ms; scan range, 60 to 900 m/z. Top 20 MS/MS spectral (dd-MS2) @ 15000 were generated with AGC target = 2e5, Maximum IT=25 ms, and (N)CE/stepped NCE = 20, 30, 40v. Metabolites detection and identification were performed using Compound Discovery 2.1 (Thermo Scientific, Waltham, MA) by searching against online database (mzCloud) and in-house database built on Sigma metabolomics library (LSMLS, Sigma-Aldrich) (mzVault). Well annotated metabolites (by a combination of high-resolution m/z ± 5 ppm, natural isotope abundances, MS2 spectra and retention time similarity with authentic standard or a similar chemistry standard^57^ were used to generate an inclusion list (Supplementary Table 3) for targeted PRM analysis. Final data acquisition was performed in Full scan and PRM modes. The PRM settings were as follows: resolution = 15000, AGC target = 2e5, Maximum IT=25 ms, loop count = 20, isolation window = 2.0, (N)CE was optimized for each metabolite.

### Lipidomics

50 μL plasma, spiked with 20 μL internal standard (Supplementary Table 2) was extracted with 1000 μL butanol/methanol (1/1, v/v, 10 mM ammonium formate) for 10 min on a shaker. Samples were pelleted by centrifugation at 4000g for 5 min at room temperature. The supernatant was moved to a clean glass tube, dried under nitrogen, and resuspended in 100 μL MTBE/methanol (1/3, v/v) with 10 mM ammonium formate. 5 μL of each sample was combined to make the QC-lipidomics sample that ran every ten samples in the mass spec sequence to monitor for retention time and signal intensity drift. The remaining samples were transferred into injection vials for lipidomic analysis. Lipids were analyzed exactly as we described before for plasma lipdiomics ^58^.

### Proteomics

5 μL plasma, was denatured in urea solution (8M), reduced with DTT (5 mM) at 37 °C for 30 min, alkylated with IAA (14 mM) for 30 min (in the dark), diluted with 100 mM ABC solution, and trypsin (1:50) digested at 37 °C overnight. The digestion was quenched by adding 10% formic acid. The digestion mixture was cleaned-up using Oasis HLB 1 cc (30 mg) SPE cartridges (WAT094225, Waters). The cartridges were activated with 500 μL solvent B (0.1% FA in 80% acetonitrile) x 3, equilibrated with 500 μL solvent A (0.1% FA in water) x 3, and after sample loading, washed with 750 μL solvent A x 4, eluted in 500 μL B x3, dried under nitrogen, and redissolved in 60 μL water with 0.1% FA. 5 μL of each sample was combined to make a QC-proteomics sample that ran every ten samples in the mass spec sequence to monitor for retention time and signal intensity drift. The remaining samples were transferred into injection vials for proteomics analysis.

Peptides were separated using a nanoACQUIY Peptide BEH C18 (130 Å, 1.7 μm, 150 μm x 100 mm) on a nanoAcquity system (Milford, MA, USA). Solvent A (99.5% water/0.5% acetonirtrile with 0.1% formic acid) and Solvent B (98 % acetonitrile/ 2% water with 0.1% formic acid) were used to elute the peptides with a 30 min gradient. The initial composition of 95% A was maintained for 3 min, and ramped to 70% A in 30 min, and washed at 2% A for 10 min and equilibrated at 95% A for 10 min. The flow rate was set at 2 μL/min. Samples were analyzed using a Q Exactive HF (QE-HF) (Thermo Scientific, Waltham, MA) equipped with a Michrom BioResources (AUBURN, CA, USA) source operated in positive ion mode. For DDA library generation (untargeted proteomics), data acquisition was performed on the pooled QC samples in Full Scan/ddMS2 mode @ 120,000 resolutions. The Full Scan settings as follows: 120,000 reolution, AGC target = 3e6; Maximum IT = 250 ms; scan range = 300 to 1650 m/z. Top 20 MS/MS spectral (dd-MS2) @ 15000 were generated with AGC target = 1e5, Maximum IT=25 ms, and (N)CE = 27 v. For untargeted proteomics analysis, ProteomeDiscover 4.2 (Thermo Scientific, Waltham, MA) was used for protein and peptide annotation. Well-annotated peptides (MS spectral match) were used to generate a inclusion list (Supplementary Table 5) for targeted proteomics analysis. Targeted proteomics were performed in full scan and PRM modes. The PRM settings were as follows: resolution = 15000, AGC target = 2e5, Maximum IT=25 ms, loop count = 32, isolation window = 2.0, (N)CE was set at 27 v.

### Data analysis

Peak detection and integration were performed using Skyline for metabolomics, lipidomics, and proteomics. For metabolites and lipids, the representative MS2 fragmentation was used for final quantification, and for peptides, the Top 3 MS2 fragmentations were summed to represent the final intensity. The missing values were replaced by LoDs (1/5 of the minimum positive value of each variable. Then data was log-transformed and normalized. For each analyte and each batch, a combination of internal standards and QC-locally estimated scatterplot smoothing (LOESS) was used for signal drift correction. We applied principal component analysis (PCA) to visualize the overall distribution of all samples (mean-centered and z-score scaled data) and exclude potential outliers using package FactoMineR and factoextra in R (https://www.r-project.org/).

To identify significantly altered metabolites or proteins, we firstly performed a two-tailed *t*-test analysis (false discovery rate, FDR<0.05), volcano plots of statistical significance against log2(fold change) were used to visualize the differences. Then a multivariate statistical analysis parameter-variable importance in projection (VIP) based on partial least square discrimination analysis (PLS-DA) were used to validate the statistically different features (metabolites, lipids, or proteins). For multiple group comparison in cell experiments, a one-way Analysis of variance (ANOVA) with Fisher’s LSD post hoc test was used to compute the significance. *P* < 0.05 *, *P* < 0.01 **, *P* < 0.001 ***. The data processing and visualization were performed using R package tidyverse, pheatmap, ggplot2, Hmisc, corrplot, ggsci, RColorBrewer, and pROC, and Graphpad Prism 9.0.

### Metabolites and lipids set enrichment analysis (MSEA)

Metabolite (and we included the lipids too) set enrichment analysis (MSEA) was performed on statistically different metabolites (P < 0.05 and FDR < 0.1) using MetaboAnalyst 5.0 (https://www.metaboanalyst.ca/MetaboAnalyst/). MSEA used 84 metabolite sets based on KEGG human metabolic pathways, and LSEA used 1072 sub chemical class metabolite (lipids) sets. Enrichment Ratio was computed by Hits / Expected.

### Metabolite correlation network

A debiased Sparse Partial Correlation (DSPC) algorithm was used to compute the correlation between all the quantified metabolites or significantly changed metabolites in Correlation Calculator 1.0.1.^59^ The MetScape 3.1.3 within Cytoscape 3.4.0 was used for the visualization of metabolite correlation network. Each node represents a compound, and each edge represents the partial correlation coefficient between two nodes.

### Pathway activity scoring

Pathway activity was assessed by metabolic pathway based PCA scores according to previous methods^60, 61^. Briefly, our quantified metabolite or lipids were mapped onto KEGG human metabolic pathways. For each pathway, intermediates were used to perform PCA, and the PC1 scores, which capture the maximum metabolite variances, were used as proxy of respective pathway activity.

### Machine Learning for FRDA prediction

26 metabolites that were altered in both the discovery set, and the validation set were used for machine learning based FRDA prediction. We applied linear support vector machine (SVM) to evaluate the feature importance by Monte-Carlo cross validation (MCCV) based sub-sampling in MetaboAnalyst 5.0. The procedure was repeated multiple times to select the important features to build SVM models and a 5-fold cross validation was performed to optimize the best model using Caret package in R. We also applied k-means clustering (KM) to detect features with similar behavior to reduce the redundancy. The final fitted model was applied to the validation set for prediction and verification.

## Data Availability

All data are available at the NIH Common Fund's National Metabolomics Data Repository (NMDR) website (supported by NIH grant U2C-DK119886), the Metabolomics Workbench, https://www.metabolomicsworkbench.org where it has been assigned Project ID PR001432. The data can be accessed directly via it's Project DOI:10.21228/M8MQ5M. Further information and requests for resources and reagents should be directed to and will be fulfilled by the lead contact, Clementina Mesaros.

## Acknowledgements

We thank Prof. Ian Blair for providing unlimited access to the use of LC-HRMS, Rachel Truitt for assistance with the iPSC-CMs, and Dr. Nathaniel Snyder for discussions and editing of the manuscript. We are very grateful to all FRDA patients for agreeing to give blood for research purposes and to the healthy volunteers that agreed to serve as controls. We recognize the positive support of the Hamilton and Finneran families and the Penn Medicine/CHOP Friedreich’s Ataxia Center of Excellence.

## Funding

National Institutes of Health grant R21NS116315 (CM)

National Institutes of Health grant P30ES013508 (CM)

Friedreich’s Ataxia Research Alliance (FARA) Postdoctoral Research Award (DW)

## Author contributions

Conceptualization: CM, DW, MGC

Methodology: DW, MGC, BEH, DRL

Investigation: DW, MGC, CM

Visualization: DW, BEH

Funding acquisition: CM, DW

Project administration: CM, DRL

Supervision: CM

Writing original draft: DW, CM, DRL

Writing review & editing: DW, MGC, CM, BEH, DRL

## Competing interests

Authors declare that they have no competing interests.

## Supplementary material

**Supplementary Fig. 1.**
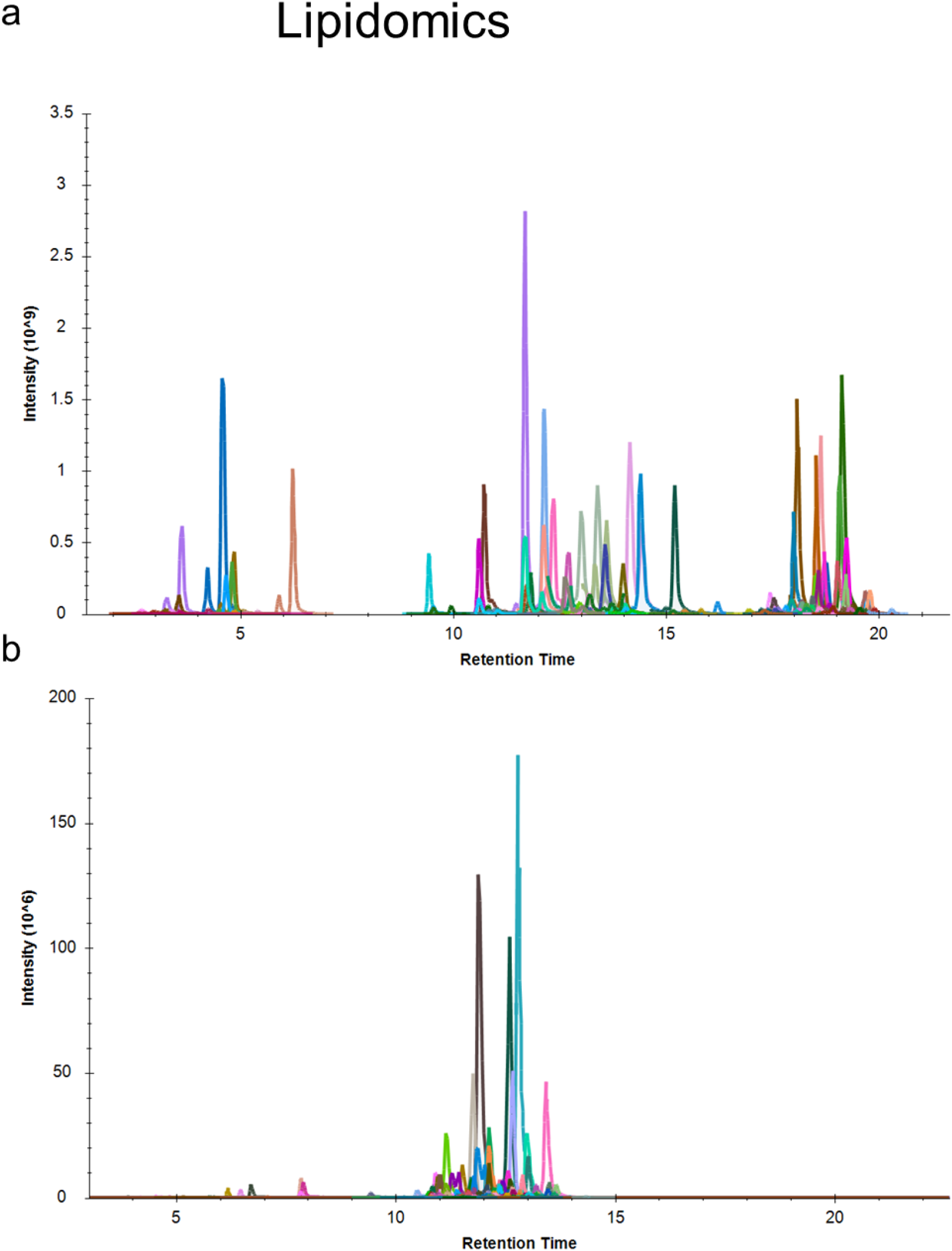
The extracted ion chromatogram (XIC) of lipids in positive ion mode (a) and in negative ion mode (b).

**Supplementary Fig. 2.**
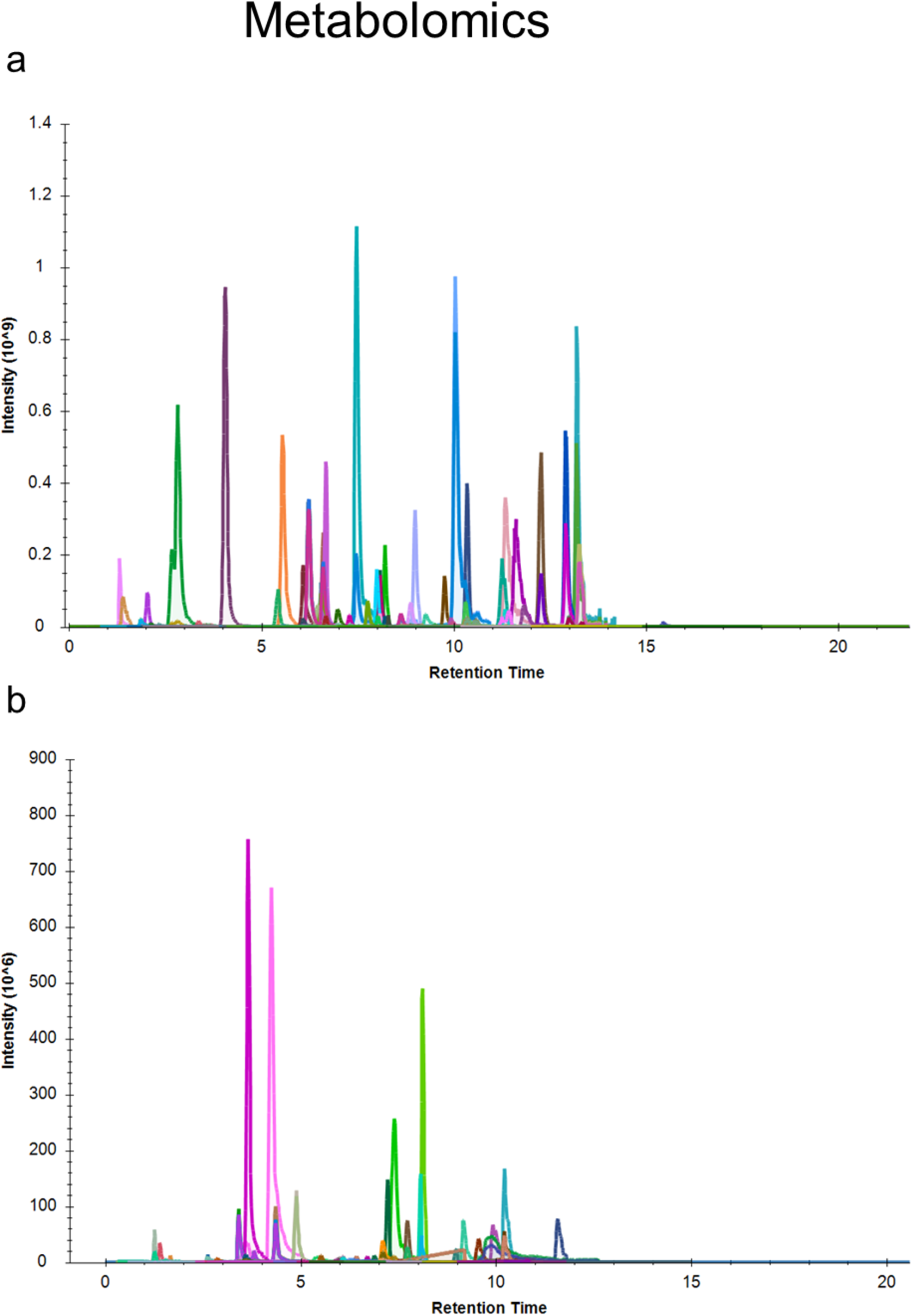
The extracted ion chromatogram (XIC) of metabolites in positive ion mode (a) and in negative ion mode (b).

**Supplementary Fig. 3.**
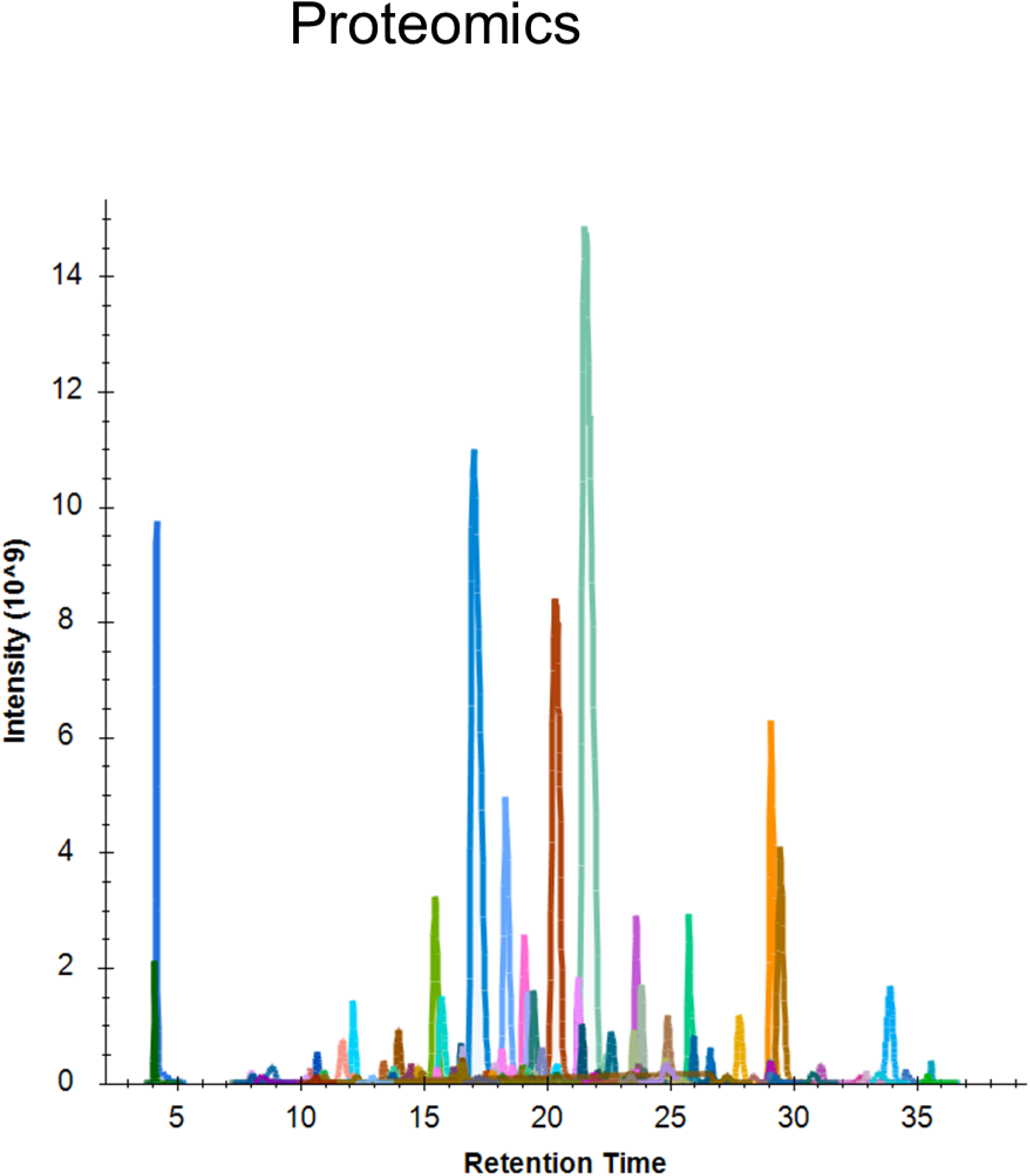
The extracted ion chromatogram (XIC) of protein (peptides) in positive ion mode.

**Supplementary Fig. 4.**
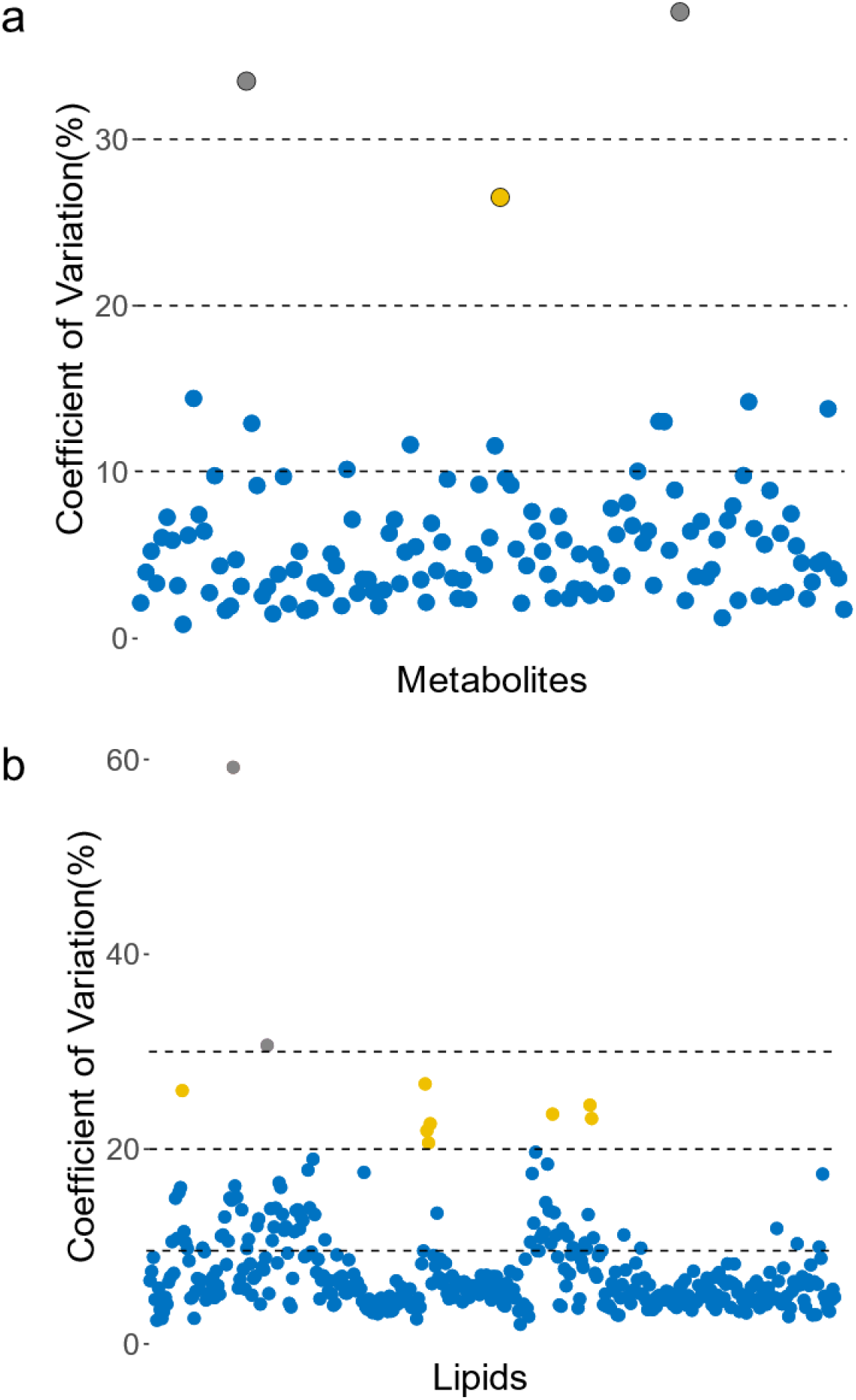
Coefficient of variances (CVs) of quantified metabolites (a) and lipids (b) in each of their quality control (QC) samples.

**Supplementary Fig. 5.**
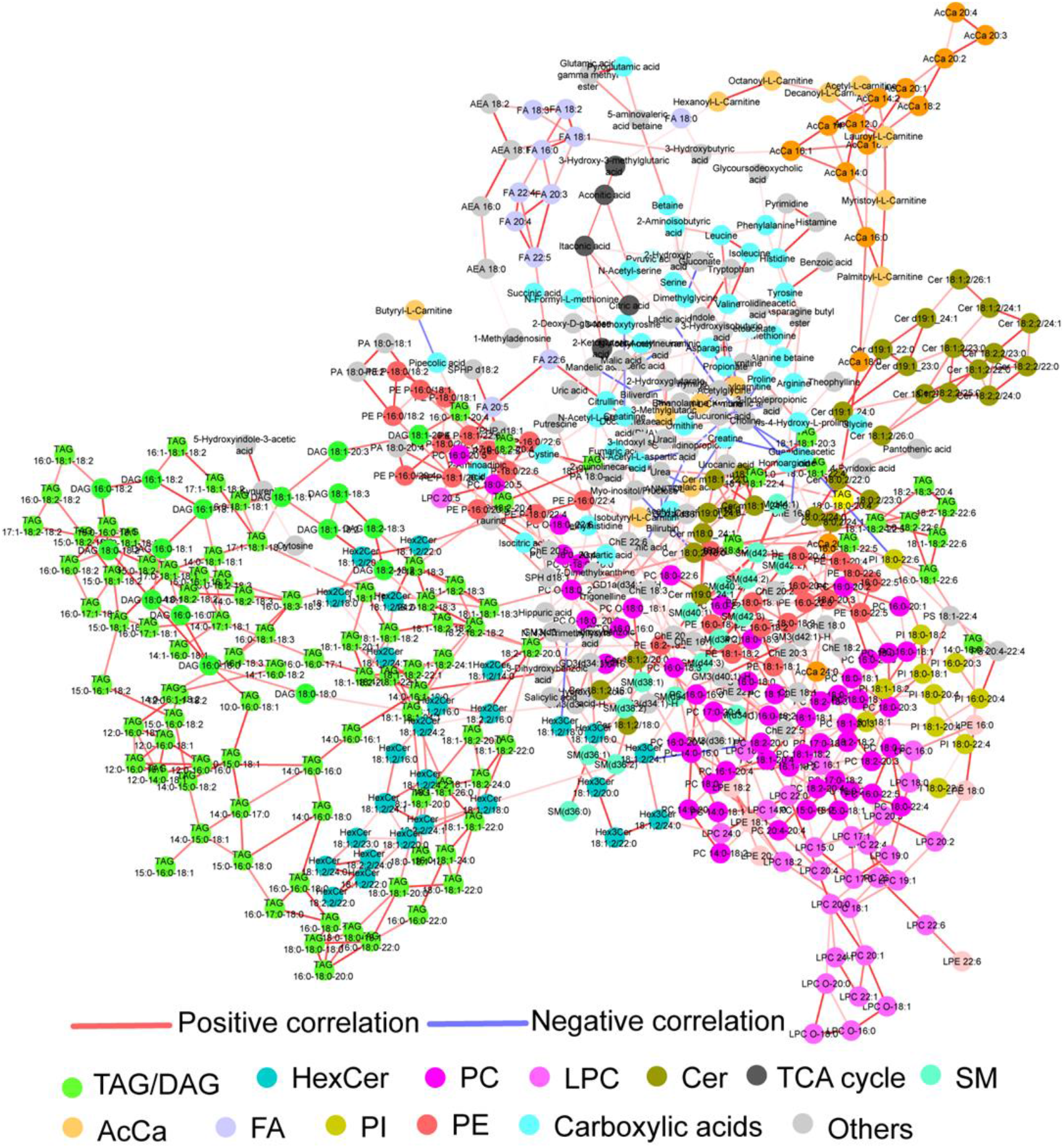
Metabolite correlation network based on all of metabolites and lipids. Each node represented one metabolite (lipid), and nodes were colored according to metabolite (lipid) class. The edge colored with red indicated positive correlation, while the edge with blue indicated negative correlation. The edge width represented the strength of partial correlation coefficient.

**Supplementary Fig. 6.**
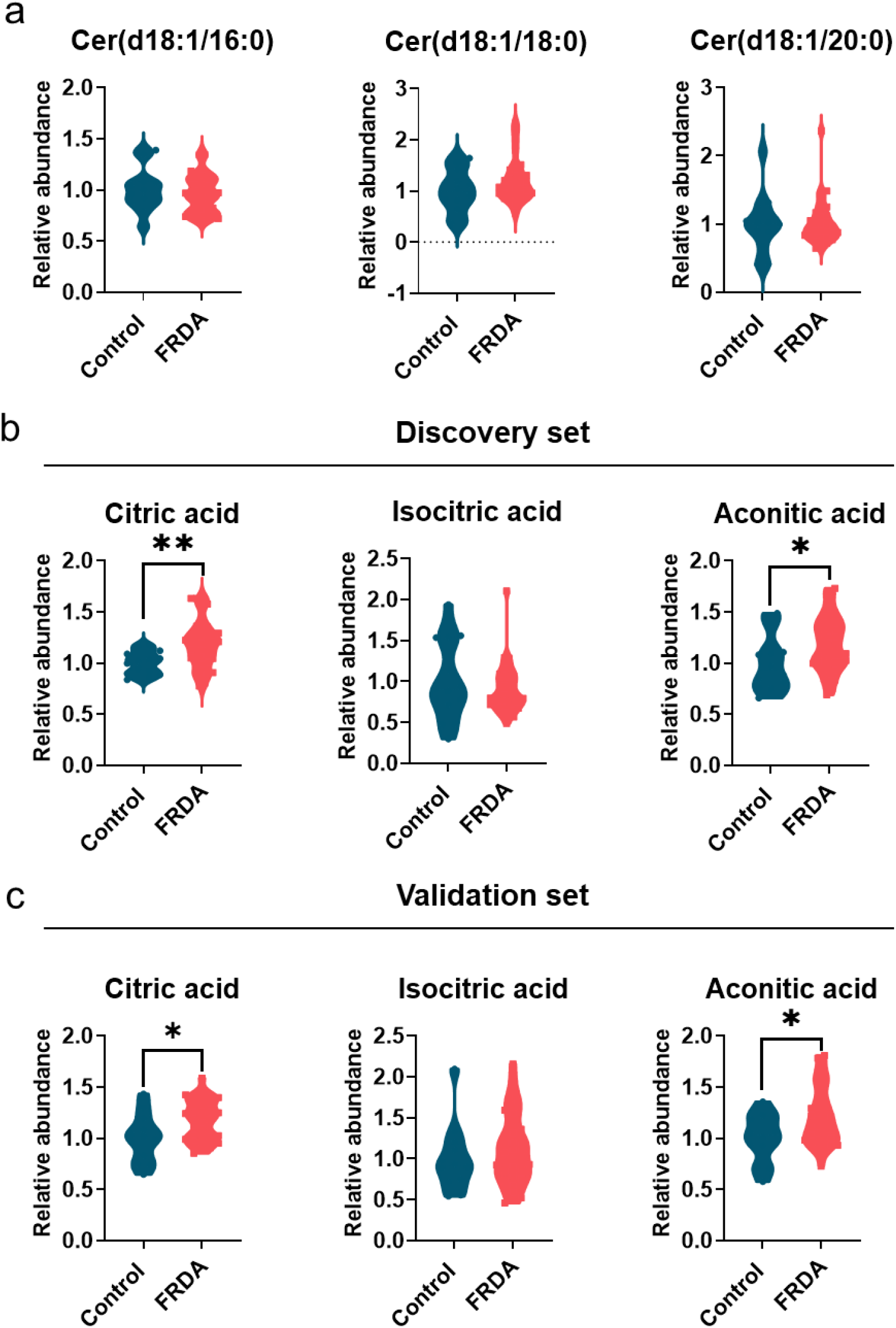
The relative abundance of long chain ceramides in validation set (a). Relative abundance of citric acid cycle intermediates between healthy controls and FRDA patients in discovery set (b) and validation set (c).

**Supplementary Fig. 7.**
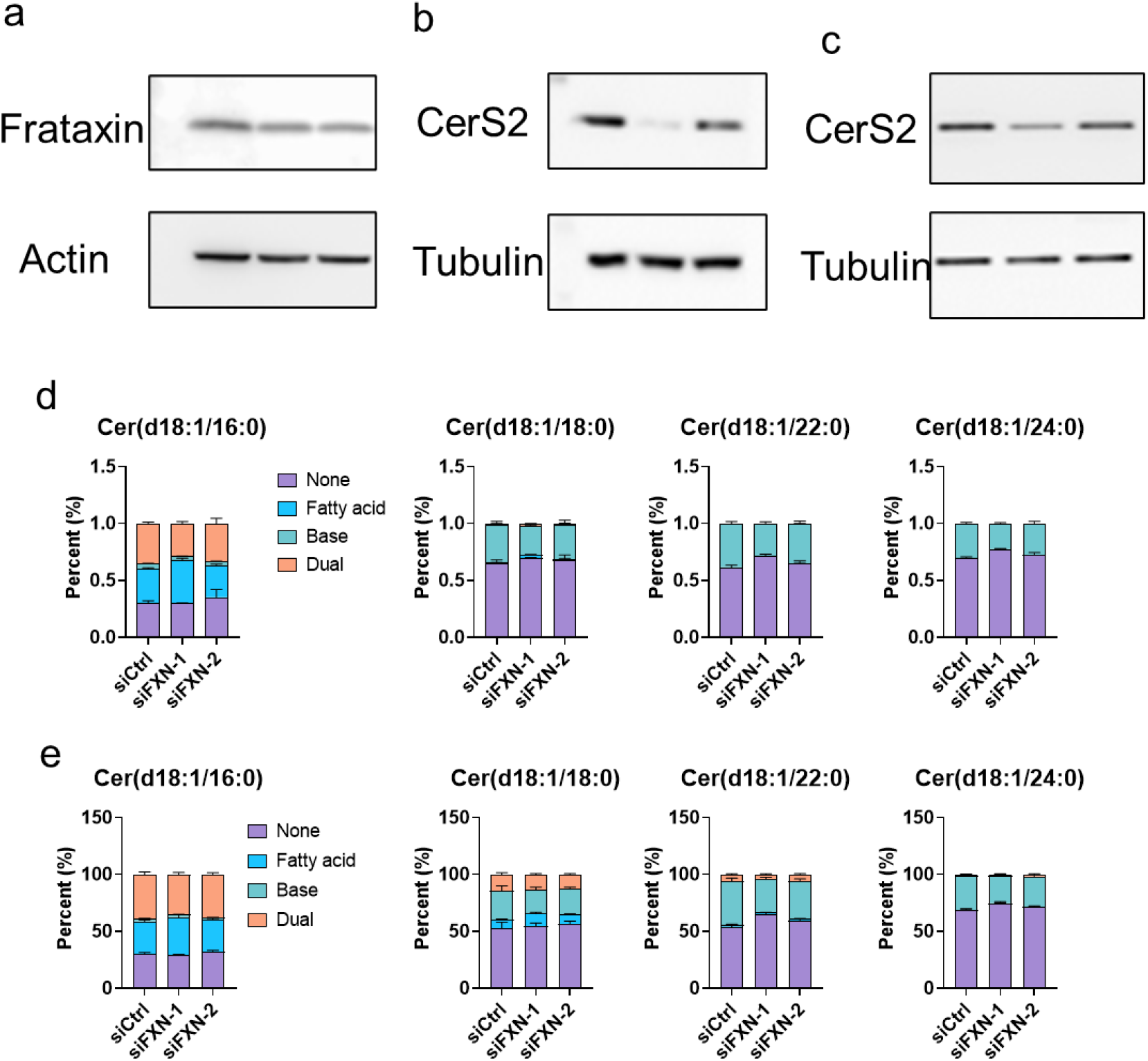
Unbalanced ceramide synthesis in frataxin-deficient cardiomyocytes. a Western blots showing the frataxin levels in FXN-knockdown cardiomyocyte 233-SeV4 b,c Western blots showing the protein levels of ceramide synthase (CerS2) in cardiomyocyte 11713 (b) and 233-SeV4 (c) after siRNA-mediated FXN knockdown doe The mass isotopomer distribution of ceramides from FXN-knockdown cardiomyocyte 11713 (d) and 233-SeV4 (e)

**Supplementary Fig. 8.**
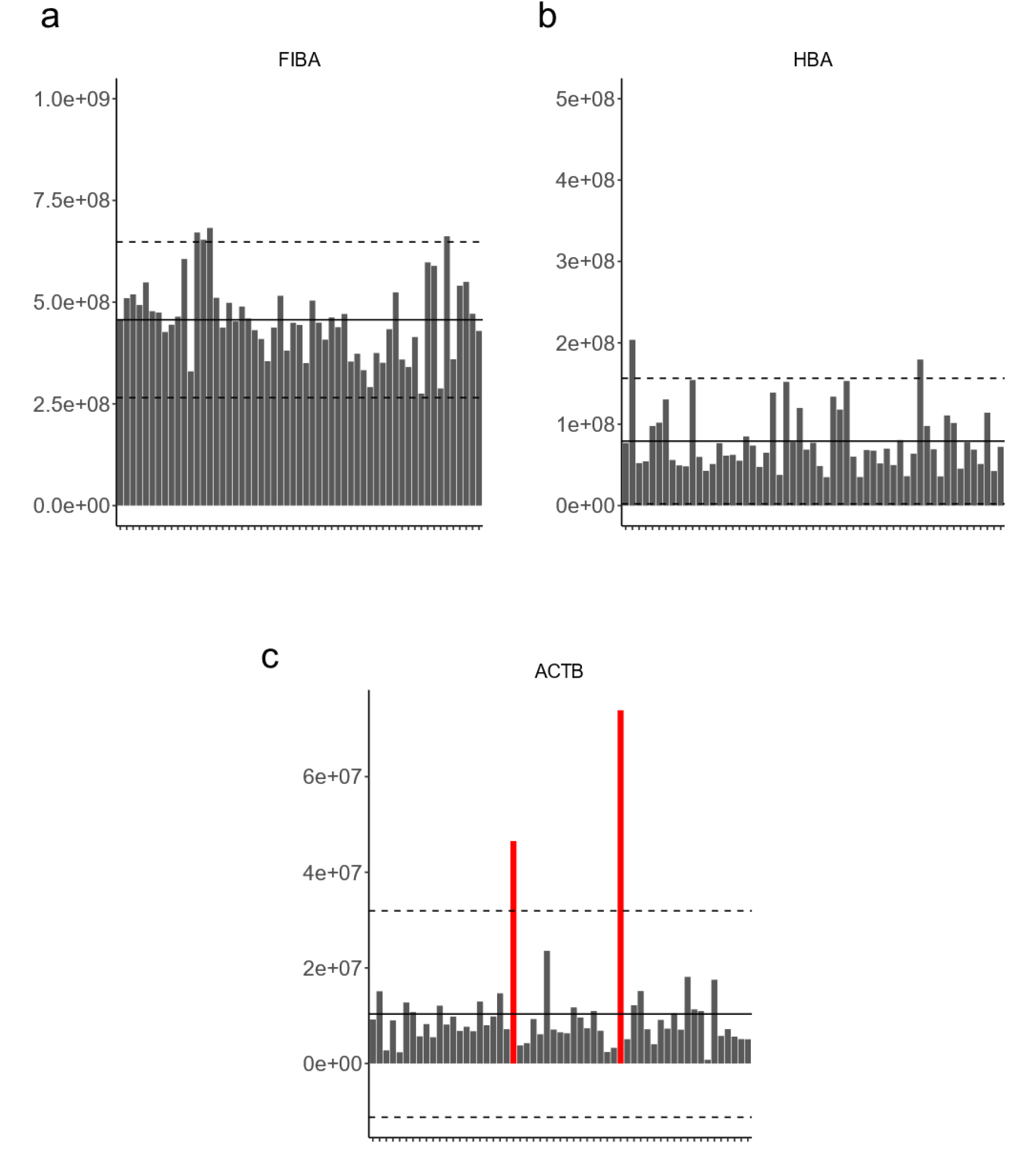
Quality assessment of each sample according to main contamination sources of coagulation (a), erythrocyte (b), and platelet (c).

**Supplementary Fig. 9.**
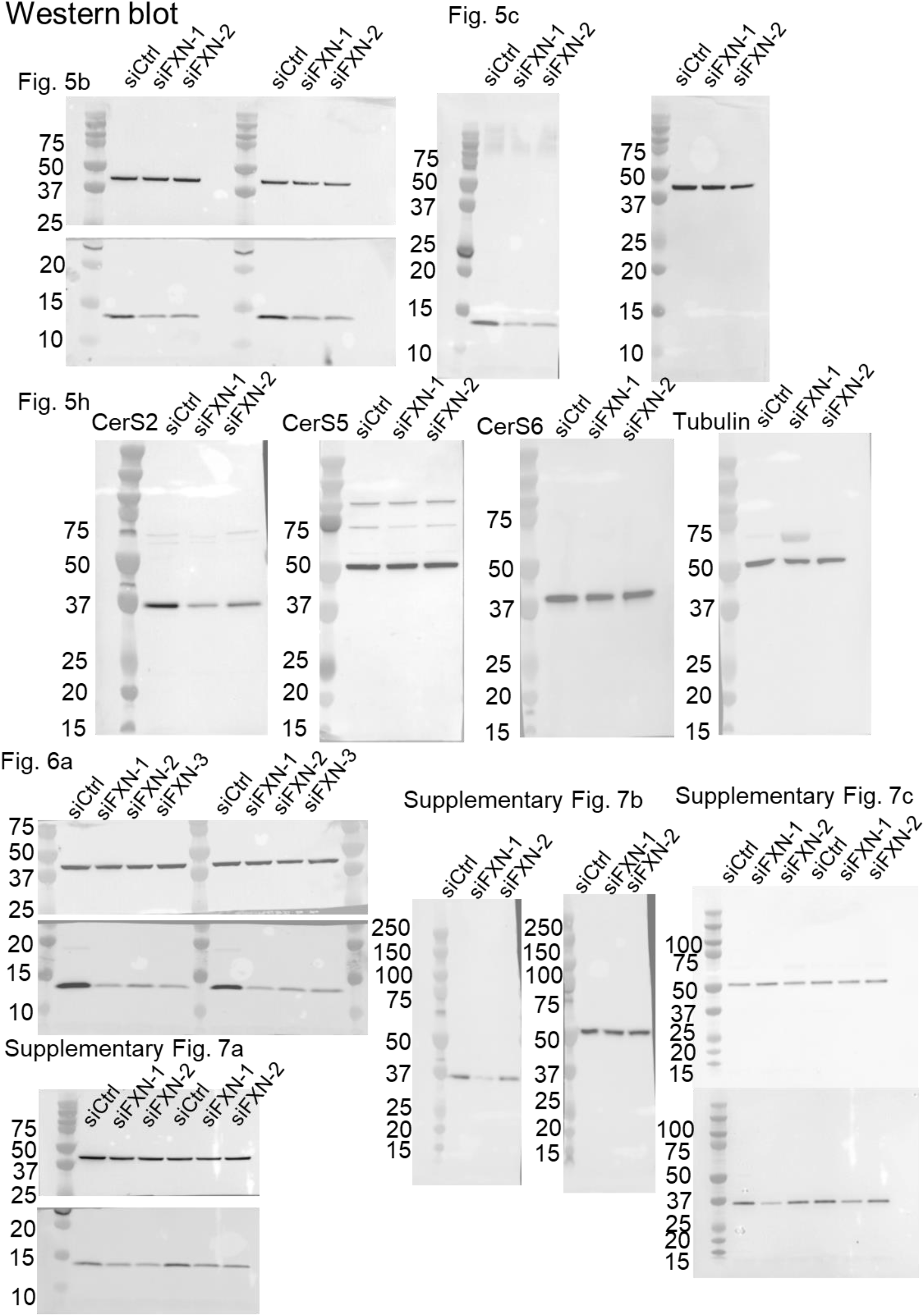
Full Western blots used in the cellular work.

**Supplementary Fig. 10.**
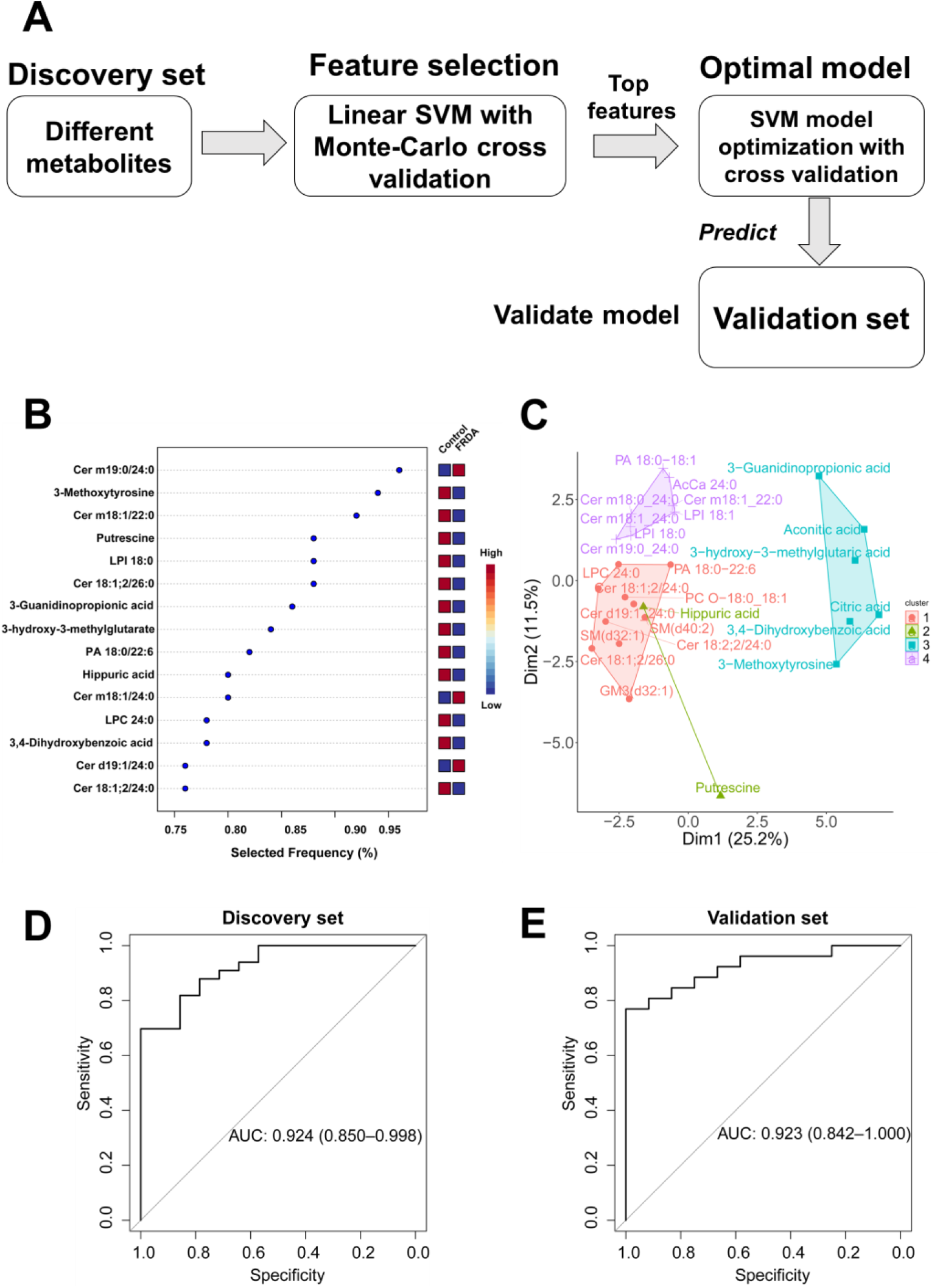
Machine learning improves the prediction of FRDA. **A.** Machine learning workflow for prediction of FRDA using metabolic features. **B.** Top 15 selected metabolites from Linear support vector machine (SVM) model. **C**. K-Means Clustering revealed different 4 clusters for these metabolites. **D.** The receiver operating characteristic (ROC) curves using three metabolites [Cer(m19:0/24:0), 3-methoxytyrosine, and putrescine] based SVM model in the discovery set **E**. same as D but in the validation set.

**Supplementary Table 1.** Demographic data for each cohort.

| **Discovery set (59)** |  |  |  | **Validation set (38)** |  |
| --- | --- | --- | --- | --- | --- |
|  | Control | FRDA | Carrier | Control | FRDA |
| **Gender** | M(8)/F(7) | M(10)/F(19)<br>4 Unknown | M(4)/F(7) | M(7)/F(5) | M(17)/F(9) |
| **Age(mean±sd)** | 32.7±11.0 | 25.6±8.1 | 54.2±9.3 | 29.4±7.6 | 21.2±7.7 |
| **GAA1(mean±sd)** | — | 634.9±184.2 | — | — | 693.3±198.2 |
| **Age of onset**<br>**(mean±sd)** | — | 11.8±5.4 | — | — | 11.6±4.2 |

**Supplementary Table 2.** Internal Standards (ISTD) used for metabolomics and lipidomics analysis.

| **Internal Standard (ISTD)** | **Concentration in Mix (ng $\mu\text{L}^{-1}$)** | **Supplier** |
| --- | --- | --- |
| **Metabolomics ISTD Mix** |  |  |
| Glycine (13C2, 99%; 15N, 99%) | 11 | CIL |
| Serine (13C3, 98%; 15N, 98%) | 60 | CIL |
| Isoleucine (13C6, 99%; 15N, 99%) | 10.5 | CIL |
| Threonine-13C4,15N | 11 | SA |
| Sodium Pyruvate (13C3, 99%) | 10 | CIL |
| Arginine (13C6, 99%) | 9.5 | CIL |
| Valine (13C5, 99%; 15N, 99%) | 110 | CIL |
| Tryptophan (13C11, 99%) | 11.5 | CIL |
| Lysine (13C6, 99%) | 10 | CIL |
| Aspartic Acid (13C4, 99%; 15N, 99%) | 10.5 | CIL |
| Glutamic Acid-13C5,15N | 10 | SA |
| Glutamine-13C5,15N2 | 10 | SA |
| D-3-Hydroxybutyrate (13C4, 99%) | 10.5 | CIL |
| Adenosine (Ribose-13c5, 98%+) | 50 | CIL |
| Alpha-Ketoglutaric Acid (13C5, 99%) | 2.5 | CIL |
| A-Hydroxyglutaric Acid-13C5 | 2.5 | SA |
| Fumaric Acid – [13]C4 | N/A | CIL |
| Succinic Acid-2,2,3,3-d4 | 11 | SA |
| Citric Acid-13C6 | 9 | SA |
| <sup>1</sup>Carnitine – D9 | 2.16 | CIL |
| <sup>1</sup>Acetylcarnitine – D3 | 0.77 | CIL |
| <sup>1</sup>Propionylcarnitine – D3 | 0.16 | CIL |
| <sup>1</sup>Butyrylcarnitine – D3 | 0.17 | CIL |
| <sup>1</sup>Isovalerylcarnitine – D9 | 0.18 | CIL |
| <sup>1</sup>Octanoylcarnitine – D3 | 0.21 | CIL |
| <sup>1</sup>Myristoylcarnitine – D9 | 0.26 | CIL |
| <sup>1</sup>Palmitoylcarnitine – D3 | 0.56 | CIL |
| **Lipidomics ISTD Mix** |  |  |
| <sup>2</sup>15:0-18:1 PC – D7 | 80 | APL |
| <sup>2</sup>15:0-18:1 PE – D7 | 2.5 | APL |
| <sup>2</sup>15:0-18:1 PS – D7 | 2.5 | APL |
| <sup>2</sup>15:0-18:1 PG – D7 | 15 | APL |
| <sup>2</sup>15:0-18:1 PI – D7 | 5 | APL |
| <sup>2</sup>15:0-18:1 PA – D7 | 3.5 | APL |
| <sup>2</sup>18:1 LPC – D7 | 12.5 | APL |
| <sup>2</sup>18:1 LPE – D7 | 2.5 | APL |
| <sup>2</sup>18:1 Cholesterol Ester – D7 | 175 | APL |
| <sup>2</sup>18:1 MG – D7 | 1 | APL |
| <sup>2</sup>15:0-18:1 DG – D7 | 5 | APL |
| <sup>2</sup>15:0-18:1-15:0 TG – D7 | 27.5 | APL |
| <sup>2</sup>18:1 SM – D9 | 15 | APL |
| <sup>2</sup>Cholesterol – D7 | 50 | APL |
| Sphingosine (C17 base) | 3.6 | APL |
| Sphinganine (C17 base) | 3.6 | APL |
| Sphingosine-1-P (C17 base) | 4.6 | APL |
| Sphinganine-1-P (C17 base) | 4.6 | APL |
| Lactosyl(β) C12 Ceramide | 8.1 | APL |
| 12:0 Sphingomyelin | 6 | APL |
| Glucosyl(β) C12 Ceramide | 8 | APL |
| 12:0 Ceramide | 10.1 | APL |
| 12:0 Ceramide-1-P | 7 | APL |
| 25:0 Ceramide | 8.3 | APL |
| Palmitoyl-1,2,3,4-13C4-L-carnitine hydrochloride | 2.2 | SA |

CIL = Cambridge Isotope Laboratories, Inc. (Tewksbury, MA, USA).
SA = Sigma-Aldrich (St. Louis, MO, USA)
TRC = Toronto Research Chemicals, Inc.,
APL = Avanti Polar Lipids (Birmingham, AL, USA). CC = Cayman Chemicals
<sup>1</sup> Labeled carnitine standards set B from CIL
<sup>2</sup> Splash Lipidomix Mass Spec Standards from APL

**Supplementary Table 3.**
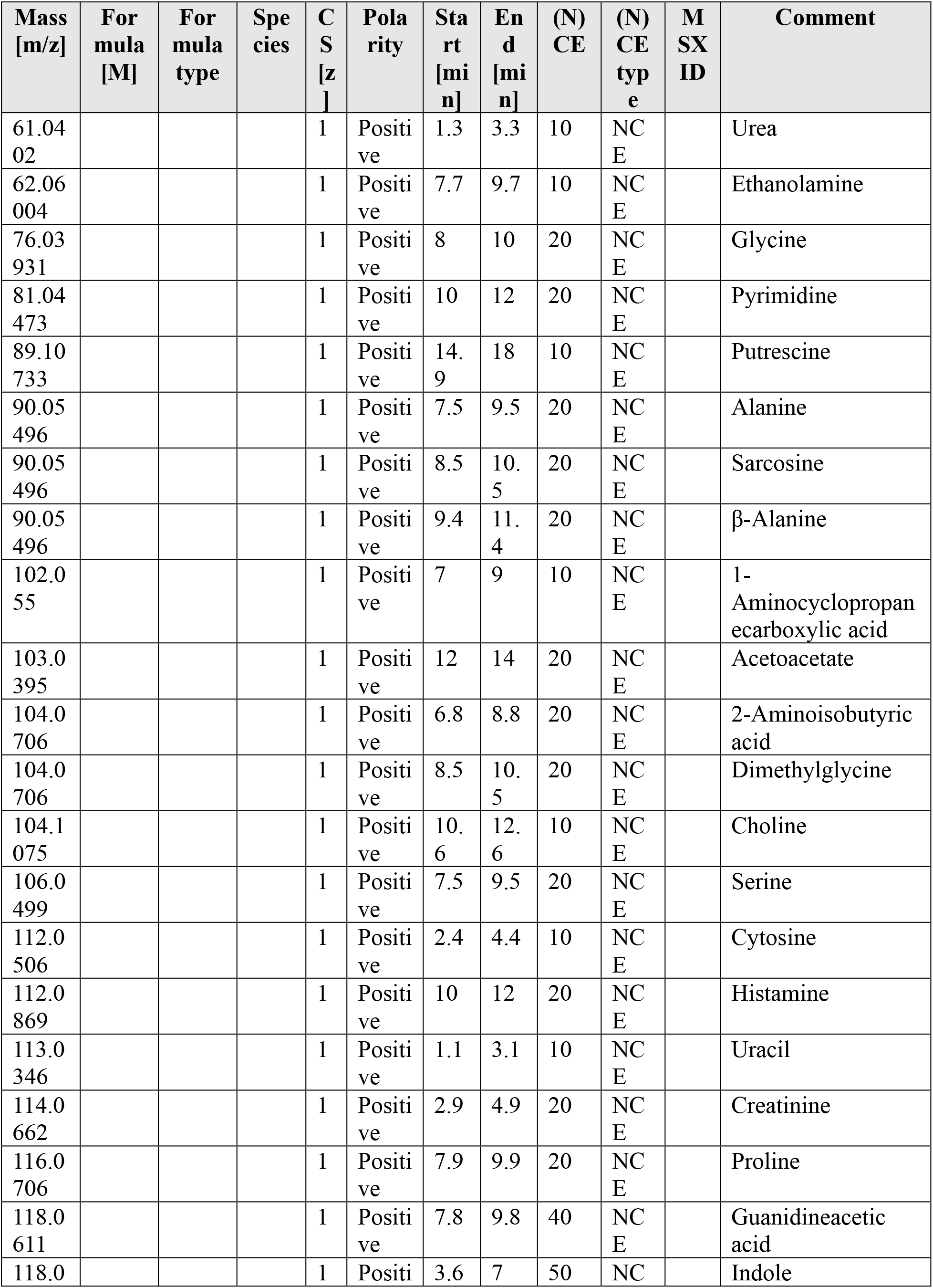
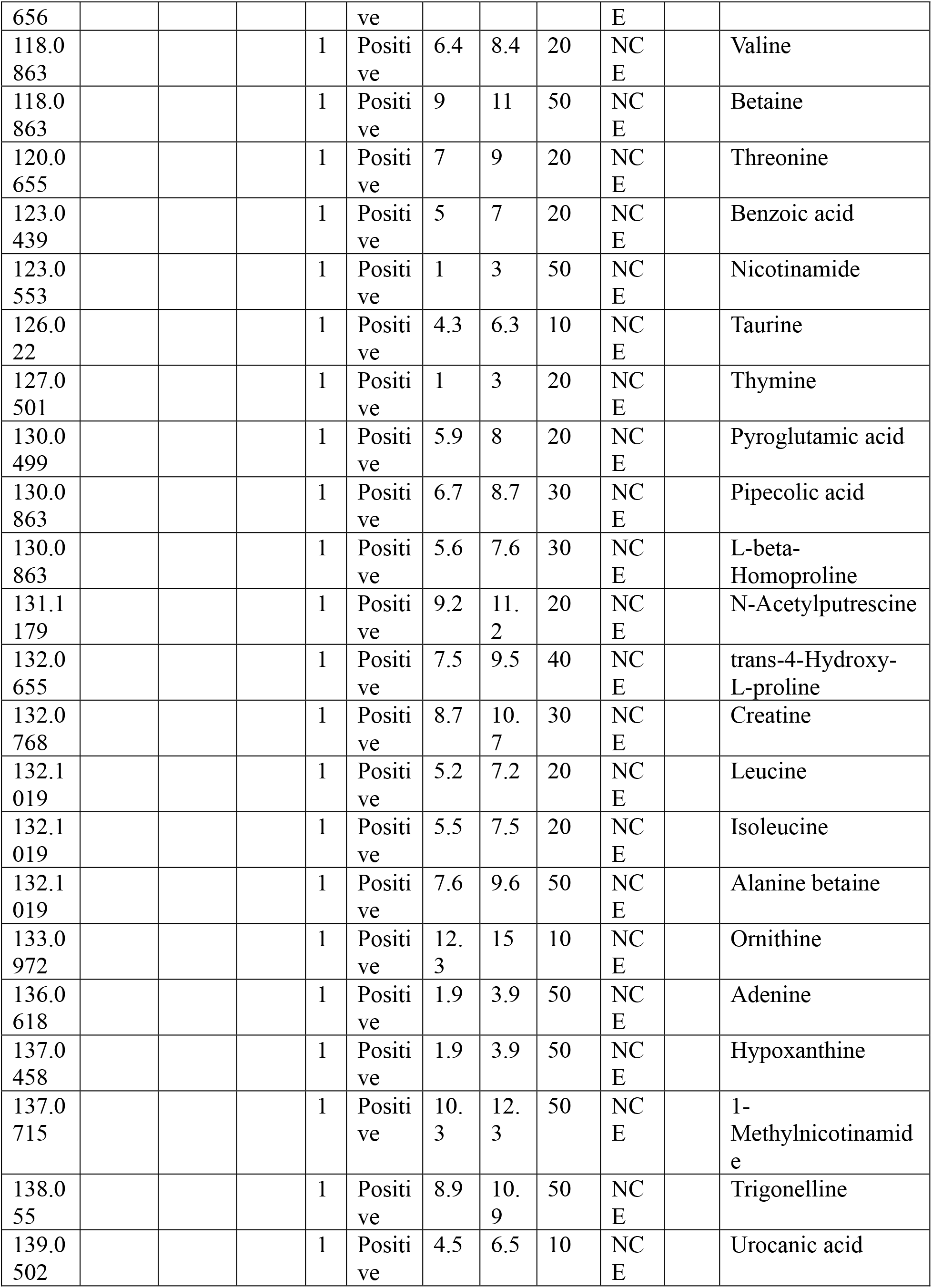
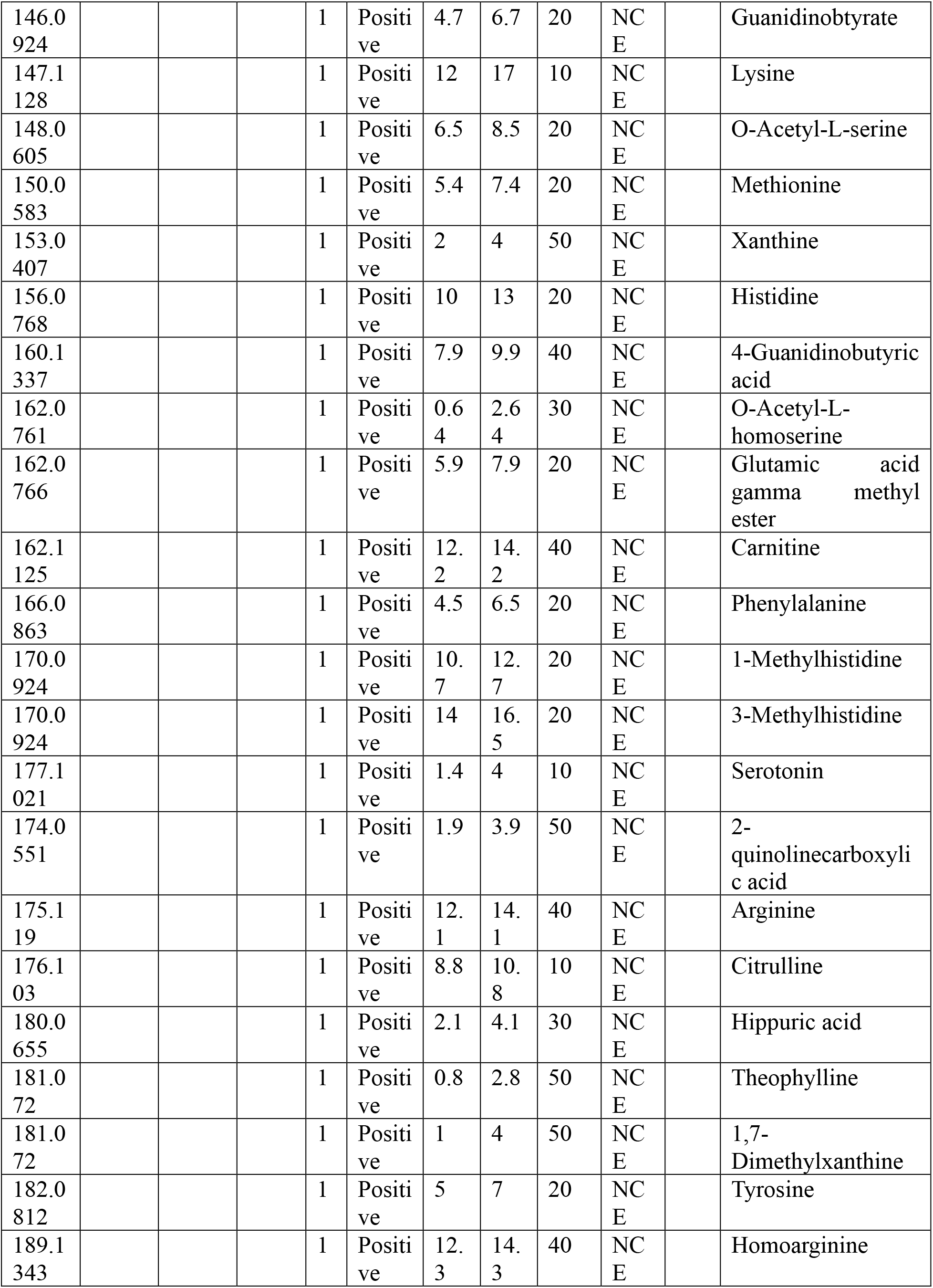
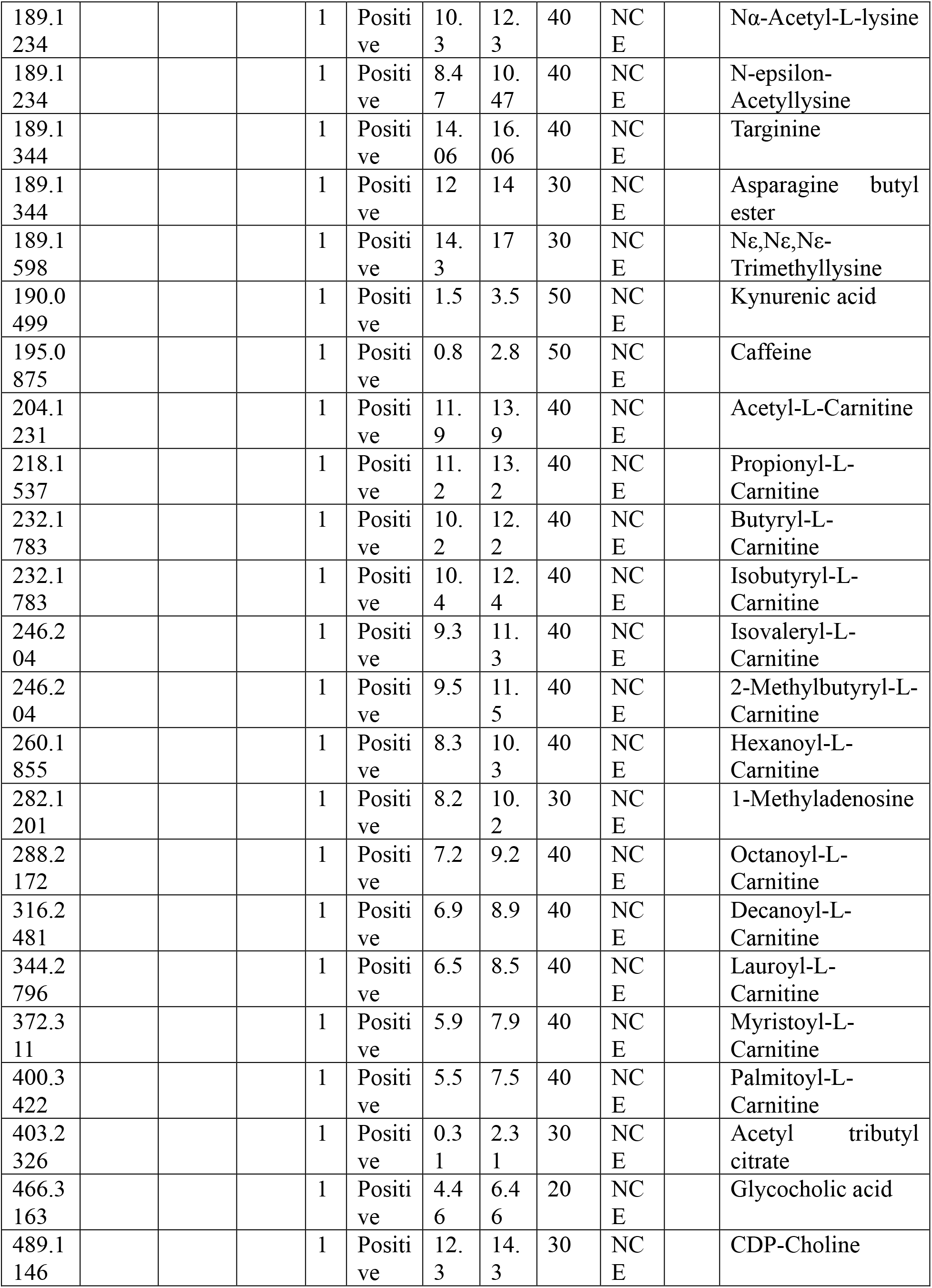
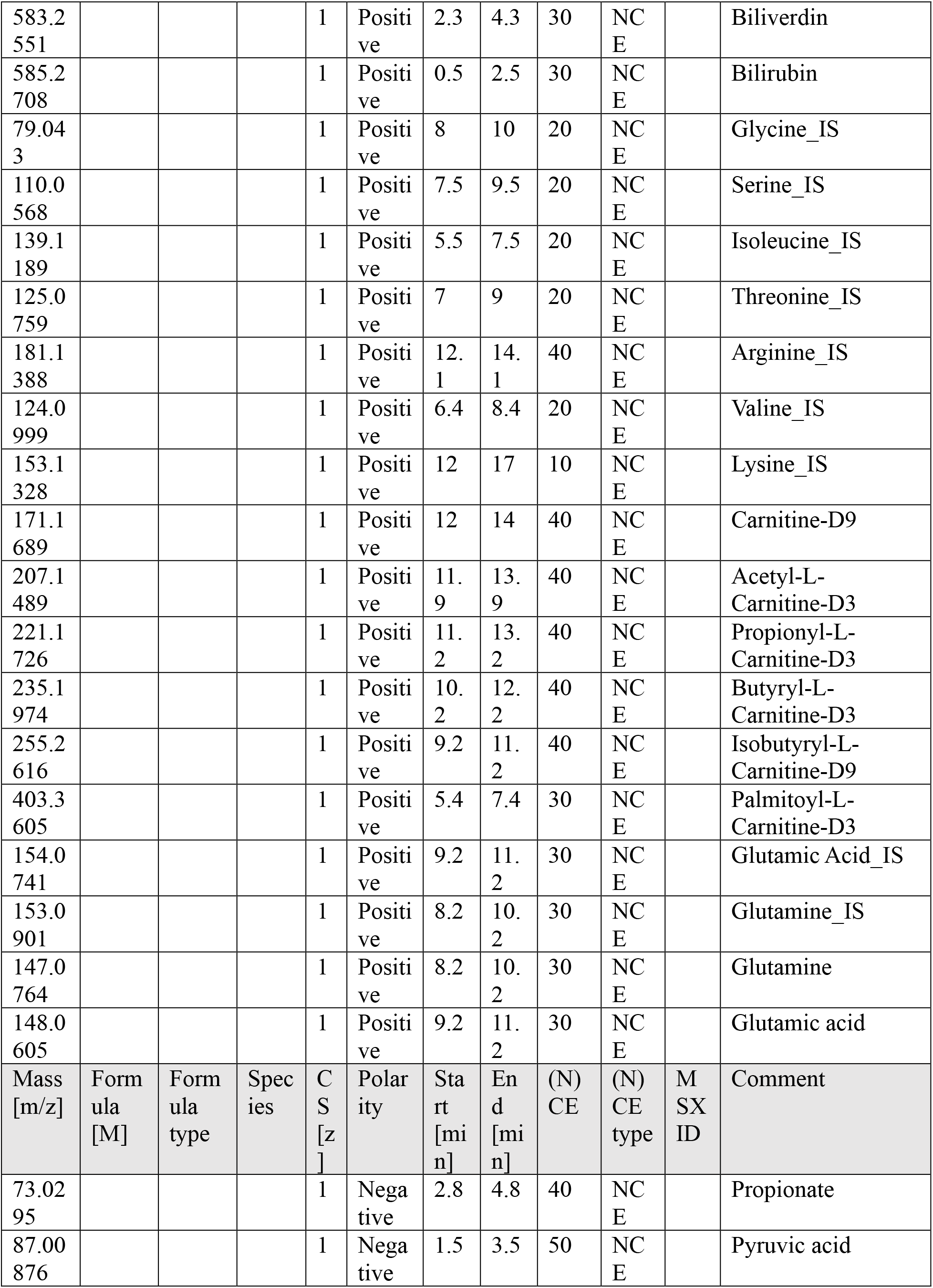
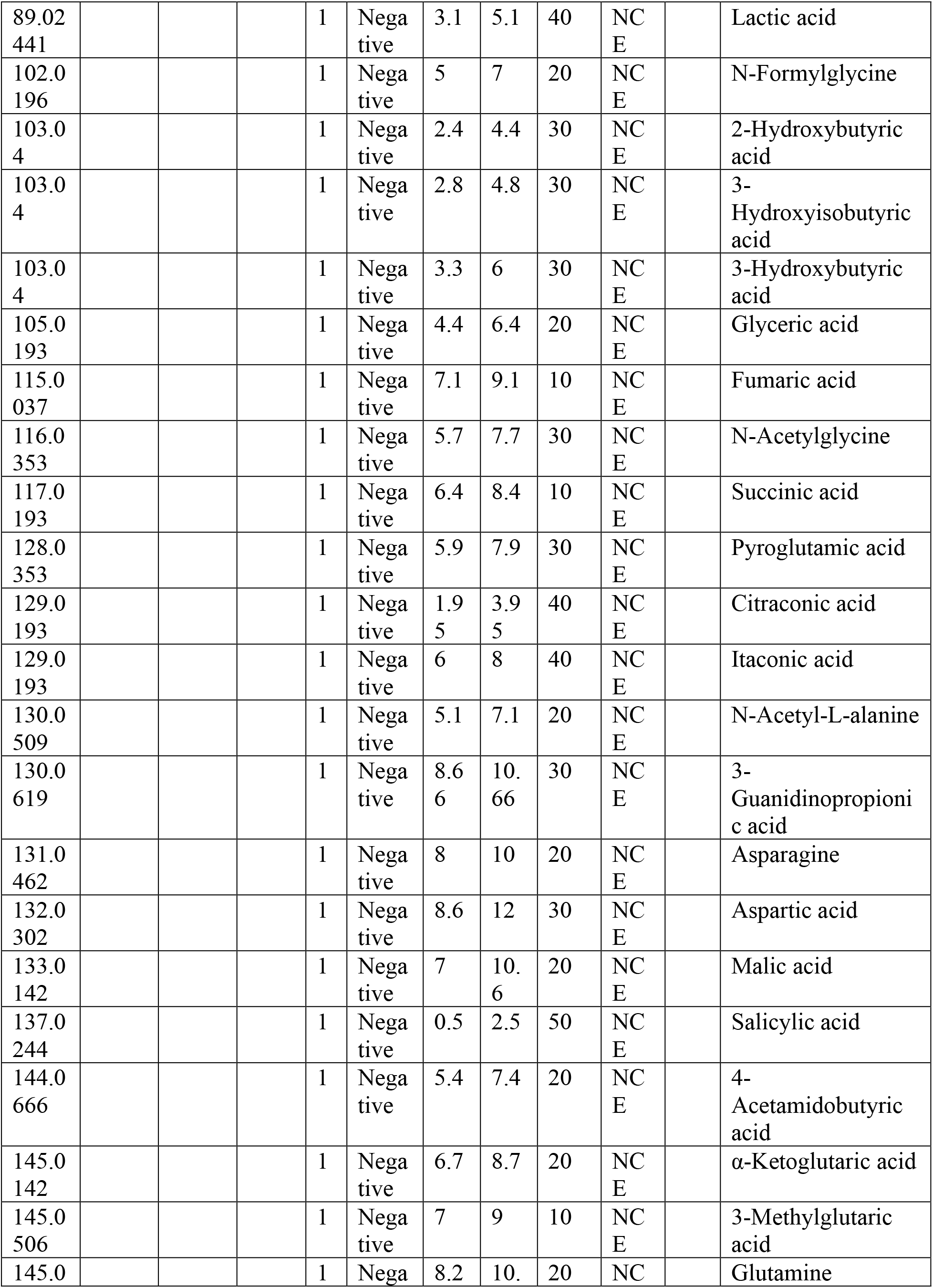
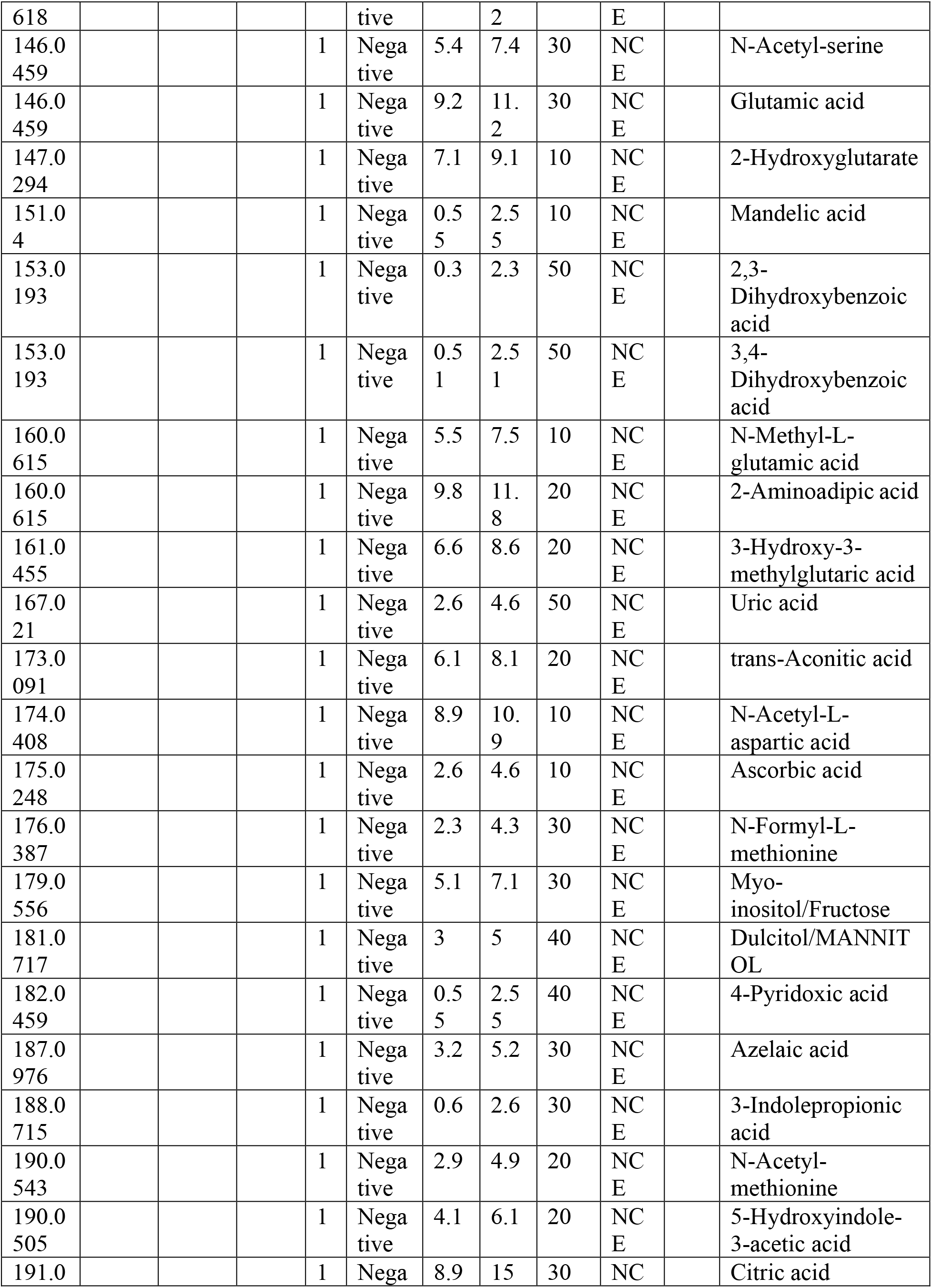
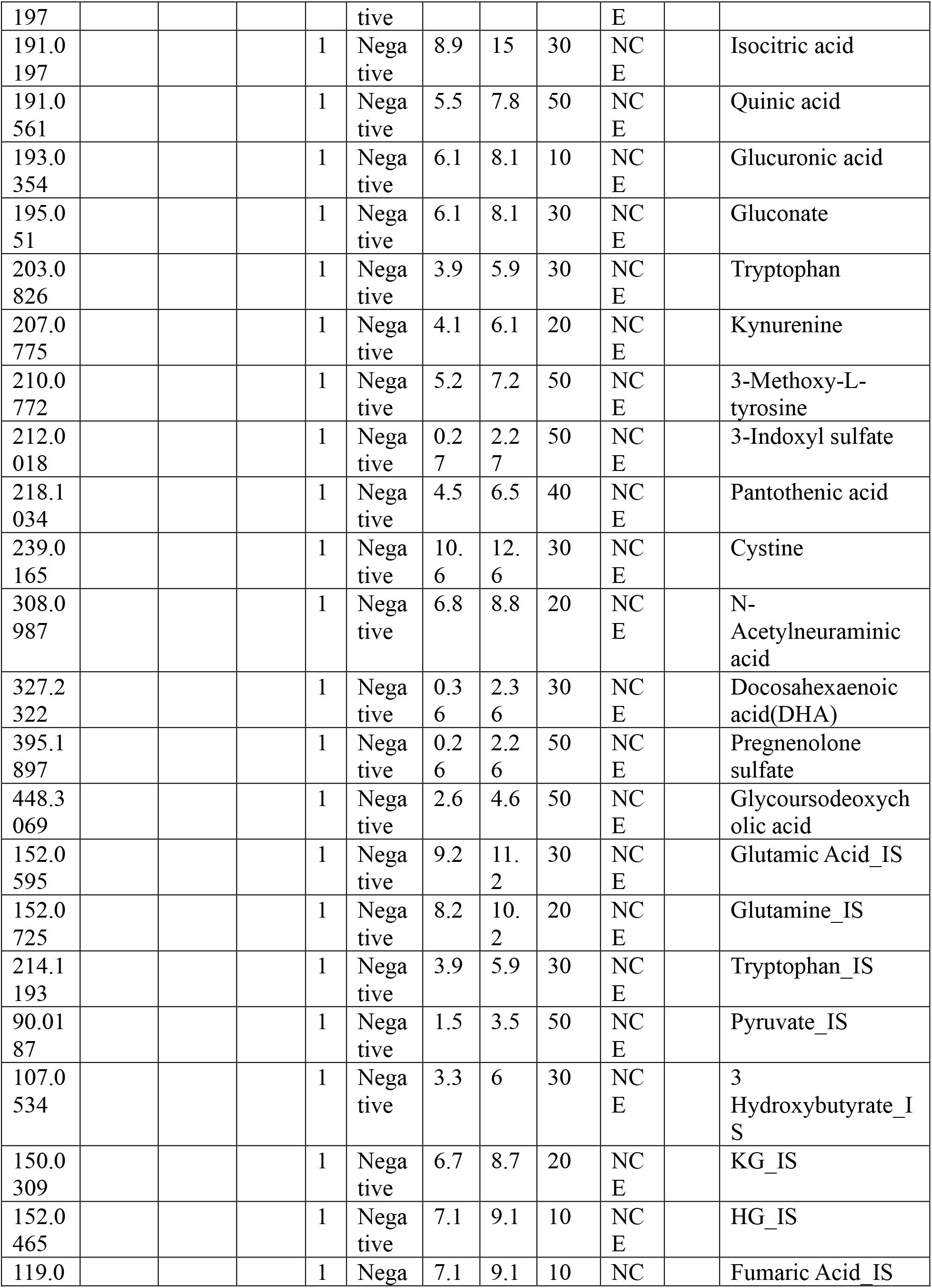
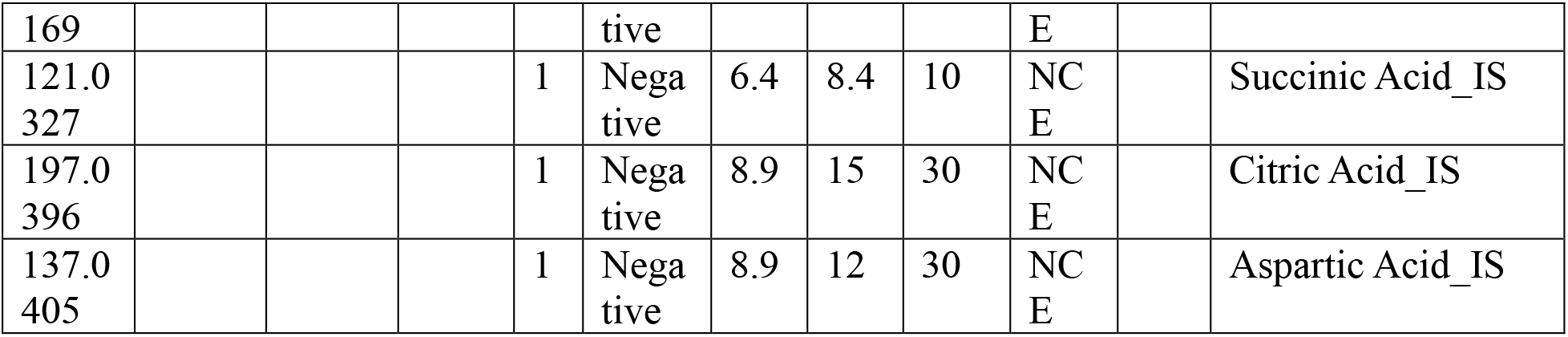
Inclusion list for targeted **metabolomics** (PRM).

**Supplementary Table 4.** Inclusion list for targeted lipidomics (PRM).

| Mass<br>[m/z] | Form<br>ula<br>[M] | Formu<br>la type | Spe<br>cies | CS<br>[z] | Polar<br>ity | Start<br>[min<br>] | End<br>[min<br>] | (N)<br>CE | (N)C<br>E<br>type | MS<br>X<br>ID | Comment |
| --- | --- | --- | --- | --- | --- | --- | --- | --- | --- | --- | --- |
| 344.2<br>795 |  |  |  | 1 | Posit<br>ive | 1 | 2 | 30 | NCE |  | AcCa 12:0 |
| 372.3<br>108 |  |  |  | 1 | Posit<br>ive | 2.5 | 3.5 | 30 | NCE |  | AcCa 14:0 |
| 370.2<br>952 |  |  |  | 1 | Posit<br>ive | 1.4 | 2.4 | 30 | NCE |  | AcCa 14:1 |
| 368.2<br>795 |  |  |  | 1 | Posit<br>ive | 1 | 2 | 30 | NCE |  | AcCa 14:2 |
| 400.3<br>421 |  |  |  | 1 | Posit<br>ive | 4.1 | 6 | 30 | NCE |  | AcCa 16:0 |
| 404.3<br>552 |  |  |  | 1 | Posit<br>ive | 4.1 | 6 | 30 | NCE |  | AcCa 16:0-13C4 |
| 403.3<br>606 |  |  |  | 1 | Posit<br>ive | 4.1 | 6 | 30 | NCE |  | AcCa 16:0-D3 |
| 398.3<br>265 |  |  |  | 1 | Posit<br>ive | 2.9 | 4.5 | 30 | NCE |  | AcCa 16:1 |
| 428.3<br>734 |  |  |  | 1 | Posit<br>ive | 5.4 | 7.5 | 30 | NCE |  | AcCa 18:0 |
| 426.3<br>578 |  |  |  | 1 | Posit<br>ive | 4.3 | 6 | 30 | NCE |  | AcCa 18:1 |
| 424.3<br>421 |  |  |  | 1 | Posit<br>ive | 3.4 | 5 | 30 | NCE |  | AcCa 18:2 |
| 456.4<br>047 |  |  |  | 1 | Posit<br>ive | 6.9 | 7.9 | 30 | NCE |  | AcCa 20:0 |
| 454.3<br>891 |  |  |  | 1 | Posit<br>ive | 5.5 | 6.5 | 30 | NCE |  | AcCa 20:1 |
| 452.3<br>734 |  |  |  | 1 | Posit<br>ive | 4.6 | 5.6 | 30 | NCE |  | AcCa 20:2 |
| 450.3<br>578 |  |  |  | 1 | Posit<br>ive | 3.9 | 4.9 | 30 | NCE |  | AcCa 20:3 |
| 448.3<br>421 |  |  |  | 1 | Posit<br>ive | 3 | 4.3 | 30 | NCE |  | AcCa 20:4 |
| 484.4 |  |  |  | 1 | Posit | 8.3 | 9.3 | 30 | NCE |  | AcCa 22:0 |
| 36 |  |  |  |  | ive |  |  |  |  |  |  |
| 476.3<br>734 |  |  |  | 1 | Posit<br>ive | 4.3 | 5.3 | 30 | NCE |  | AcCa 22:4 |
| 474.3<br>578 |  |  |  | 1 | Posit<br>ive | 3.5 | 4.5 | 30 | NCE |  | AcCa 22:5 |
| 472.3<br>421 |  |  |  | 1 | Posit<br>ive | 3.1 | 4.1 | 30 | NCE |  | AcCa 22:6 |
| 512.4<br>673 |  |  |  | 1 | Posit<br>ive | 9.5 | 10.5 | 30 | NCE |  | AcCa 24:0 |
| 300.2<br>897 |  |  |  | 1 | Posit<br>ive | 5.3 | 6.3 | 30 | NCE |  | AEA 16:0 |
| 328.3<br>21 |  |  |  | 1 | Posit<br>ive | 6.9 | 7.9 | 30 | NCE |  | AEA 18:0 |
| 326.3<br>054 |  |  |  | 1 | Posit<br>ive | 5.4 | 6.4 | 30 | NCE |  | AEA 18:1 |
| 324.2<br>897 |  |  |  | 1 | Posit<br>ive | 4.5 | 5.5 | 30 | NCE |  | AEA 18:2 |
| 540.5<br>35 |  |  |  | 1 | Posit<br>ive | 12.6 | 13.6 | 25 | NCE |  | Cer<br>18:0;2/16:0 |
| 624.6<br>289 |  |  |  | 1 | Posit<br>ive | 15.1 | 16.1 | 25 | NCE |  | Cer<br>18:0;2/22:0 |
| 638.6<br>446 |  |  |  | 1 | Posit<br>ive | 15.5 | 16.5 | 25 | NCE |  | Cer<br>18:0;2/23:0 |
| 652.6<br>602 |  |  |  | 1 | Posit<br>ive | 15.9 | 16.9 | 25 | NCE |  | Cer<br>18:0;2/24:0 |
| 650.6<br>446 |  |  |  | 1 | Posit<br>ive | 14.9 | 15.9 | 25 | NCE |  | Cer<br>18:0;2/24:1 |
| 666.6<br>759 |  |  |  | 1 | Posit<br>ive | 16.2 | 17.2 | 25 | NCE |  | Cer<br>18:0;2/25:0 |
| 482.4<br>568 |  |  |  | 1 | Posit<br>ive | 10.2 | 11.2 | 25 | NCE |  | Cer<br>18:1;2/12:0 |
| 538.5<br>194 |  |  |  | 1 | Posit<br>ive | 12.2 | 13.2 | 25 | NCE |  | Cer<br>18:1;2/16:0 |
| 566.5<br>507 |  |  |  | 1 | Posit<br>ive | 13.1 | 14.1 | 25 | NCE |  | Cer<br>18:1;2/18:0 |
| 594.5<br>82 |  |  |  | 1 | Posit<br>ive | 14 | 15 | 25 | NCE |  | Cer<br>18:1;2/20:0 |
| 622.6<br>133 |  |  |  | 1 | Posit<br>ive | 14.8 | 15.8 | 25 | NCE |  | Cer<br>18:1;2/22:0 |
| 636.6<br>289 |  |  |  | 1 | Posit<br>ive | 15.2 | 16.2 | 25 | NCE |  | Cer<br>18:1;2/23:0 |
| 650.6<br>446 |  |  |  | 1 | Posit<br>ive | 15.6 | 16.6 | 25 | NCE |  | Cer<br>18:1;2/24:0 |
| 648.6<br>289 |  |  |  | 1 | Posit<br>ive | 14.6 | 15.6 | 25 | NCE |  | Cer<br>18:1;2/24:1 |
| 646.6 |  |  |  | 1 | Posit | 13.9 | 14.9 | 25 | NCE |  | Cer |
| 133 |  |  |  |  | ive |  |  |  |  |  | 18:1;2/24:2 |
| 678.6<br>759 |  |  |  | 1 | Posit<br>ive | 16.3 | 17.3 | 25 | NCE |  | Cer<br>18:1;2/26:0 |
| 676.6<br>602 |  |  |  | 1 | Posit<br>ive | 15.3 | 16.3 | 25 | NCE |  | Cer<br>18:1;2/26:1 |
| 620.5<br>976 |  |  |  | 1 | Posit<br>ive | 14 | 15 | 25 | NCE |  | Cer<br>18:2;2/22:0 |
| 634.6<br>133 |  |  |  | 1 | Posit<br>ive | 14.4 | 15.4 | 25 | NCE |  | Cer<br>18:2;2/23:0 |
| 648.6<br>289 |  |  |  | 1 | Posit<br>ive | 14.8 | 15.8 | 25 | NCE |  | Cer<br>18:2;2/24:0 |
| 646.6<br>133 |  |  |  | 1 | Posit<br>ive | 13.9 | 14.9 | 25 | NCE |  | Cer<br>18:2;2/24:1 |
| 662.6<br>446 |  |  |  | 1 | Posit<br>ive | 15.2 | 16.2 | 25 | NCE |  | Cer<br>18:2;2/25:0 |
| 636.6<br>289 |  |  |  | 1 | Posit<br>ive | 15.1 | 16.1 | 25 | NCE |  | Cer<br>d19:1 22:0 |
| 650.6<br>446 |  |  |  | 1 | Posit<br>ive | 15.4 | 16.4 | 25 | NCE |  | Cer<br>d19:1 23:0 |
| 664.6<br>61 |  |  |  | 1 | Posit<br>ive | 15.7 | 16.7 | 25 | NCE |  | Cer<br>d19:1 24:0 |
| 662.6<br>446 |  |  |  | 1 | Posit<br>ive | 14.7 | 15.7 | 25 | NCE |  | Cer<br>d19:1 24:1 |
| 608.6<br>341 |  |  |  | 1 | Posit<br>ive | 15.6 | 16.6 | 25 | NCE |  | Cer<br>m18:0_22:<br>0 |
| 636.6<br>654 |  |  |  | 1 | Posit<br>ive | 16.2 | 17.2 | 25 | NCE |  | Cer<br>m18:0_24:<br>0 |
| 634.6<br>497 |  |  |  | 1 | Posit<br>ive | 15.2 | 16.2 | 25 | NCE |  | Cer<br>m18:0_24:<br>1 |
| 606.6<br>18 |  |  |  | 1 | Posit<br>ive | 14.8 | 15.8 | 25 | NCE |  | Cer<br>m18:1_22:<br>0 |
| 634.6<br>498 |  |  |  | 1 | Posit<br>ive | 15.6 | 16.6 | 25 | NCE |  | Cer<br>m18:1_24:<br>0 |
| 650.6<br>812 |  |  |  | 1 | Posit<br>ive | 16.5 | 17.5 | 25 | NCE |  | Cer<br>m19:0_24:<br>0 |
| 648.6<br>653 |  |  |  | 1 | Posit<br>ive | 15.4 | 16.4 | 25 | NCE |  | Cer<br>m19:0_24:<br>1 |
| 642.6<br>184 |  |  |  | 1 | Posit<br>ive | 19.3 | 20.3 | 10 | NCE |  | ChE 16:0 |
| 640.6<br>027 |  |  |  | 1 | Positive | 18.4 | 19.4 | 10 | NCE |  | ChE 16:1 |
| 670.6<br>497 |  |  |  | 1 | Positive | 19.9 | 20.9 | 10 | NCE |  | ChE 18:0 |
| 668.6<br>34 |  |  |  | 1 | Positive | 19.1 | 20.5 | 10 | NCE |  | ChE 18:1 |
| 675.6<br>779 |  |  |  | 1 | Positive | 19.1 | 20.5 | 10 | NCE |  | ChE 18:1D7 |
| 666.6<br>184 |  |  |  | 1 | Positive | 18.6 | 19.6 | 10 | NCE |  | ChE 18:2 |
| 664.6<br>027 |  |  |  | 1 | Positive | 18 | 19 | 10 | NCE |  | ChE 18:3 |
| 696.6<br>653 |  |  |  | 1 | Positive | 19.7 | 20.7 | 10 | NCE |  | ChE 20:1 |
| 694.6<br>497 |  |  |  | 1 | Positive | 19.3 | 20.3 | 10 | NCE |  | ChE 20:2 |
| 692.6<br>34 |  |  |  | 1 | Positive | 18.6 | 19.6 | 10 | NCE |  | ChE 20:3 |
| 690.6<br>184 |  |  |  | 1 | Positive | 18.2 | 19.5 | 10 | NCE |  | ChE 20:4 |
| 688.6<br>027 |  |  |  | 1 | Positive | 17.6 | 18.6 | 10 | NCE |  | ChE 20:5 |
| 718.6<br>497 |  |  |  | 1 | Positive | 18.7 | 19.7 | 10 | NCE |  | ChE 22:4 |
| 716.6<br>34 |  |  |  | 1 | Positive | 18.1 | 19.5 | 10 | NCE |  | ChE 22:5 |
| 714.6<br>184 |  |  |  | 1 | Positive | 18 | 19 | 10 | NCE |  | ChE 22:6 |
| 863.6<br>912 |  |  |  | 1 | Positive | 16.3 | 17.3 | 30 | NCE |  | CoQ10 |
| 584.5<br>249 |  |  |  | 1 | Positive | 12.8 | 13.8 | 20 | NCE |  | DAG 14:0-18:1 |
| 605.5<br>844 |  |  |  | 1 | Positive | 13.2 | 14.2 | 20 | NCE |  | DAG 15:0-18:1D7 |
| 586.5<br>405 |  |  |  | 1 | Positive | 13.7 | 14.7 | 20 | NCE |  | DAG 16:0-16:0 |
| 584.5<br>249 |  |  |  | 1 | Positive | 12.8 | 13.8 | 20 | NCE |  | DAG 16:0-16:1 |
| 614.5<br>718 |  |  |  | 1 | Positive | 14.5 | 15.5 | 20 | NCE |  | DAG 16:0-18:0 |
| 612.5<br>562 |  |  |  | 1 | Positive | 13.6 | 14.6 | 20 | NCE |  | DAG 16:0-18:1 |
| 610.5<br>405 |  |  |  | 1 | Positive | 12.9 | 13.9 | 20 | NCE |  | DAG 16:0-18:2 |
| 610.5<br>405 |  |  |  | 1 | Positive | 12.8 | 13.8 | 20 | NCE |  | DAG 16:1-18:1 |
| 608.5<br>249 |  |  |  | 1 | Posit<br>ive | 12.2 | 13.2 | 20 | NCE |  | DAG 16:1-18:2 |
| 642.6<br>031 |  |  |  | 1 | Posit<br>ive | 15.5 | 16.5 | 20 | NCE |  | DAG 18:0-18:0 |
| 640.5<br>875 |  |  |  | 1 | Posit<br>ive | 14.5 | 15.5 | 20 | NCE |  | DAG 18:0-18:1 |
| 638.5<br>718 |  |  |  | 1 | Posit<br>ive | 13.7 | 14.7 | 20 | NCE |  | DAG 18:0-18:2 |
| 638.5<br>718 |  |  |  | 1 | Posit<br>ive | 13.6 | 14.6 | 20 | NCE |  | DAG 18:1-18:1 |
| 636.5<br>562 |  |  |  | 1 | Posit<br>ive | 12.9 | 13.9 | 20 | NCE |  | DAG 18:1-18:2 |
| 634.5<br>405 |  |  |  | 1 | Posit<br>ive | 12.4 | 13.4 | 20 | NCE |  | DAG 18:1-18:3 |
| 668.6<br>188 |  |  |  | 1 | Posit<br>ive | 14.3 | 15.3 | 20 | NCE |  | DAG 18:1-20:0 |
| 662.5<br>718 |  |  |  | 1 | Posit<br>ive | 13.1 | 14.1 | 20 | NCE |  | DAG 18:1-20:3 |
| 660.5<br>562 |  |  |  | 1 | Posit<br>ive | 12.7 | 13.7 | 20 | NCE |  | DAG 18:1-20:4 |
| 634.5<br>405 |  |  |  | 1 | Posit<br>ive | 12.4 | 13.4 | 20 | NCE |  | DAG 18:2-18:2 |
| 632.5<br>249 |  |  |  | 1 | Posit<br>ive | 11.6 | 12.6 | 20 | NCE |  | DAG 18:2-18:3 |
| 806.5<br>624 |  |  |  | 1 | Posit<br>ive | 8.9 | 9.9 | 25 | NCE |  | Hex2Cer<br>18:1;2/12:0 |
| 834.5<br>937 |  |  |  | 1 | Posit<br>ive | 10.1 | 11.1 | 25 | NCE |  | Hex2Cer<br>18:1;2/14:0 |
| 862.6<br>25 |  |  |  | 1 | Posit<br>ive | 11.1 | 12.1 | 25 | NCE |  | Hex2Cer<br>18:1;2/16:0 |
| 890.6<br>563 |  |  |  | 1 | Posit<br>ive | 12.1 | 13.1 | 25 | NCE |  | Hex2Cer<br>18:1;2/18:0 |
| 918.6<br>876 |  |  |  | 1 | Posit<br>ive | 12.9 | 13.9 | 25 | NCE |  | Hex2Cer<br>18:1;2/20:0 |
| 946.7<br>189 |  |  |  | 1 | Posit<br>ive | 13.7 | 14.7 | 25 | NCE |  | Hex2Cer<br>18:1;2/22:0 |
| 974.7<br>502 |  |  |  | 1 | Posit<br>ive | 14.4 | 15.4 | 25 | NCE |  | Hex2Cer<br>18:1;2/24:0 |
| 972.7<br>346 |  |  |  | 1 | Posit<br>ive | 13.5 | 14.5 | 25 | NCE |  | Hex2Cer<br>18:1;2/24:1 |
| 970.7<br>189 |  |  |  | 1 | Posit<br>ive | 12.8 | 13.8 | 25 | NCE |  | Hex2Cer<br>18:1;2/24:2 |
| 860.6<br>094 |  |  |  | 1 | Posit<br>ive | 10.2 | 11.2 | 25 | NCE |  | Hex2Cer<br>18:2;2/16:0 |
| 1024.<br>678 |  |  |  | 1 | Posit<br>ive | 10.8 | 11.8 | 25 | NCE |  | Hex3Cer<br>18:1;2/16:0 |
| 1052.709 |  |  |  | 1 | Positive | 11.8 | 12.8 | 25 | NCE |  | Hex3Cer 18:1;2/18:0 |
| 1080.74 |  |  |  | 1 | Positive | 12.7 | 13.7 | 25 | NCE |  | Hex3Cer 18:1;2/20:0 |
| 1108.772 |  |  |  | 1 | Positive | 13.4 | 14.4 | 25 | NCE |  | Hex3Cer 18:1;2/22:0 |
| 1136.803 |  |  |  | 1 | Positive | 14.2 | 15.2 | 25 | NCE |  | Hex3Cer 18:1;2/24:0 |
| 1134.787 |  |  |  | 1 | Positive | 13.2 | 14.2 | 25 | NCE |  | Hex3Cer 18:1;2/24:1 |
| 644.5096 |  |  |  | 1 | Positive | 9.3 | 10.3 | 25 | NCE |  | HexCer 18:1;2/12:0 |
| 700.5722 |  |  |  | 1 | Positive | 11.5 | 12.5 | 25 | NCE |  | HexCer 18:1;2/16:0 |
| 728.6035 |  |  |  | 1 | Positive | 12.4 | 13.4 | 25 | NCE |  | HexCer 18:1;2/18:0 |
| 756.6348 |  |  |  | 1 | Positive | 13.2 | 14.2 | 25 | NCE |  | HexCer 18:1;2/20:0 |
| 784.6661 |  |  |  | 1 | Positive | 14 | 15 | 25 | NCE |  | HexCer 18:1;2/22:0 |
| 798.6818 |  |  |  | 1 | Positive | 14.4 | 15.4 | 25 | NCE |  | HexCer 18:1;2/23:0 |
| 812.6974 |  |  |  | 1 | Positive | 14.8 | 15.8 | 25 | NCE |  | HexCer 18:1;2/24:0 |
| 810.6818 |  |  |  | 1 | Positive | 13.8 | 14.8 | 25 | NCE |  | HexCer 18:1;2/24:1 |
| 808.6661 |  |  |  | 1 | Positive | 13.1 | 14.1 | 25 | NCE |  | HexCer 18:1;2/24:2 |
| 782.6505 |  |  |  | 1 | Positive | 13.3 | 14.3 | 25 | NCE |  | HexCer 18:2;2/22:0 |
| 810.6818 |  |  |  | 1 | Positive | 14.1 | 15.1 | 25 | NCE |  | HexCer 18:2;2/24:0 |
| 808.6661 |  |  |  | 1 | Positive | 13.1 | 14.1 | 25 | NCE |  | HexCer 18:2;2/24:1 |
| 468.3085 |  |  |  | 1 | Positive | 1.5 | 3 | 30 | NCE |  | LPC 14:0 |
| 454.3292 |  |  |  | 1 | Positive | 2.4 | 3.4 | 30 | NCE |  | LPC 14:0e |
| 482.3241 |  |  |  | 1 | Positive | 2.4 | 4 | 30 | NCE |  | LPC 15:0 |
| 496.3398 |  |  |  | 1 | Positive | 3.5 | 5.5 | 30 | NCE |  | LPC 16:0 |
| 482.3605 |  |  |  | 1 | Positive | 4.5 | 6 | 30 | NCE |  | LPC 16:0e |
| 494.3241 |  |  |  | 1 | Positive | 1.9 | 3.9 | 30 | NCE |  | LPC 16:1 |
| 480.3<br>449 |  |  |  | 1 | Positive | 4.5 | 5.5 | 30 | NCE |  | LPC 16:1e |
| 478.3<br>292 |  |  |  | 1 | Positive | 3.9 | 4.9 | 30 | NCE |  | LPC 16:2e |
| 510.3<br>554 |  |  |  | 1 | Positive | 4.2 | 6.2 | 30 | NCE |  | LPC 17:0 |
| 508.3<br>398 |  |  |  | 1 | Positive | 2.9 | 4.9 | 30 | NCE |  | LPC 17:1 |
| 524.3<br>711 |  |  |  | 1 | Positive | 5.2 | 7.2 | 30 | NCE |  | LPC 18:0 |
| 510.3<br>918 |  |  |  | 1 | Positive | 6.4 | 7.4 | 30 | NCE |  | LPC 18:0e |
| 522.3<br>554 |  |  |  | 1 | Positive | 3.9 | 5.9 | 30 | NCE |  | LPC 18:1 |
| 529.3<br>994 |  |  |  | 1 | Positive | 3.9 | 5.9 | 30 | NCE |  | LPC<br>18:1D7 |
| 508.3<br>762 |  |  |  | 1 | Positive | 4.7 | 6 | 30 | NCE |  | LPC 18:1e |
| 520.3<br>398 |  |  |  | 1 | Positive | 2.6 | 4.6 | 30 | NCE |  | LPC 18:2 |
| 518.3<br>241 |  |  |  | 1 | Positive | 1.8 | 3 | 30 | NCE |  | LPC 18:3 |
| 538.3<br>867 |  |  |  | 1 | Positive | 6 | 7.5 | 30 | NCE |  | LPC 19:0 |
| 536.3<br>711 |  |  |  | 1 | Positive | 4.9 | 6.5 | 30 | NCE |  | LPC 19:1 |
| 552.4<br>024 |  |  |  | 1 | Positive | 7 | 8.5 | 30 | NCE |  | LPC 20:0 |
| 538.4<br>231 |  |  |  | 1 | Positive | 7.9 | 8.9 | 30 | NCE |  | LPC 20:0e |
| 550.3<br>867 |  |  |  | 1 | Positive | 5.3 | 7 | 30 | NCE |  | LPC 20:1 |
| 548.3<br>711 |  |  |  | 1 | Positive | 4.3 | 5.6 | 30 | NCE |  | LPC 20:2 |
| 546.3<br>554 |  |  |  | 1 | Positive | 3.5 | 5 | 30 | NCE |  | LPC 20:3 |
| 544.3<br>398 |  |  |  | 1 | Positive | 2.6 | 4 | 30 | NCE |  | LPC 20:4 |
| 542.3<br>241 |  |  |  | 1 | Positive | 1.8 | 3 | 30 | NCE |  | LPC 20:5 |
| 580.4<br>337 |  |  |  | 1 | Positive | 8.4 | 9.7 | 30 | NCE |  | LPC 22:0 |
| 578.4<br>18 |  |  |  | 1 | Positive | 7.1 | 8.1 | 30 | NCE |  | LPC 22:1 |
| 574.3<br>867 |  |  |  | 1 | Positive | 4.9 | 5.9 | 30 | NCE |  | LPC 22:3 |
| 572.3<br>711 |  |  |  | 1 | Positive | 4.2 | 5.2 | 30 | NCE |  | LPC 22:4 |
| 570.3<br>554 |  |  |  | 1 | Positive | 3.2 | 4.7 | 30 | NCE |  | LPC 22:5 |
| 568.3<br>398 |  |  |  | 1 | Positive | 2.4 | 4 | 30 | NCE |  | LPC 22:6 |
| 608.4<br>65 |  |  |  | 1 | Positive | 9.7 | 11 | 30 | NCE |  | LPC 24:0 |
| 606.4<br>493 |  |  |  | 1 | Positive | 8.5 | 9.5 | 30 | NCE |  | LPC 24:1 |
| 604.4<br>337 |  |  |  | 1 | Positive | 7.3 | 8.3 | 30 | NCE |  | LPC 24:2 |
| 426.2<br>615 |  |  |  | 1 | Positive | 1.8 | 2.8 | 30 | NCE |  | LPE 14:0 |
| 454.2<br>928 |  |  |  | 1 | Positive | 3.7 | 5 | 30 | NCE |  | LPE 16:0 |
| 482.3<br>241 |  |  |  | 1 | Positive | 5.4 | 6.7 | 30 | NCE |  | LPE 18:0 |
| 480.3<br>085 |  |  |  | 1 | Positive | 4 | 5.3 | 30 | NCE |  | LPE 18:1 |
| 487.3<br>524 |  |  |  | 1 | Positive | 4 | 5.3 | 30 | NCE |  | LPE<br>18:1D7 |
| 478.2<br>928 |  |  |  | 1 | Positive | 2.6 | 4 | 30 | NCE |  | LPE 18:2 |
| 504.3<br>085 |  |  |  | 1 | Positive | 3.4 | 4.7 | 30 | NCE |  | LPE 20:3 |
| 502.2<br>928 |  |  |  | 1 | Positive | 2.6 | 3.9 | 30 | NCE |  | LPE 20:4 |
| 526.2<br>928 |  |  |  | 1 | Positive | 2.5 | 3.8 | 30 | NCE |  | LPE 22:6 |
| 647.5<br>123 |  |  |  | 1 | Positive | 8.8 | 10.8 | 30 | NCE |  | SM(d30:1) |
| 645.4<br>966 |  |  |  | 1 | Positive | 7.6 | 8.6 | 30 | NCE |  | SM(d30:2) |
| 677.5<br>592 |  |  |  | 1 | Positive | 10.5 | 11.5 | 30 | NCE |  | SM(d32:0) |
| 675.5<br>436 |  |  |  | 1 | Positive | 10 | 12 | 30 | NCE |  | SM(d32:1) |
| 673.5<br>279 |  |  |  | 1 | Positive | 8.9 | 10.5 | 30 | NCE |  | SM(d32:2) |
| 705.5<br>905 |  |  |  | 1 | Positive | 11.5 | 12.5 | 30 | NCE |  | SM(d34:0) |
| 703.5<br>749 |  |  |  | 1 | Positive | 11.1 | 12.5 | 30 | NCE |  | SM(d34:1) |
| 701.5<br>592 |  |  |  | 1 | Positive | 10.1 | 12 | 30 | NCE |  | SM(d34:2) |
| 699.5<br>436 |  |  |  | 1 | Positive | 9.3 | 11 | 30 | NCE |  | SM(d34:3) |
| 733.6<br>218 |  |  |  | 1 | Positive | 12.2 | 13.6 | 30 | NCE |  | SM(d36:0) |
| 731.6<br>062 |  |  |  | 1 | Positive | 12.1 | 13.5 | 30 | NCE |  | SM(d36:1) |
| 729.5<br>905 |  |  |  | 1 | Positive | 11 | 13 | 30 | NCE |  | SM(d36:2) |
| 738.6<br>47 |  |  |  | 1 | Positive | 11 | 13 | 30 | NCE |  | SM(d36:2<br>D9) |
| 727.5<br>749 |  |  |  | 1 | Positive | 10.4 | 12.4 | 30 | NCE |  | SM(d36:3) |
| 759.6<br>375 |  |  |  | 1 | Positive | 13 | 14.5 | 30 | NCE |  | SM(d38:1) |
| 757.6<br>218 |  |  |  | 1 | Positive | 12.2 | 13.5 | 30 | NCE |  | SM(d38:2) |
| 755.6<br>062 |  |  |  | 1 | Positive | 11.5 | 13 | 30 | NCE |  | SM(d38:3) |
| 787.6<br>688 |  |  |  | 1 | Positive | 13.8 | 16 | 30 | NCE |  | SM(d40:1) |
| 785.6<br>531 |  |  |  | 1 | Positive | 13 | 14.5 | 30 | NCE |  | SM(d40:2) |
| 783.6<br>375 |  |  |  | 1 | Positive | 12 | 13.5 | 30 | NCE |  | SM(d40:3) |
| 815.7<br>001 |  |  |  | 1 | Positive | 14.6 | 16.6 | 30 | NCE |  | SM(d42:1) |
| 813.6<br>844 |  |  |  | 1 | Positive | 13.5 | 15.5 | 30 | NCE |  | SM(d42:2) |
| 811.6<br>688 |  |  |  | 1 | Positive | 12.8 | 14.5 | 30 | NCE |  | SM(d42:3) |
| 845.7<br>47 |  |  |  | 1 | Positive | 16.2 | 17.2 | 30 | NCE |  | SM(d44:0) |
| 843.7<br>314 |  |  |  | 1 | Positive | 15.3 | 17 | 30 | NCE |  | SM(d44:1) |
| 841.7<br>157 |  |  |  | 1 | Positive | 14.4 | 16 | 30 | NCE |  | SM(d44:2) |
| 839.7<br>001 |  |  |  | 1 | Positive | 13.6 | 15 | 30 | NCE |  | SM(d44:3) |
| 288.2<br>897 |  |  |  | 1 | Positive | 2 | 4 | 20 | NCE |  | SPH d17:0 |
| 286.2<br>741 |  |  |  | 1 | Positive | 2 | 3.5 | 20 | NCE |  | SPH d17:1 |
| 302.3<br>054 |  |  |  | 1 | Positive | 3 | 5 | 20 | NCE |  | SPH d18:0 |
| 300.2<br>897 |  |  |  | 1 | Positive | 3 | 4.5 | 20 | NCE |  | SPH d18:1 |
| 352.2<br>247 |  |  |  | 1 | Positive | 1 | 3 | 20 | NCE |  | SPHP<br>d16:1 |
| 368.2<br>56 |  |  |  | 1 | Positive | 1 | 5 | 20 | NCE |  | SPHP<br>d17:0 |
| 366.2<br>404 |  |  |  | 1 | Positive | 1 | 5 | 20 | NCE |  | SPHP<br>d17:1 |
| 382.2<br>717 |  |  |  | 1 | Positive | 2 | 6 | 20 | NCE |  | SPHP<br>d18:0 |
| 380.2<br>56 |  |  |  | 1 | Positive | 2 | 5 | 20 | NCE |  | SPHP<br>d18:1 |
| 378.2<br>404 |  |  |  | 1 | Positive | 1 | 4 | 20 | NCE |  | SPHP<br>d18:2 |
| 766.6<br>919 |  |  |  | 1 | Positive | 16.8 | 17.8 | 20 | NCE |  | TAG 12:0-14:0-18:1 |
| 780.7<br>076 |  |  |  | 1 | Positive | 17.1 | 18.1 | 20 | NCE |  | TAG 12:0-15:0-18:1 |
| 768.7<br>076 |  |  |  | 1 | Positive | 17.6 | 18.6 | 20 | NCE |  | TAG 12:0-16:0-16:0 |
| 796.7<br>389 |  |  |  | 1 | Positive | 18.1 | 19.1 | 20 | NCE |  | TAG 12:0-16:0-18:0 |
| 794.7<br>232 |  |  |  | 1 | Positive | 17.4 | 18.4 | 20 | NCE |  | TAG 12:0-16:0-18:1 |
| 792.7<br>076 |  |  |  | 1 | Positive | 16.9 | 17.9 | 20 | NCE |  | TAG 12:0-16:0-18:2 |
| 818.7<br>232 |  |  |  | 1 | Positive | 17 | 18 | 20 | NCE |  | TAG 12:0-18:1-18:2 |
| 816.7<br>076 |  |  |  | 1 | Positive | 16.4 | 17.4 | 20 | NCE |  | TAG 12:0-18:1-18:3 |
| 808.7<br>389 |  |  |  | 1 | Positive | 17.7 | 18.7 | 20 | NCE |  | TAG 14:0-15:0-18:1 |
| 806.7<br>232 |  |  |  | 1 | Positive | 17.2 | 18.2 | 20 | NCE |  | TAG 14:0-15:0-18:2 |
| 796.7<br>389 |  |  |  | 1 | Positive | 18.2 | 19.2 | 20 | NCE |  | TAG 14:0-16:0-16:0 |
| 810.7<br>545 |  |  |  | 1 | Positive | 18.4 | 19.4 | 20 | NCE |  | TAG 14:0-16:0-17:0 |
| 824.7<br>702 |  |  |  | 1 | Positive | 18.9 | 19.9 | 20 | NCE |  | TAG 14:0-16:0-18:0 |
| 822.7<br>545 |  |  |  | 1 | Positive | 18.2 | 19.2 | 20 | NCE |  | TAG 14:0-16:0-18:1 |
| 820.7<br>389 |  |  |  | 1 | Positive | 17.5 | 18.5 | 20 | NCE |  | TAG 14:0-16:0-18:2 |
| 868.7<br>389 |  |  |  | 1 | Positive | 17 | 18 | 20 | NCE |  | TAG 14:0-16:0-22:6 |
| 834.7<br>545 |  |  |  | 1 | Positive | 17.7 | 18.7 | 20 | NCE |  | TAG 14:0-17:1-18:1 |
| 848.7<br>702 |  |  |  | 1 | Posit<br>ive | 18 | 19 | 20 | NCE |  | TAG 14:0-18:1-18:1 |
| 844.7<br>389 |  |  |  | 1 | Posit<br>ive | 16.9 | 17.9 | 20 | NCE |  | TAG 14:0-18:2-18:2 |
| 842.7<br>232 |  |  |  | 1 | Posit<br>ive | 16.5 | 17.5 | 20 | NCE |  | TAG 14:0-18:2-18:3 |
| 820.7<br>389 |  |  |  | 1 | Posit<br>ive | 17.5 | 18.5 | 20 | NCE |  | TAG 14:1-16:0-18:1 |
| 840.7<br>076 |  |  |  | 1 | Posit<br>ive | 15.9 | 16.9 | 20 | NCE |  | TAG 14:1-18:2-18:3 |
| 932.8<br>641 |  |  |  | 1 | Posit<br>ive | 19.8 | 20.8 | 20 | NCE |  | TAG 14:2-20:0-22:0 |
| 838.7<br>858 |  |  |  | 1 | Posit<br>ive | 19 | 20 | 20 | NCE |  | TAG 15:0-16:0-18:0 |
| 836.7<br>702 |  |  |  | 1 | Posit<br>ive | 18.4 | 19.4 | 20 | NCE |  | TAG 15:0-16:0-18:1 |
| 834.7<br>545 |  |  |  | 1 | Posit<br>ive | 17.7 | 18.7 | 20 | NCE |  | TAG 15:0-16:0-18:2 |
| 832.7<br>389 |  |  |  | 1 | Posit<br>ive | 17.2 | 18.2 | 20 | NCE |  | TAG 15:0-16:0-18:3 |
| 806.7<br>232 |  |  |  | 1 | Posit<br>ive | 17.2 | 18.2 | 20 | NCE |  | TAG 15:0-16:1-16:1 |
| 832.7<br>389 |  |  |  | 1 | Posit<br>ive | 17.2 | 18.2 | 20 | NCE |  | TAG 15:0-16:1-18:2 |
| 862.7<br>858 |  |  |  | 1 | Posit<br>ive | 18.3 | 19.3 | 20 | NCE |  | TAG 15:0-18:1-18:1 |
| 860.7<br>702 |  |  |  | 1 | Posit<br>ive | 17.8 | 18.8 | 20 | NCE |  | TAG 15:0-18:1-18:2 |
| 858.7<br>545 |  |  |  | 1 | Posit<br>ive | 17.3 | 18.3 | 20 | NCE |  | TAG 15:0-18:2-18:2 |
| 856.7<br>389 |  |  |  | 1 | Posit<br>ive | 16.8 | 17.8 | 20 | NCE |  | TAG 15:0-18:2-18:3 |
| 836.7<br>702 |  |  |  | 1 | Posit<br>ive | 18.4 | 19.4 | 20 | NCE |  | TAG 16:0-16:0-17:1 |
| 852.8<br>015 |  |  |  | 1 | Posit<br>ive | 19.5 | 20.5 | 20 | NCE |  | TAG 16:0-16:0-18:0 |
| 850.7<br>858 |  |  |  | 1 | Posit<br>ive | 18.7 | 19.7 | 20 | NCE |  | TAG 16:0-16:0-18:1 |
| 848.7<br>702 |  |  |  | 1 | Posit<br>ive | 18.2 | 19.2 | 20 | NCE |  | TAG 16:0-16:0-18:2 |
| 846.7<br>545 |  |  |  | 1 | Posit<br>ive | 18 | 19 | 20 | NCE |  | TAG 16:0-16:0-18:3 |
| 844.7<br>389 |  |  |  | 1 | Posit<br>ive | 16.9 | 17.9 | 20 | NCE |  | TAG 16:0-16:1-18:3 |
| 866.8<br>171 |  |  |  | 1 | Posit<br>ive | 19.6 | 20.6 | 20 | NCE |  | TAG 16:0-17:0-18:0 |
| 864.8<br>015 |  |  |  | 1 | Positive | 18.9 | 19.9 | 20 | NCE |  | TAG 16:0-17:0-18:1 |
| 862.7<br>858 |  |  |  | 1 | Positive | 18.3 | 19.3 | 20 | NCE |  | TAG 16:0-17:1-18:1 |
| 860.7<br>702 |  |  |  | 1 | Positive | 17.8 | 18.8 | 20 | NCE |  | TAG 16:0-17:1-18:2 |
| 880.8<br>328 |  |  |  | 1 | Positive | 19.8 | 20.8 | 20 | NCE |  | TAG 16:0-18:0-18:0 |
| 878.8<br>171 |  |  |  | 1 | Positive | 19.3 | 20.3 | 20 | NCE |  | TAG 16:0-18:0-18:1 |
| 908.8<br>641 |  |  |  | 1 | Positive | 20.4 | 21.4 | 20 | NCE |  | TAG 16:0-18:0-20:0 |
| 936.8<br>954 |  |  |  | 1 | Positive | 20.7 | 21.7 | 20 | NCE |  | TAG 16:0-18:0-22:0 |
| 876.8<br>015 |  |  |  | 1 | Positive | 18.6 | 19.6 | 20 | NCE |  | TAG 16:0-18:1-18:1 |
| 874.7<br>858 |  |  |  | 1 | Positive | 18.1 | 19.1 | 20 | NCE |  | TAG 16:0-18:1-18:2 |
| 872.7<br>702 |  |  |  | 1 | Positive | 17.5 | 18.5 | 20 | NCE |  | TAG 16:0-18:1-18:3 |
| 898.7<br>858 |  |  |  | 1 | Positive | 17.9 | 18.9 | 20 | NCE |  | TAG 16:0-18:1-20:4 |
| 924.8<br>015 |  |  |  | 1 | Positive | 17.8 | 18.8 | 20 | NCE |  | TAG 16:0-18:1-22:5 |
| 922.7<br>858 |  |  |  | 1 | Positive | 17.7 | 18.7 | 20 | NCE |  | TAG 16:0-18:1-22:6 |
| 872.7<br>702 |  |  |  | 1 | Positive | 17.5 | 18.5 | 20 | NCE |  | TAG 16:0-18:2-18:2 |
| 870.7<br>545 |  |  |  | 1 | Positive | 17.1 | 18.1 | 20 | NCE |  | TAG 16:0-18:2-18:3 |
| 868.7<br>389 |  |  |  | 1 | Positive | 16.6 | 17.6 | 20 | NCE |  | TAG 16:0-18:3-18:3 |
| 944.7<br>702 |  |  |  | 1 | Positive | 16.5 | 17.5 | 20 | NCE |  | TAG 16:0-20:4-22:6 |
| 892.8<br>328 |  |  |  | 1 | Positive | 19.4 | 20.4 | 20 | NCE |  | TAG 17:0-18:0-18:1 |
| 890.8<br>171 |  |  |  | 1 | Positive | 18.9 | 19.9 | 20 | NCE |  | TAG 17:0-18:1-18:1 |
| 888.8<br>015 |  |  |  | 1 | Positive | 18.4 | 19.4 | 20 | NCE |  | TAG 17:0-18:1-18:2 |
| 886.7<br>858 |  |  |  | 1 | Positive | 17.8 | 18.8 | 20 | NCE |  | TAG 17:0-18:1-18:3 |
| 884.7<br>702 |  |  |  | 1 | Positive | 17.2 | 18.2 | 20 | NCE |  | TAG 17:0-18:2-18:3 |
| 910.7<br>858 |  |  |  | 1 | Positive | 17.7 | 18.7 | 20 | NCE |  | TAG 17:0-18:2-20:4 |
| 890.8<br>171 |  |  |  | 1 | Posit<br>ive | 18.9 | 19.9 | 20 | NCE |  | TAG 17:1-18:0-18:1 |
| 888.8<br>015 |  |  |  | 1 | Posit<br>ive | 18.4 | 19.4 | 20 | NCE |  | TAG 17:1-18:1-18:1 |
| 886.7<br>858 |  |  |  | 1 | Posit<br>ive | 17.8 | 18.8 | 20 | NCE |  | TAG 17:1-18:1-18:2 |
| 910.7<br>858 |  |  |  | 1 | Posit<br>ive | 17.7 | 18.7 | 20 | NCE |  | TAG 17:1-18:1-20:4 |
| 884.7<br>702 |  |  |  | 1 | Posit<br>ive | 17.2 | 18.2 | 20 | NCE |  | TAG 17:1-18:2-18:2 |
| 908.8<br>641 |  |  |  | 1 | Posit<br>ive | 20.3 | 21.3 | 20 | NCE |  | TAG 18:0-18:0-18:0 |
| 906.8<br>484 |  |  |  | 1 | Posit<br>ive | 19.7 | 20.7 | 20 | NCE |  | TAG 18:0-18:0-18:1 |
| 936.8<br>954 |  |  |  | 1 | Posit<br>ive | 20.7 | 21.7 | 20 | NCE |  | TAG 18:0-18:0-20:0 |
| 928.8<br>328 |  |  |  | 1 | Posit<br>ive | 18.6 | 19.6 | 20 | NCE |  | TAG 18:0-18:0-20:4 |
| 904.8<br>328 |  |  |  | 1 | Posit<br>ive | 19.3 | 20.3 | 20 | NCE |  | TAG 18:0-18:1-18:1 |
| 902.8<br>171 |  |  |  | 1 | Posit<br>ive | 18.7 | 19.7 | 20 | NCE |  | TAG 18:0-18:1-18:2 |
| 920.8<br>641 |  |  |  | 1 | Posit<br>ive | 19.9 | 20.9 | 20 | NCE |  | TAG 18:0-18:1-19:0 |
| 918.8<br>484 |  |  |  | 1 | Posit<br>ive | 19.4 | 20.4 | 20 | NCE |  | TAG 18:0-18:1-19:1 |
| 934.8<br>797 |  |  |  | 1 | Posit<br>ive | 20.1 | 21.1 | 20 | NCE |  | TAG 18:0-18:1-20:0 |
| 928.8<br>328 |  |  |  | 1 | Posit<br>ive | 18.6 | 19.6 | 20 | NCE |  | TAG 18:0-18:1-20:3 |
| 962.9<br>11 |  |  |  | 1 | Posit<br>ive | 20.6 | 21.6 | 20 | NCE |  | TAG 18:0-18:1-22:0 |
| 954.8<br>484 |  |  |  | 1 | Posit<br>ive | 18.9 | 19.9 | 20 | NCE |  | TAG 18:0-18:1-22:4 |
| 952.8<br>328 |  |  |  | 1 | Posit<br>ive | 18.3 | 19.3 | 20 | NCE |  | TAG 18:0-18:1-22:5 |
| 900.8<br>015 |  |  |  | 1 | Posit<br>ive | 18.2 | 19.2 | 20 | NCE |  | TAG 18:0-18:2-18:2 |
| 916.8<br>328 |  |  |  | 1 | Posit<br>ive | 18.9 | 19.9 | 20 | NCE |  | TAG 18:0-18:2-19:1 |
| 948.8<br>015 |  |  |  | 1 | Posit<br>ive | 17.6 | 18.6 | 20 | NCE |  | TAG 18:0-18:2-22:6 |
| 902.8<br>171 |  |  |  | 1 | Posit<br>ive | 18.7 | 19.7 | 20 | NCE |  | TAG 18:1-18:1-18:1 |
| 900.8<br>015 |  |  |  | 1 | Posit<br>ive | 18 | 19 | 20 | NCE |  | TAG 18:1-18:1-18:2 |
| 898.7<br>858 |  |  |  | 1 | Positive | 17.5 | 18.5 | 20 | NCE |  | TAG 18:1-18:1-18:3 |
| 932.8<br>641 |  |  |  | 1 | Positive | 19.8 | 20.8 | 20 | NCE |  | TAG 18:1-18:1-20:0 |
| 930.8<br>484 |  |  |  | 1 | Positive | 19 | 20 | 20 | NCE |  | TAG 18:1-18:1-20:1 |
| 926.8<br>171 |  |  |  | 1 | Positive | 18.3 | 19.3 | 20 | NCE |  | TAG 18:1-18:1-20:3 |
| 960.8<br>954 |  |  |  | 1 | Positive | 20.3 | 21.3 | 20 | NCE |  | TAG 18:1-18:1-22:0 |
| 958.8<br>797 |  |  |  | 1 | Positive | 19.7 | 20.7 | 20 | NCE |  | TAG 18:1-18:1-22:1 |
| 952.8<br>328 |  |  |  | 1 | Positive | 18.3 | 19.3 | 20 | NCE |  | TAG 18:1-18:1-22:4 |
| 950.8<br>171 |  |  |  | 1 | Positive | 17.8 | 18.8 | 20 | NCE |  | TAG 18:1-18:1-22:5 |
| 974.9<br>11 |  |  |  | 1 | Positive | 20.3 | 21.3 | 20 | NCE |  | TAG 18:1-18:1-23:0 |
| 986.9<br>11 |  |  |  | 1 | Positive | 20.1 | 21.1 | 20 | NCE |  | TAG 18:1-18:1-24:1 |
| 1016.<br>958 |  |  |  | 1 | Positive | 20.8 | 21.8 | 20 | NCE |  | TAG 18:1-18:1-26:0 |
| 896.7<br>702 |  |  |  | 1 | Positive | 17 | 18 | 20 | NCE |  | TAG 18:1-18:2-18:3 |
| 930.8<br>484 |  |  |  | 1 | Positive | 19.2 | 20.2 | 20 | NCE |  | TAG 18:1-18:2-20:0 |
| 928.8<br>328 |  |  |  | 1 | Positive | 18.6 | 19.6 | 20 | NCE |  | TAG 18:1-18:2-20:1 |
| 922.7<br>858 |  |  |  | 1 | Positive | 17.3 | 18.3 | 20 | NCE |  | TAG 18:1-18:2-20:4 |
| 958.8<br>797 |  |  |  | 1 | Positive | 19.7 | 20.7 | 20 | NCE |  | TAG 18:1-18:2-22:0 |
| 956.8<br>641 |  |  |  | 1 | Positive | 19 | 20 | 20 | NCE |  | TAG 18:1-18:2-22:1 |
| 948.8<br>015 |  |  |  | 1 | Positive | 17.3 | 18.3 | 20 | NCE |  | TAG 18:1-18:2-22:5 |
| 946.7<br>858 |  |  |  | 1 | Positive | 17.1 | 18.1 | 20 | NCE |  | TAG 18:1-18:2-22:6 |
| 986.9<br>11 |  |  |  | 1 | Positive | 20.1 | 21.1 | 20 | NCE |  | TAG 18:1-18:2-24:0 |
| 984.8<br>954 |  |  |  | 1 | Positive | 19.5 | 20.5 | 20 | NCE |  | TAG 18:1-18:2-24:1 |
| 1014.<br>942 |  |  |  | 1 | Positive | 20.5 | 21.5 | 20 | NCE |  | TAG 18:1-18:2-26:0 |
| 1012.<br>927 |  |  |  | 1 | Positive | 19.9 | 20.9 | 20 | NCE |  | TAG 18:1-18:2-26:1 |
| 896.7<br>702 |  |  |  | 1 | Positive | 17 | 18 | 20 | NCE |  | TAG 18:2-18:2-18:2 |
| 894.7<br>545 |  |  |  | 1 | Positive | 16.4 | 17.4 | 20 | NCE |  | TAG 18:2-18:2-18:3 |
| 928.8<br>328 |  |  |  | 1 | Positive | 18.6 | 19.6 | 20 | NCE |  | TAG 18:2-18:2-20:0 |
| 920.7<br>702 |  |  |  | 1 | Positive | 16.8 | 17.8 | 20 | NCE |  | TAG 18:2-18:2-20:4 |
| 944.7<br>702 |  |  |  | 1 | Positive | 16.5 | 17.5 | 20 | NCE |  | TAG 18:2-18:2-22:6 |
| 892.7<br>389 |  |  |  | 1 | Positive | 15.9 | 16.9 | 20 | NCE |  | TAG 18:2-18:3-18:3 |
| 920.7<br>702 |  |  |  | 1 | Positive | 16.8 | 17.8 | 20 | NCE |  | TAG 18:2-18:3-20:3 |
| 918.7<br>545 |  |  |  | 1 | Positive | 16.3 | 17.3 | 20 | NCE |  | TAG 18:2-18:3-20:4 |
| 829.7<br>985 |  |  |  | 1 | Positive | 18.2 | 19.2 | 20 | NCE |  | TG<br>15:0_18:1<br>D7_15:0 |
| 720.5<br>902 |  |  |  | 1 | Positive | 12.6<br>7 | 14 | 30 | NCE |  | PC(16:0e_<br>16:0) |
| 768.5<br>901 |  |  |  | 1 | Positive | 11.8<br>5 | 12.8<br>5 | 30 | NCE |  | PC(16:0e_<br>20:4) |
| 796.6<br>215 |  |  |  | 1 | Positive | 12.5 | 14 | 30 | NCE |  | PC(16:0e_<br>22:4) |
| 748.6<br>215 |  |  |  | 1 | Positive | 13.5<br>3 | 14.5<br>3 | 30 | NCE |  | PC(18:0e_<br>16:0) |
| 774.6<br>371 |  |  |  | 1 | Positive | 13.5<br>1 | 14.5<br>1 | 30 | NCE |  | PC(18:0e_<br>18:1) |
| 798.6<br>372 |  |  |  | 1 | Positive | 13.0<br>5 | 14.0<br>5 | 30 | NCE |  | PC(18:0e_<br>20:3) |
| 824.6<br>528 |  |  |  | 1 | Positive | 13.5 | 14.5 | 30 | NCE |  | PC(18:0e_<br>22:4) |
| 820.6<br>21 |  |  |  | 1 | Positive | 12.4<br>7 | 13.4<br>7 | 30 | NCE |  | PC(18:0e_<br>22:6) |
| 702.5<br>432 |  |  |  | 1 | Positive | 12.7<br>4 | 13.7<br>4 | 30 | NCE |  | PE(16:0p_<br>18:1) |
| 700.5<br>275 |  |  |  | 1 | Positive | 12.1 | 13.1 | 30 | NCE |  | PE(16:0p_<br>18:2) |
| 724.5<br>275 |  |  |  | 1 | Positive | 11.9<br>5 | 12.9<br>5 | 30 | NCE |  | PE(16:0p_<br>20:4) |
| 722.5<br>119 |  |  |  | 1 | Positive | 11.3<br>7 | 12.3<br>7 | 30 | NCE |  | PE(16:0p_<br>20:5) |
| 752.5<br>58 |  |  |  | 1 | Positive | 12.5 | 14 | 30 | NCE |  | PE(16:0p_<br>22:4) |
| 748.5<br>275 |  |  |  | 1 | Positive | 11.7<br>1 | 12.7<br>1 | 30 | NCE |  | PE(16:0p_22:6) |
| 730.5<br>745 |  |  |  | 1 | Positive | 13.5<br>6 | 14.5<br>6 | 30 | NCE |  | PE(18:0p_18:1) |
| 728.5<br>588 |  |  |  | 1 | Positive | 13.2 | 14.2 | 30 | NCE |  | PE(18:0p_18:2) |
| 752.5<br>587 |  |  |  | 1 | Positive | 12.7 | 13.7 | 30 | NCE |  | PE(18:0p_20:4) |
| 780.5<br>902 |  |  |  | 1 | Positive | 13.3<br>1 | 14.3<br>1 | 30 | NCE |  | PE(18:0p_22:4) |
| 778.5<br>745 |  |  |  | 1 | Positive | 12.7<br>9 | 13.7<br>9 | 30 | NCE |  | PE(18:0p_22:5) |
| 776.5<br>588 |  |  |  | 1 | Positive | 12.5<br>6 | 13.5<br>6 | 30 | NCE |  | PE(18:0p_22:6) |
| 750.5<br>432 |  |  |  | 1 | Positive | 11.8 | 13 | 30 | NCE |  | PE(18:1p_20:4) |
| 774.5<br>411 |  |  |  | 1 | Positive | 11.6<br>5 | 12.6<br>5 | 30 | NCE |  | PE(18:1p_22:6) |
| 753.6<br>13 |  |  |  | 1 | Positive | 11.5<br>5 | 13 | 30 | NCE |  | PC(18:1D7_15:0) |
| 711.5<br>664 |  |  |  | 1 | Positive | 11.8 | 12.8 | 30 | NCE |  | PE(18:1D7_15:0) |
| Mass<br>[m/z] | Form<br>ula<br>[M] | Formu<br>la type | Spe<br>cies | CS<br>[z] | Polar<br>ity | Start<br>[min<br>] | End<br>[min<br>] | (N)<br>CE | (N)C<br>E<br>type | MS<br>X<br>ID | Comment |
| 599.3<br>202 |  |  |  | 1 | Negative | 4.5 | 6 |  |  |  | LPI (18:0) |
| 597.3<br>045 |  |  |  | 1 | Negative | 2.5 | 4.5 |  |  |  | LPI (18:1) |
| 595.2<br>889 |  |  |  | 1 | Negative | 1.5 | 3 |  |  |  | LPI (18:2) |
| 621.3<br>045 |  |  |  | 1 | Negative | 2 | 3.5 |  |  |  | LPI (20:3) |
| 619.2<br>889 |  |  |  | 1 | Negative | 1.5 | 3 |  |  |  | LPI (20:4) |
| 433.2<br>361 |  |  |  | 1 | Negative | 2.5 | 4 |  |  |  | LPA (18:2) |
| 903.4<br>631 |  |  |  | 2 | Negative | 8.95 | 9.95 |  |  |  | GD1a(d34:1)-2H |
| 917.4<br>788 |  |  |  | 2 | Negative | 10.0<br>3 | 11.0<br>3 |  |  |  | GD1a(d36:1)-2H |
| 720.8<br>97 |  |  |  | 2 | Negative | 9.64 | 10.6<br>4 |  |  |  | GD3(d34:1)-2H |
| 1123.<br>675 |  |  |  | 1 | Negative | 8.84 | 9.84 |  |  |  | GM3(d32:1)-H |
| 1151. |  |  |  | 1 | Nega | 9.94 | 10.9 |  |  |  | GM3(d34: |
| 706 |  |  |  |  | tive |  | 4 |  |  |  | 1)-H |
| 1149.69 |  |  |  | 1 | Negative | 9 | 10 |  |  |  | GM3(d34:2)-H |
| 1179.737 |  |  |  | 1 | Negative | 10.91 | 11.91 |  |  |  | GM3(d36:1)-H |
| 1235.8 |  |  |  | 1 | Negative | 12.51 | 13.51 |  |  |  | GM3(d40:1)-H |
| 1263.831 |  |  |  | 1 | Negative | 13.19 | 14.19 |  |  |  | GM3(d42:1)-H |
| 701.5127 |  |  |  | 1 | Negative | 12.68 | 13.68 |  |  |  | PA 18:0-18:1 |
| 699.497 |  |  |  | 1 | Negative | 11.91 | 12.91 |  |  |  | PA 18:0-18:2 |
| 723.497 |  |  |  | 1 | Negative | 11.86 | 12.86 |  |  |  | PA 18:0-20:4 |
| 747.497 |  |  |  | 1 | Negative | 11.6 | 12.6 |  |  |  | PA 18:0-22:6 |
| 666.5097 |  |  |  | 1 | Negative | 11.23 | 13 |  |  |  | PA(18:1D7_15:0) |
| 750.5291 |  |  |  | 1 | Negative | 11.08 | 12.08 |  |  |  | PC 14:0-16:0 |
| 776.5447 |  |  |  | 1 | Negative | 11.15 | 12.15 |  |  |  | PC 14:0-18:1 |
| 774.5291 |  |  |  | 1 | Negative | 10.5 | 11.5 |  |  |  | PC 14:0-18:2 |
| 798.5291 |  |  |  | 1 | Negative | 10.19 | 11.19 |  |  |  | PC 14:0-20:4 |
| 790.5604 |  |  |  | 1 | Negative | 11.7 | 12.7 |  |  |  | PC 15:0-18:1 |
| 788.5447 |  |  |  | 1 | Negative | 10.86 | 11.86 |  |  |  | PC 15:0-18:2 |
| 778.5604 |  |  |  | 1 | Negative | 12.01 | 13.51 |  |  |  | PC 16:0-16:0 |
| 776.5447 |  |  |  | 1 | Negative | 11.2 | 12.5 |  |  |  | PC 16:0-16:1 |
| 806.5917 |  |  |  | 1 | Negative | 13 | 14.5 |  |  |  | PC 16:0-18:0 |
| 804.576 |  |  |  | 1 | Negative | 12.01 | 13.5 |  |  |  | PC 16:0-18:1 |
| 802.5604 |  |  |  | 1 | Negative | 11.5 | 13 |  |  |  | PC 16:0-18:2 |
| 800.5447 |  |  |  | 1 | Negative | 11 | 12 |  |  |  | PC 16:0-18:3 |
| 832.6073 |  |  |  | 1 | Negative | 12.83 | 13.83 |  |  |  | PC 16:0-20:1 |
| 830.5 |  |  |  | 1 | Negative | 12.2 | 13.2 |  |  |  | PC 16:0- |
| 917 |  |  |  |  | tive |  |  |  |  |  | 20:2 |
| 828.5<br>76 |  |  |  | 1 | Nega<br>tive | 11.5<br>7 | 12.5<br>7 |  |  |  | PC 16:0-20:3 |
| 826.5<br>604 |  |  |  | 1 | Nega<br>tive | 11.1<br>8 | 12.5<br>8 |  |  |  | PC 16:0-20:4 |
| 824.5<br>447 |  |  |  | 1 | Nega<br>tive | 10.8 | 11.8 |  |  |  | PC 16:0-20:5 |
| 854.5<br>917 |  |  |  | 1 | Nega<br>tive | 11.8<br>2 | 12.8<br>2 |  |  |  | PC 16:0-22:4 |
| 852.5<br>76 |  |  |  | 1 | Nega<br>tive | 11.2<br>4 | 12.2<br>4 |  |  |  | PC 16:0-22:5 |
| 850.5<br>604 |  |  |  | 1 | Nega<br>tive | 10.9<br>7 | 11.9<br>7 |  |  |  | PC 16:0-22:6 |
| 800.5<br>447 |  |  |  | 1 | Nega<br>tive | 10.7 | 11.7 |  |  |  | PC 16:1-18:2 |
| 824.5<br>447 |  |  |  | 1 | Nega<br>tive | 10.2<br>7 | 11.2<br>7 |  |  |  | PC 16:1-20:4 |
| 818.5<br>917 |  |  |  | 1 | Nega<br>tive | 12.5 | 14 |  |  |  | PC 17:0-18:1 |
| 816.5<br>76 |  |  |  | 1 | Nega<br>tive | 11.7<br>9 | 13 |  |  |  | PC 17:0-18:2 |
| 840.5<br>76 |  |  |  | 1 | Nega<br>tive | 11.8<br>6 | 12.8<br>6 |  |  |  | PC 17:0-20:4 |
| 834.6<br>23 |  |  |  | 1 | Nega<br>tive | 14 | 15 |  |  |  | PC 18:0-18:0 |
| 832.6<br>073 |  |  |  | 1 | Nega<br>tive | 12.8<br>3 | 14.5 |  |  |  | PC 18:0-18:1 |
| 830.5<br>917 |  |  |  | 1 | Nega<br>tive | 12.3 | 14 |  |  |  | PC 18:0-18:2 |
| 828.5<br>76 |  |  |  | 1 | Nega<br>tive | 12 | 13 |  |  |  | PC 18:0-18:3 |
| 858.6<br>23 |  |  |  | 1 | Nega<br>tive | 13.2 | 14.2 |  |  |  | PC 18:0-20:2 |
| 856.6<br>073 |  |  |  | 1 | Nega<br>tive | 12.7 | 13.7 |  |  |  | PC 18:0-20:3 |
| 854.5<br>917 |  |  |  | 1 | Nega<br>tive | 12.3 | 13.5 |  |  |  | PC 18:0-20:4 |
| 852.5<br>76 |  |  |  | 1 | Nega<br>tive | 11.5<br>4 | 12.5<br>4 |  |  |  | PC 18:0-20:5 |
| 882.6<br>23 |  |  |  | 1 | Nega<br>tive | 13 | 14 |  |  |  | PC 18:0-22:4 |
| 880.6<br>073 |  |  |  | 1 | Nega<br>tive | 12.3 | 13.5 |  |  |  | PC 18:0-22:5 |
| 878.5<br>917 |  |  |  | 1 | Nega<br>tive | 11.9 | 12.9 |  |  |  | PC 18:0-22:6 |
| 830.5 |  |  |  | 1 | Nega | 12.0 | 13.0 |  |  |  | PC 18:1- |
| 917 |  |  |  |  | tive | 1 | 1 |  |  |  | 18:1 |
| 828.5<br>76 |  |  |  | 1 | Nega<br>tive | 11.5<br>7 | 12.5<br>7 |  |  |  | PC 18:1-18:2 |
| 846.6<br>23 |  |  |  | 1 | Nega<br>tive | 13.1<br>6 | 14.1<br>6 |  |  |  | PC 18:1-19:0 |
| 854.5<br>917 |  |  |  | 1 | Nega<br>tive | 11.8<br>2 | 12.8<br>2 |  |  |  | PC 18:1-20:3 |
| 852.5<br>76 |  |  |  | 1 | Nega<br>tive | 11.2<br>4 | 12.2<br>4 |  |  |  | PC 18:1-20:4 |
| 850.5<br>604 |  |  |  | 1 | Nega<br>tive | 10.4<br>9 | 11.4<br>9 |  |  |  | PC 18:1-20:5 |
| 878.5<br>917 |  |  |  | 1 | Nega<br>tive | 11.9 | 12.9 |  |  |  | PC 18:1-22:5 |
| 876.5<br>76 |  |  |  | 1 | Nega<br>tive | 10.9<br>6 | 11.9<br>6 |  |  |  | PC 18:1-22:6 |
| 826.5<br>604 |  |  |  | 1 | Nega<br>tive | 10.6<br>3 | 11.6<br>3 |  |  |  | PC 18:2-18:2 |
| 824.5<br>447 |  |  |  | 1 | Nega<br>tive | 9.97 | 10.9<br>7 |  |  |  | PC 18:2-18:3 |
| 858.6<br>23 |  |  |  | 1 | Nega<br>tive | 13.3 | 14.3 |  |  |  | PC 18:2-20:0 |
| 852.5<br>76 |  |  |  | 1 | Nega<br>tive | 10.9<br>7 | 11.9<br>7 |  |  |  | PC 18:2-20:3 |
| 850.5<br>604 |  |  |  | 1 | Nega<br>tive | 10.4<br>6 | 11.4<br>6 |  |  |  | PC 18:2-20:4 |
| 876.5<br>76 |  |  |  | 1 | Nega<br>tive | 10.7 | 11.7 |  |  |  | PC 20:3-20:4 |
| 874.5<br>604 |  |  |  | 1 | Nega<br>tive | 10.2<br>6 | 11.2<br>6 |  |  |  | PC 20:4-20:4 |
| 797.6<br>043 |  |  |  | 1 | Nega<br>tive | 11.5<br>5 | 13 |  |  |  | PC(18:1D7<br>_15:0) |
| 718.5<br>392 |  |  |  | 1 | Nega<br>tive | 13.0<br>9 | 14.0<br>9 |  |  |  | PE 16:0-18:0 |
| 716.5<br>236 |  |  |  | 1 | Nega<br>tive | 12.2<br>7 | 13.2<br>7 |  |  |  | PE 16:0-18:1 |
| 714.5<br>079 |  |  |  | 1 | Nega<br>tive | 11.5<br>5 | 12.5<br>5 |  |  |  | PE 16:0-18:2 |
| 738.5<br>079 |  |  |  | 1 | Nega<br>tive | 11.4<br>7 | 12.4<br>7 |  |  |  | PE 16:0-20:4 |
| 766.5<br>392 |  |  |  | 1 | Nega<br>tive | 12.0<br>4 | 13.0<br>4 |  |  |  | PE 16:0-22:4 |
| 764.5<br>236 |  |  |  | 1 | Nega<br>tive | 11.4<br>8 | 12.4<br>8 |  |  |  | PE 16:0-22:5 |
| 762.5<br>079 |  |  |  | 1 | Nega<br>tive | 11.2<br>1 | 12.2<br>1 |  |  |  | PE 16:0-22:6 |
| 744.5 |  |  |  | 1 | Nega | 13.0 | 14.0 |  |  |  | PE 18:0- |
| 549 |  |  |  |  | tive | 5 | 5 |  |  |  | 18:1 |
| 742.5<br>392 |  |  |  | 1 | Nega<br>tive | 12.4<br>5 | 13.4<br>5 |  |  |  | PE 18:0-18:2 |
| 768.5<br>549 |  |  |  | 1 | Nega<br>tive | 12.6<br>4 | 13.6<br>4 |  |  |  | PE 18:0-20:3 |
| 766.5<br>392 |  |  |  | 1 | Nega<br>tive | 12.3<br>3 | 13.3<br>3 |  |  |  | PE 18:0-20:4 |
| 794.5<br>705 |  |  |  | 1 | Nega<br>tive | 12.8<br>3 | 13.8<br>3 |  |  |  | PE 18:0-22:4 |
| 792.5<br>549 |  |  |  | 1 | Nega<br>tive | 12.3<br>4 | 13.3<br>4 |  |  |  | PE 18:0-22:5 |
| 790.5<br>392 |  |  |  | 1 | Nega<br>tive | 12.1 | 13.1 |  |  |  | PE 18:0-22:6 |
| 742.5<br>392 |  |  |  | 1 | Nega<br>tive | 12.2<br>3 | 13.2<br>3 |  |  |  | PE 18:1-18:1 |
| 740.5<br>236 |  |  |  | 1 | Nega<br>tive | 11.5<br>5 | 12.5<br>5 |  |  |  | PE 18:1-18:2 |
| 764.5<br>236 |  |  |  | 1 | Nega<br>tive | 11.4<br>8 | 12.4<br>8 |  |  |  | PE 18:1-20:4 |
| 738.5<br>079 |  |  |  | 1 | Nega<br>tive | 10.8<br>5 | 11.8<br>5 |  |  |  | PE 18:2-18:2 |
| 786.5<br>079 |  |  |  | 1 | Nega<br>tive | 10.4<br>9 | 11.4<br>9 |  |  |  | PE 20:4-20:4 |
| 709.5<br>519 |  |  |  | 1 | Nega<br>tive | 11.8 | 12.8 |  |  |  | PE(18:1D7<br>15:0) |
| 788.5<br>447 |  |  |  | 1 | Nega<br>tive | 12.1 | 13.1 |  |  |  | PS 18:0-18:1 |
| 810.5<br>291 |  |  |  | 1 | Nega<br>tive | 11.3<br>2 | 12.3<br>2 |  |  |  | PS 18:0-20:4 |
| 838.5<br>604 |  |  |  | 1 | Nega<br>tive | 11.9<br>1 | 12.9<br>1 |  |  |  | PS 18:0-22:4 |
| 836.5<br>447 |  |  |  | 1 | Nega<br>tive | 11.1 | 12.1 |  |  |  | PS 18:1-22:4 |
| 830.4<br>978 |  |  |  | 1 | Nega<br>tive | 9.48 | 10.4<br>8 |  |  |  | PS 20:4-20:4 |
| 858.5<br>291 |  |  |  | 1 | Nega<br>tive | 10.2<br>6 | 11.2<br>6 |  |  |  | PS 20:4-22:4 |
| 753.5<br>417 |  |  |  | 1 | Nega<br>tive | 10.7<br>1 | 11.7<br>1 |  |  |  | PS(18:1D7<br>15:0) |
| 828.5<br>625 |  |  |  | 1 | Nega<br>tive | 10.6 | 11.6 |  |  |  | PI(18:1D7<br>15:0) |
| 861.5<br>499 |  |  |  | 1 | Nega<br>tive | 11.2<br>7 | 12.2<br>7 |  |  |  | PI 18:0-18:2 |
| 859.5<br>342 |  |  |  | 1 | Nega<br>tive | 10.6<br>4 | 11.6<br>4 |  |  |  | PI 16:0-20:3 |
| 857.5 |  |  |  | 1 | Nega | 10.2 | 11.2 |  |  |  | PI 16:0- |
| 186 |  |  |  |  | tive | 7 | 7 |  |  |  | 20:4 |
| 871.5<br>342 |  |  |  | 1 | Nega<br>tive | 10.7<br>4 | 11.7<br>4 |  |  |  | PI 17:0-20:4 |
| 863.5<br>655 |  |  |  | 1 | Nega<br>tive | 11.9<br>4 | 12.9<br>4 |  |  |  | PI 18:0-18:1 |
| 887.5<br>655 |  |  |  | 1 | Nega<br>tive | 11.5<br>2 | 12.5<br>2 |  |  |  | PI 18:0-20:3 |
| 885.5<br>499 |  |  |  | 1 | Nega<br>tive | 11.2 | 12.2 |  |  |  | PI 18:0-20:4 |
| 883.5<br>342 |  |  |  | 1 | Nega<br>tive | 10.3 | 11.3 |  |  |  | PI 18:0-20:5 |
| 913.5<br>812 |  |  |  | 1 | Nega<br>tive | 11.7<br>9 | 12.7<br>9 |  |  |  | PI 18:0-22:4 |
| 911.5<br>655 |  |  |  | 1 | Nega<br>tive | 11.2<br>3 | 12.2<br>3 |  |  |  | PI 18:0-22:5 |
| 909.5<br>499 |  |  |  | 1 | Nega<br>tive | 10.9<br>7 | 11.9<br>7 |  |  |  | PI 18:0-22:6 |
| 861.5<br>499 |  |  |  | 1 | Nega<br>tive | 11.2<br>7 | 12.2<br>7 |  |  |  | PI 18:1-18:1 |
| 859.5<br>342 |  |  |  | 1 | Nega<br>tive | 10.3<br>8 | 11.3<br>8 |  |  |  | PI 18:1-18:2 |
| 883.5<br>342 |  |  |  | 1 | Nega<br>tive | 10.3 | 11.3 |  |  |  | PI 18:1-20:4 |

**Supplementary Tables 5.**
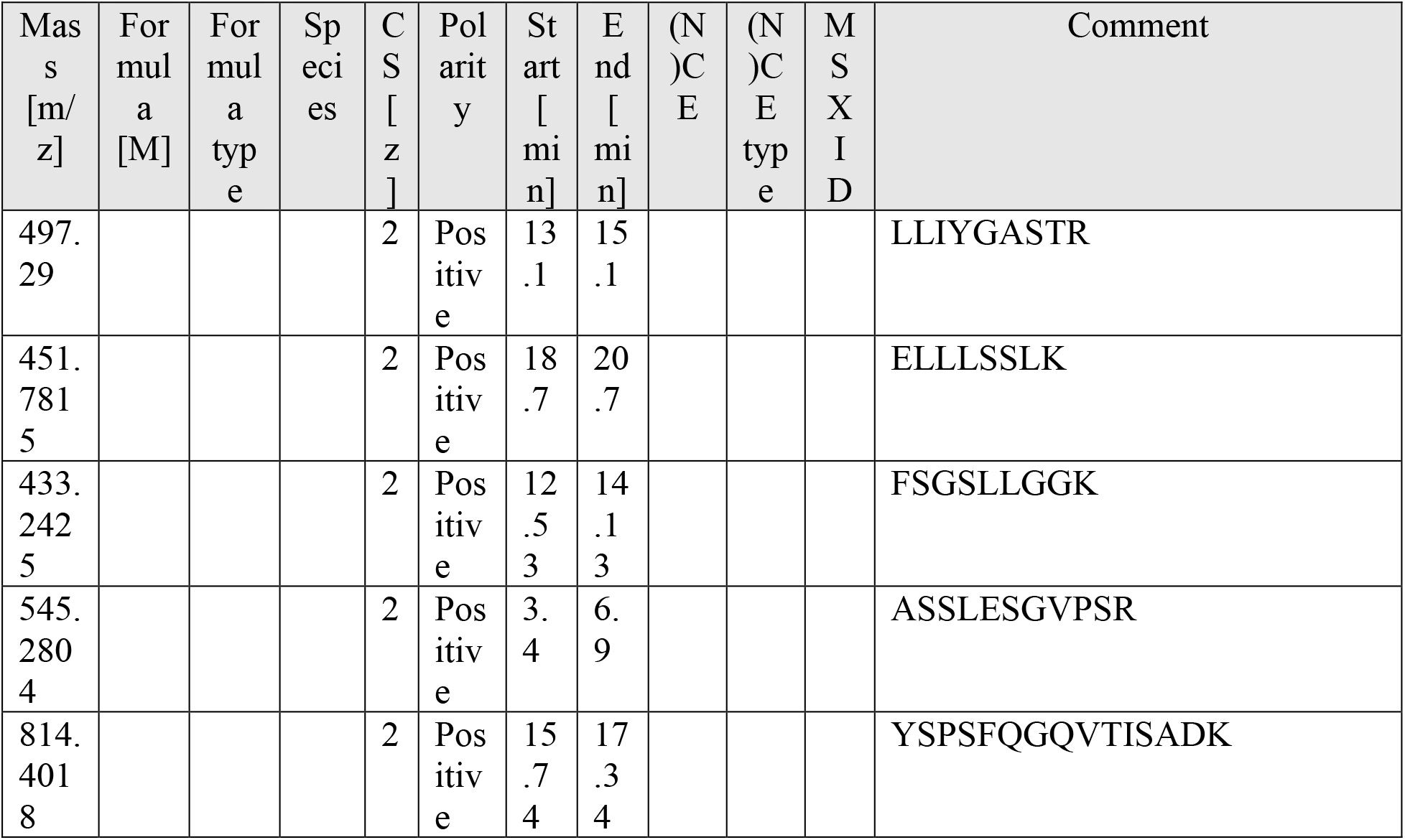
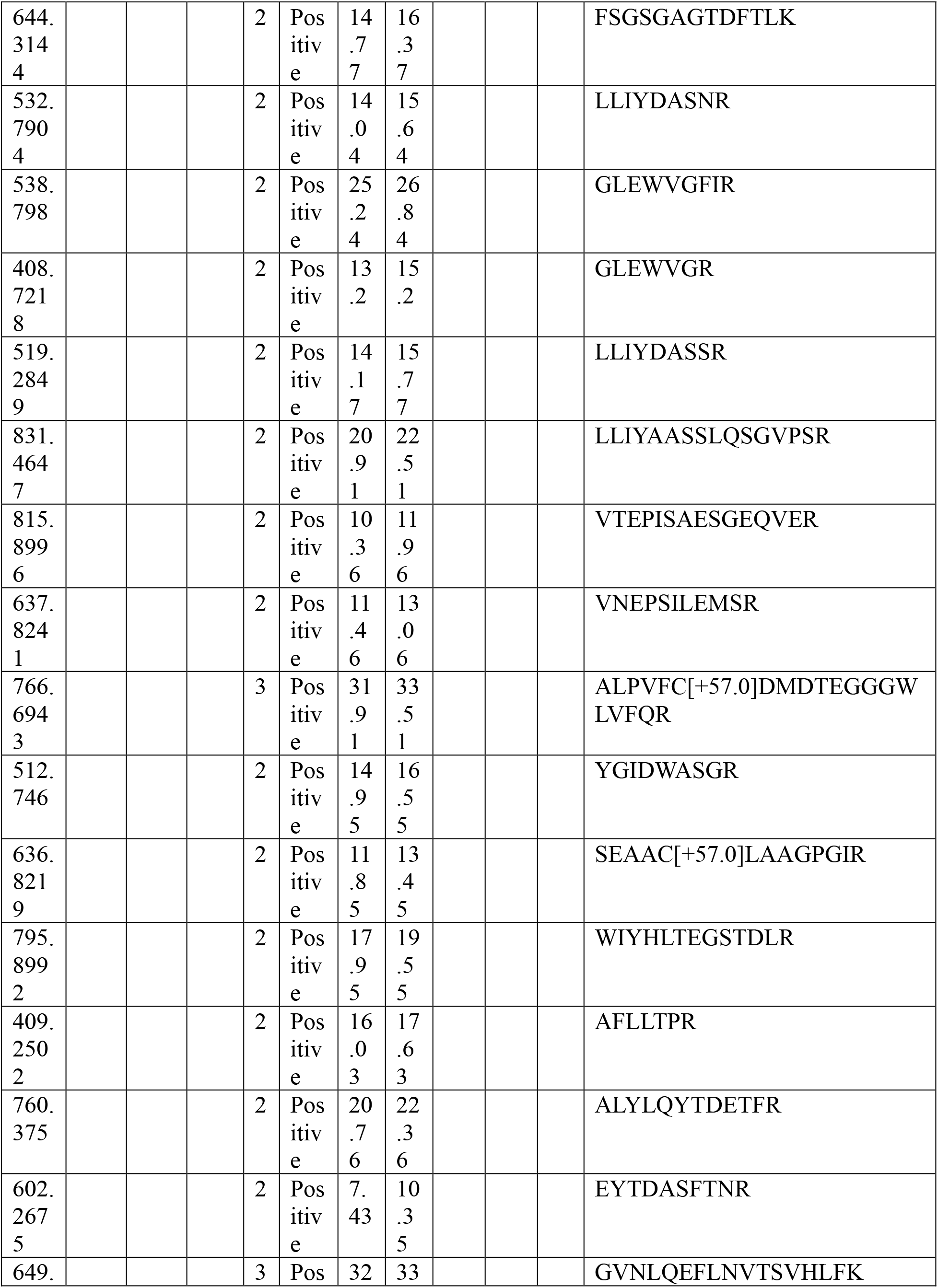
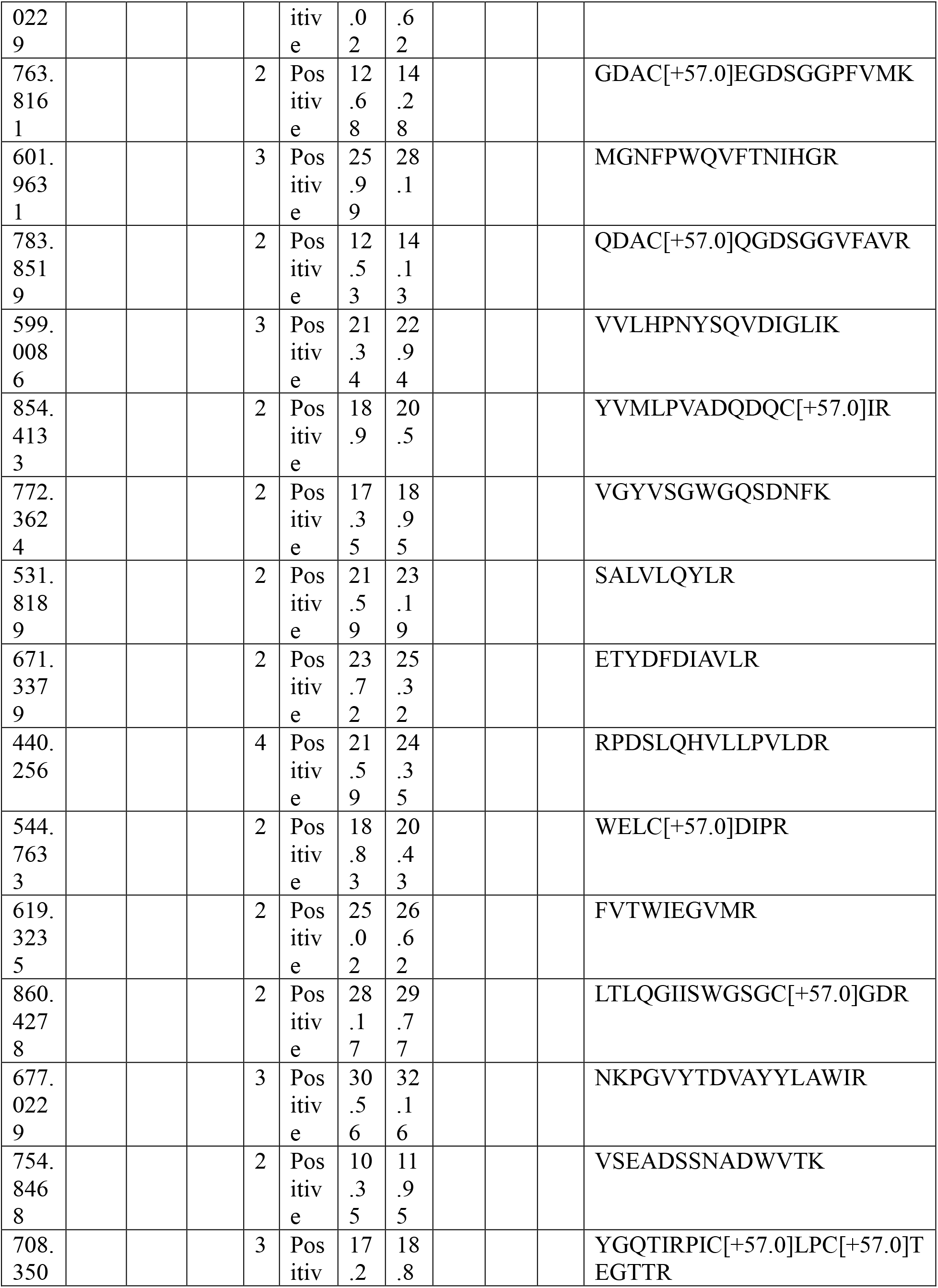
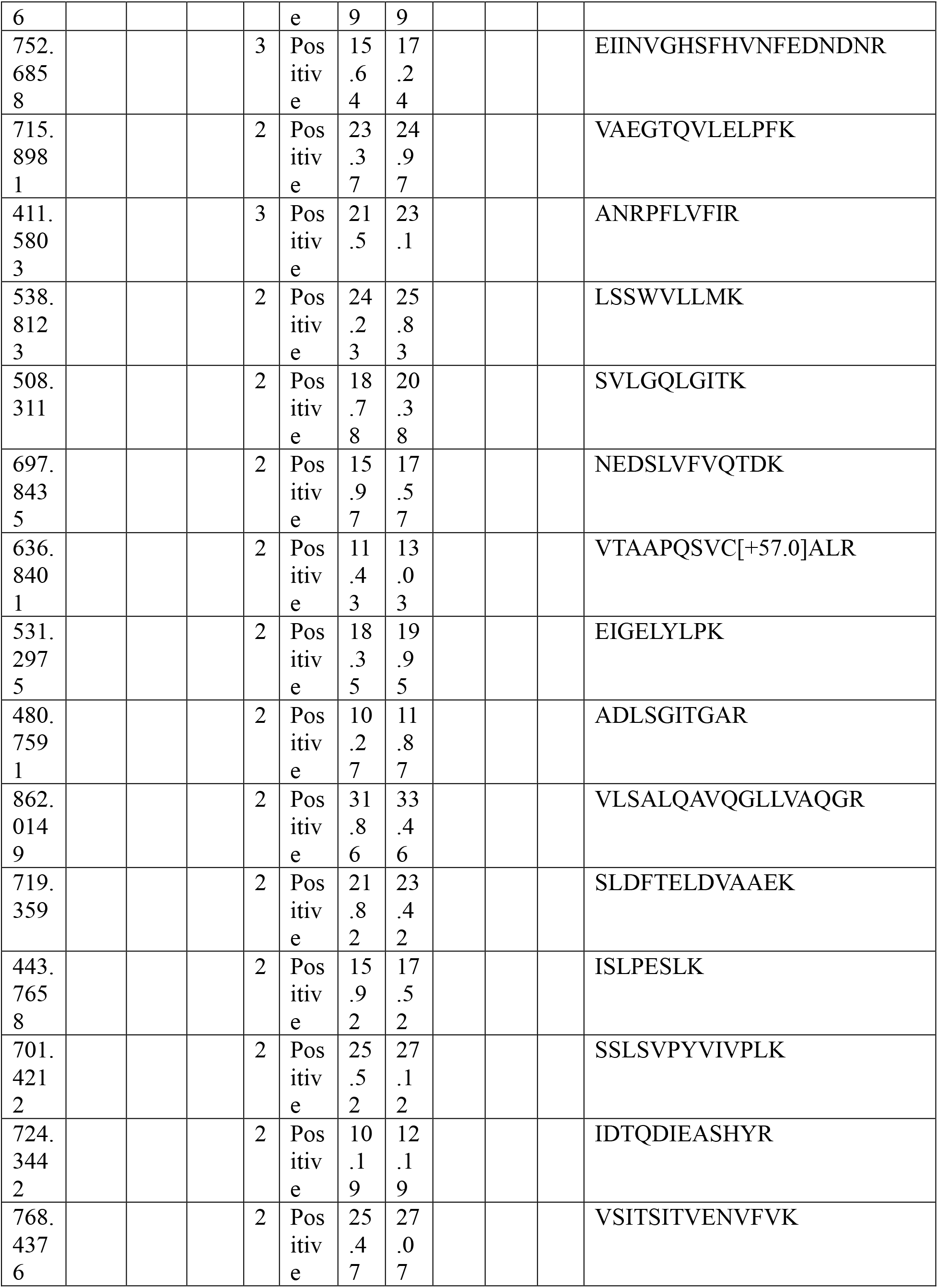
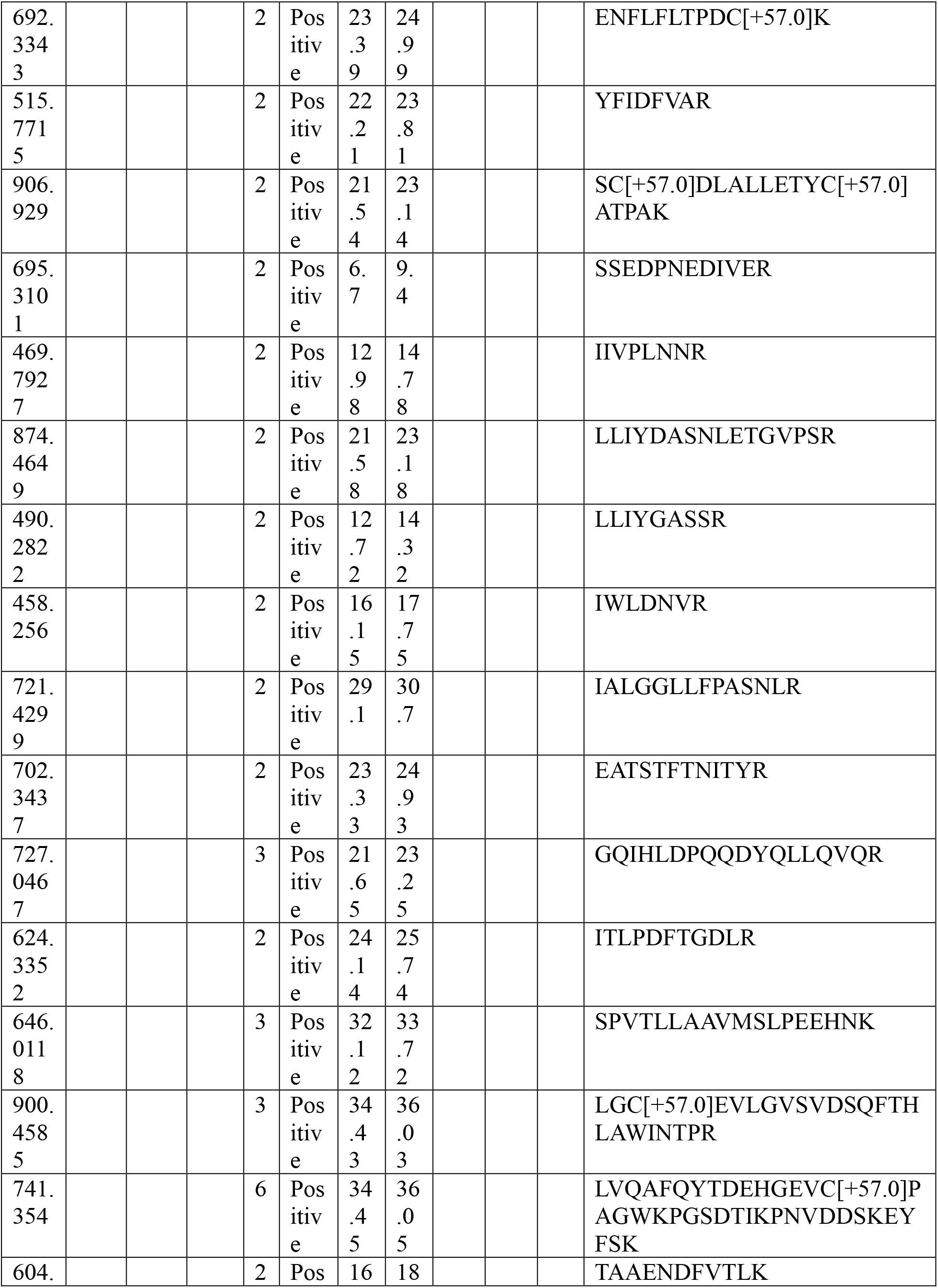
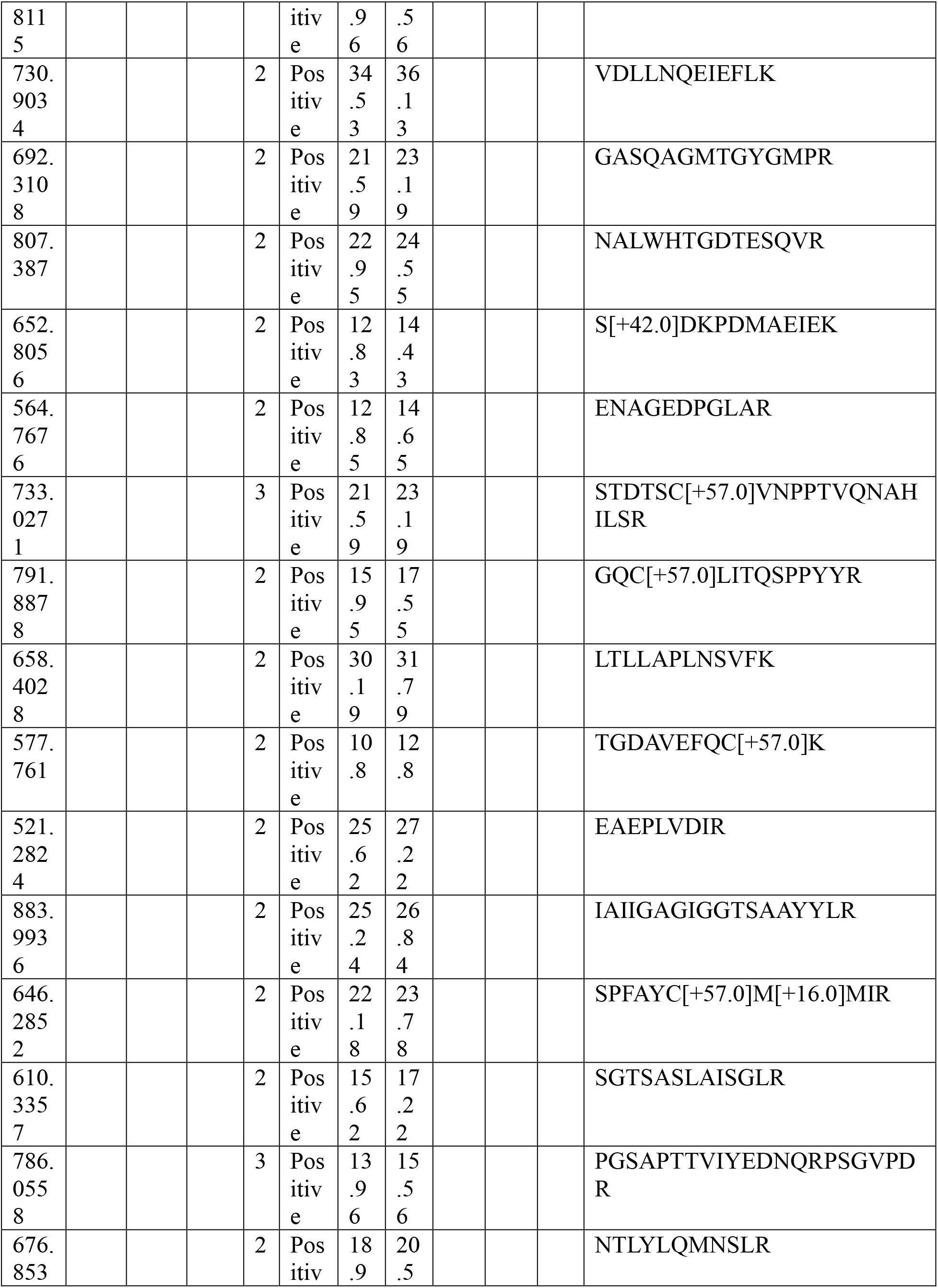
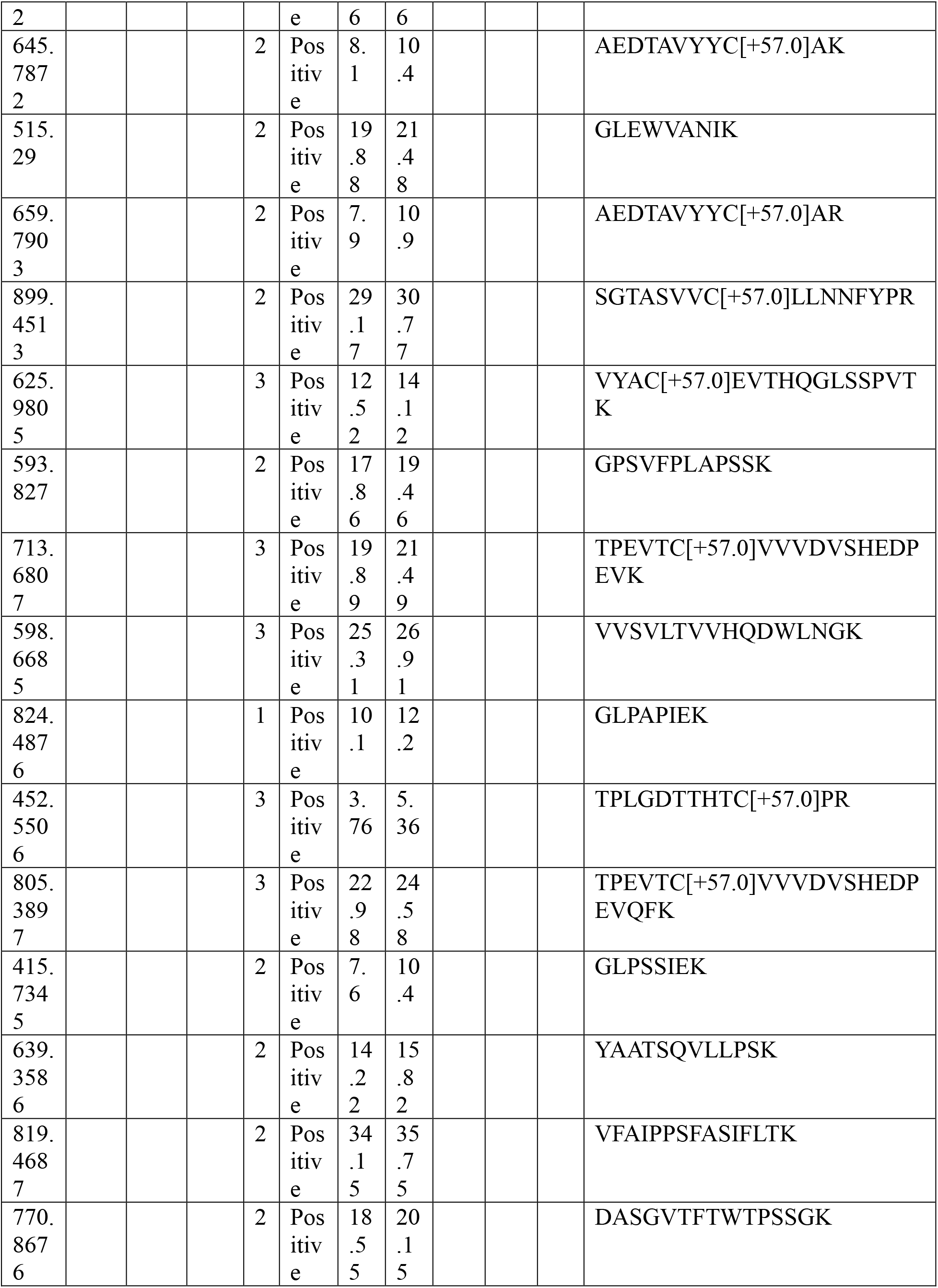
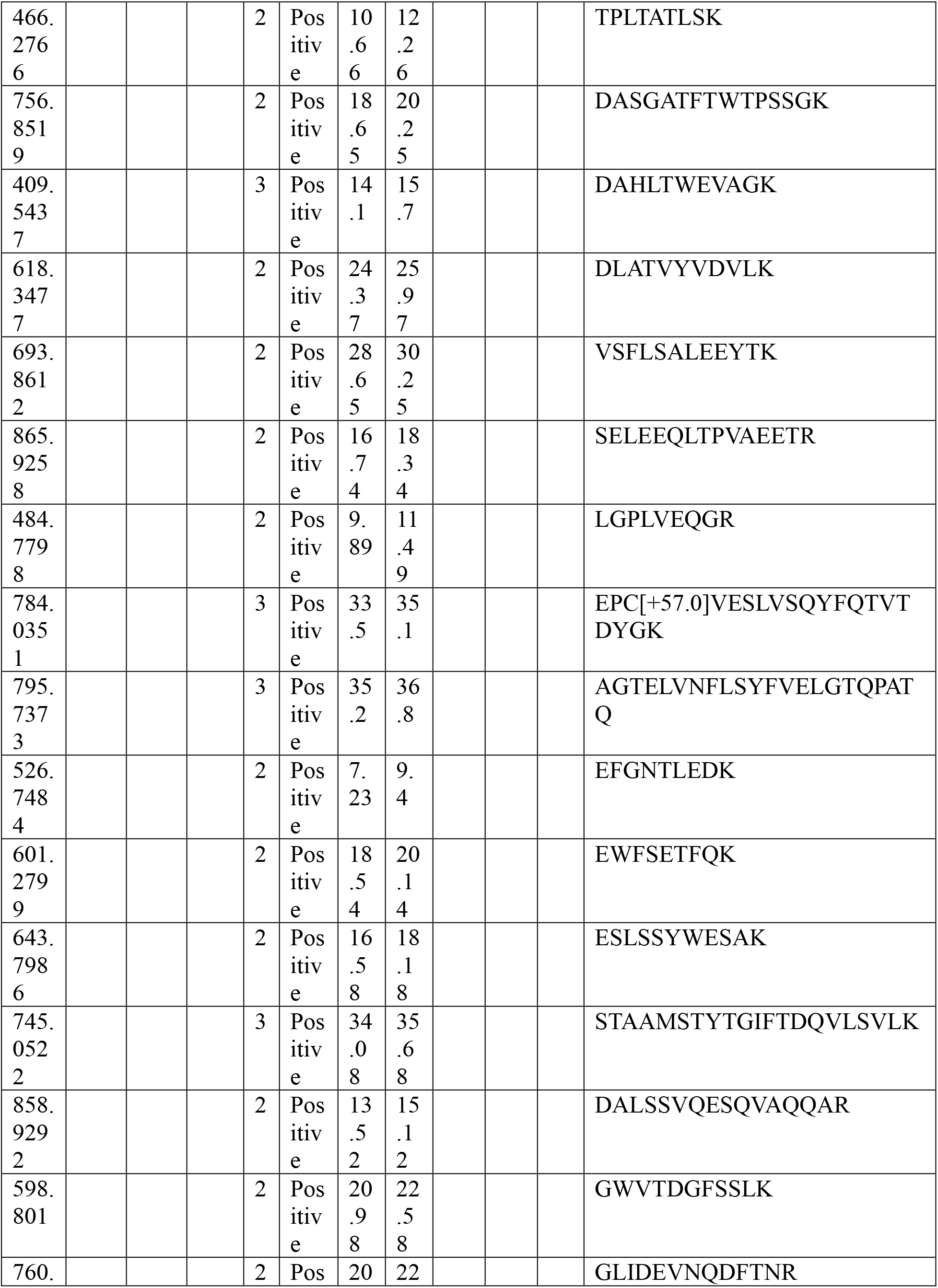
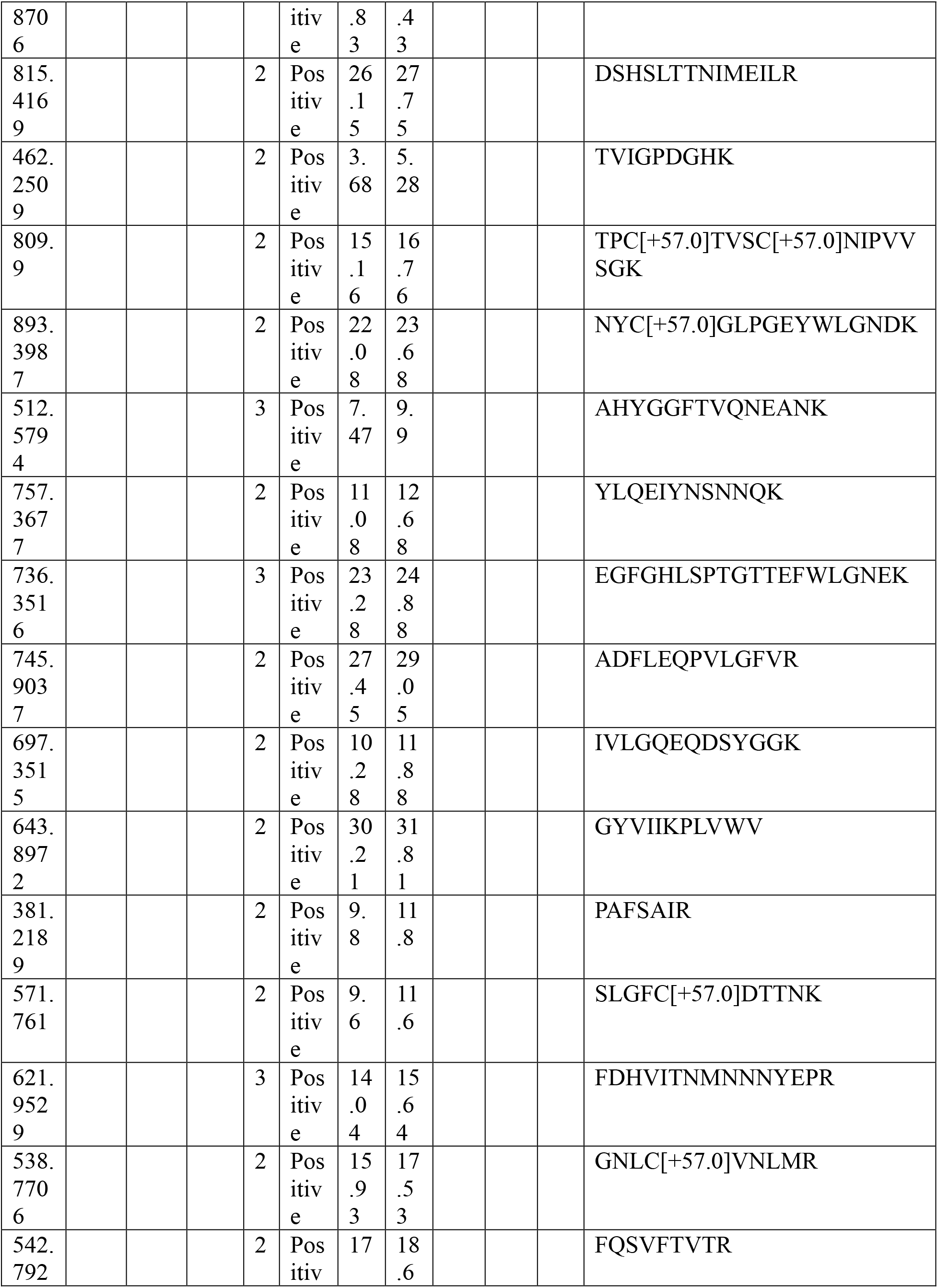
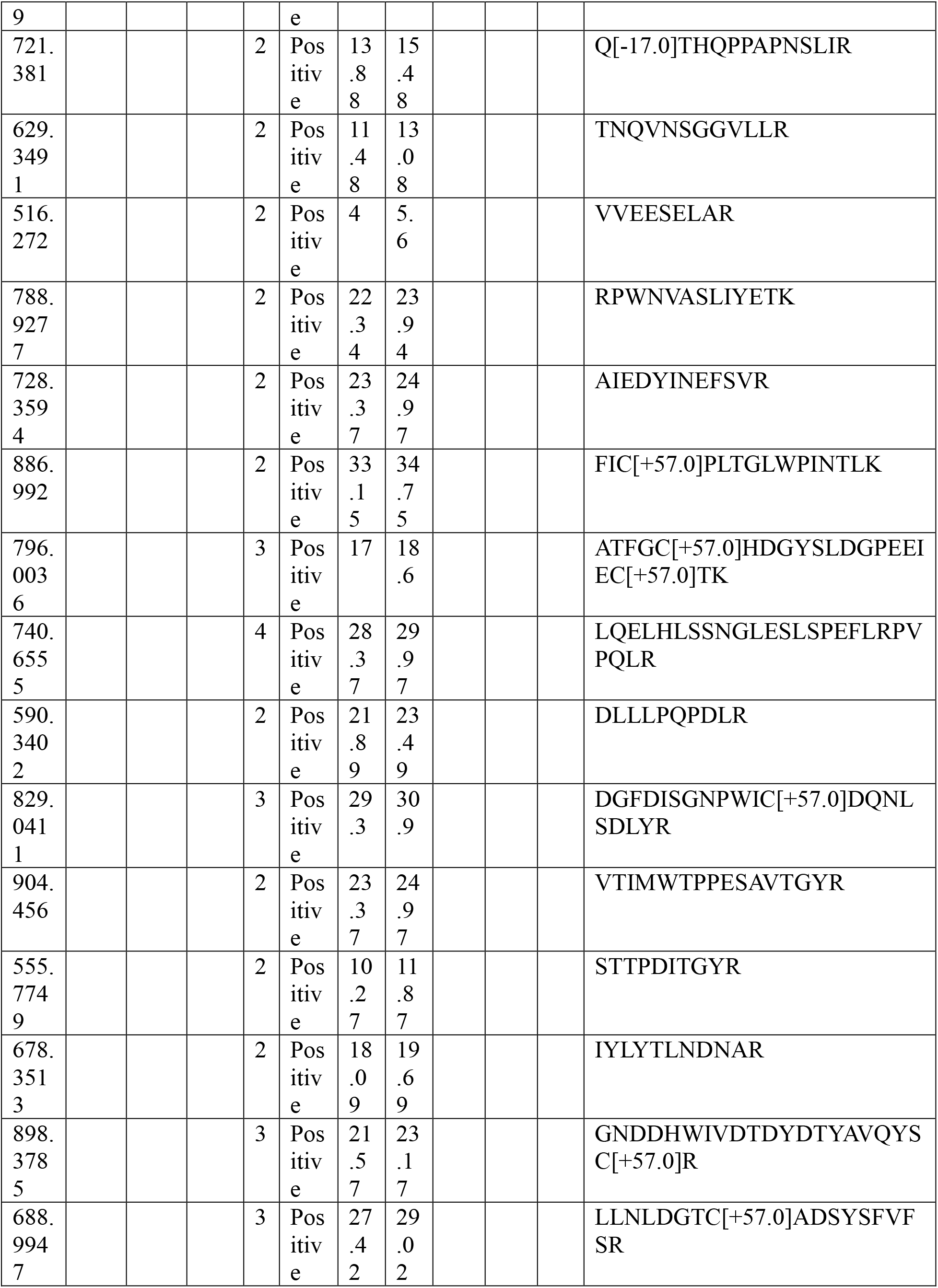
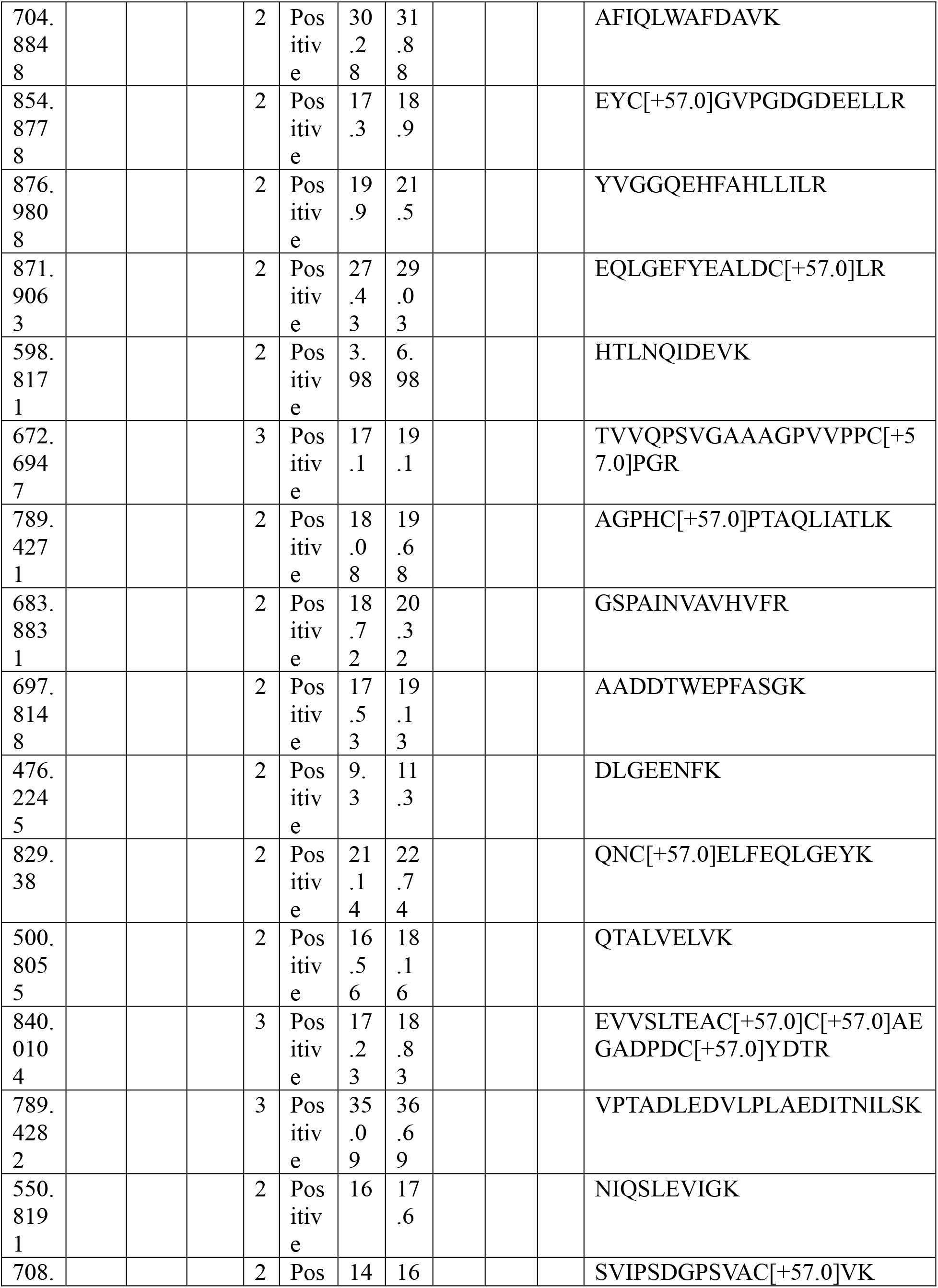
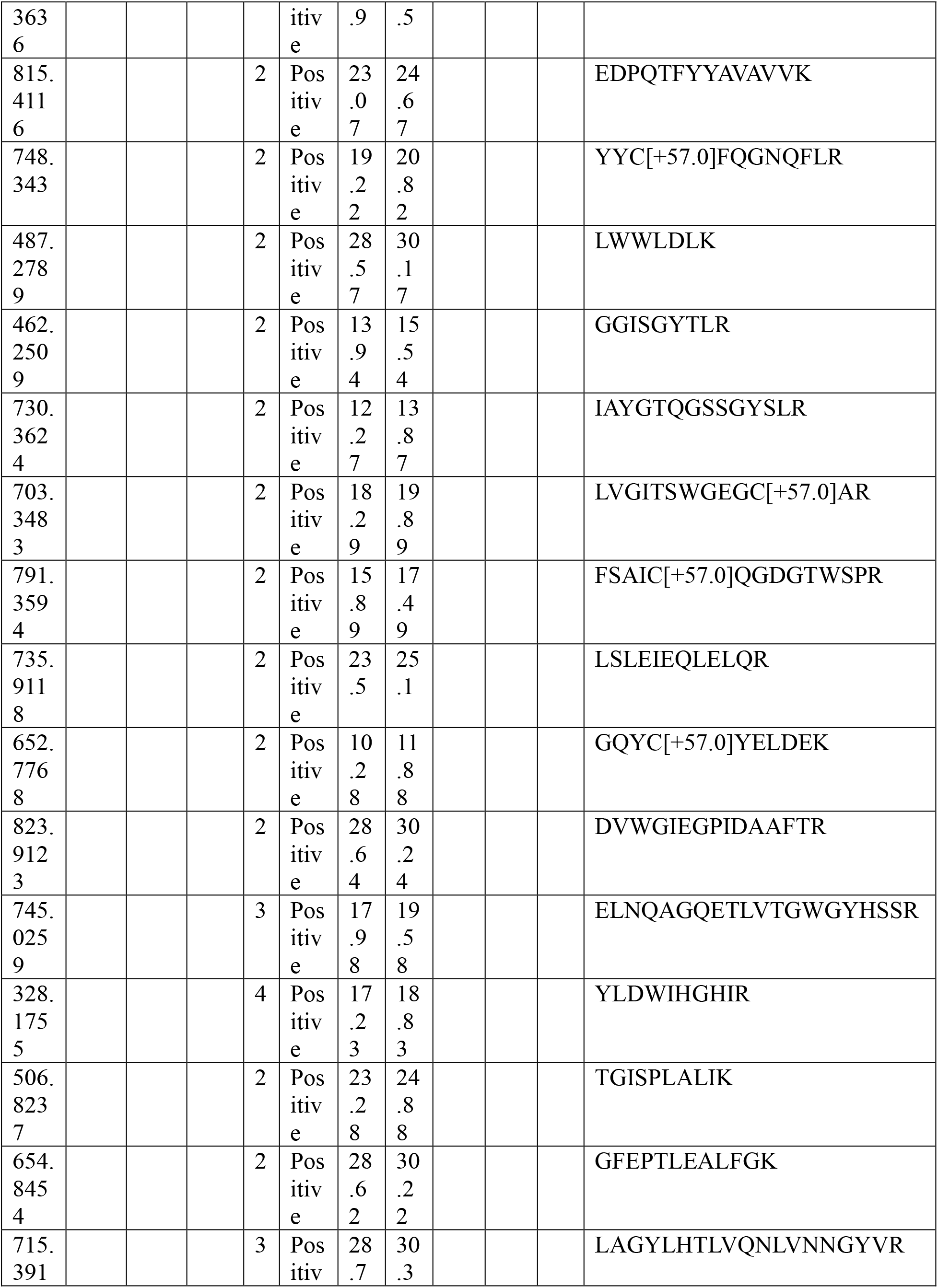
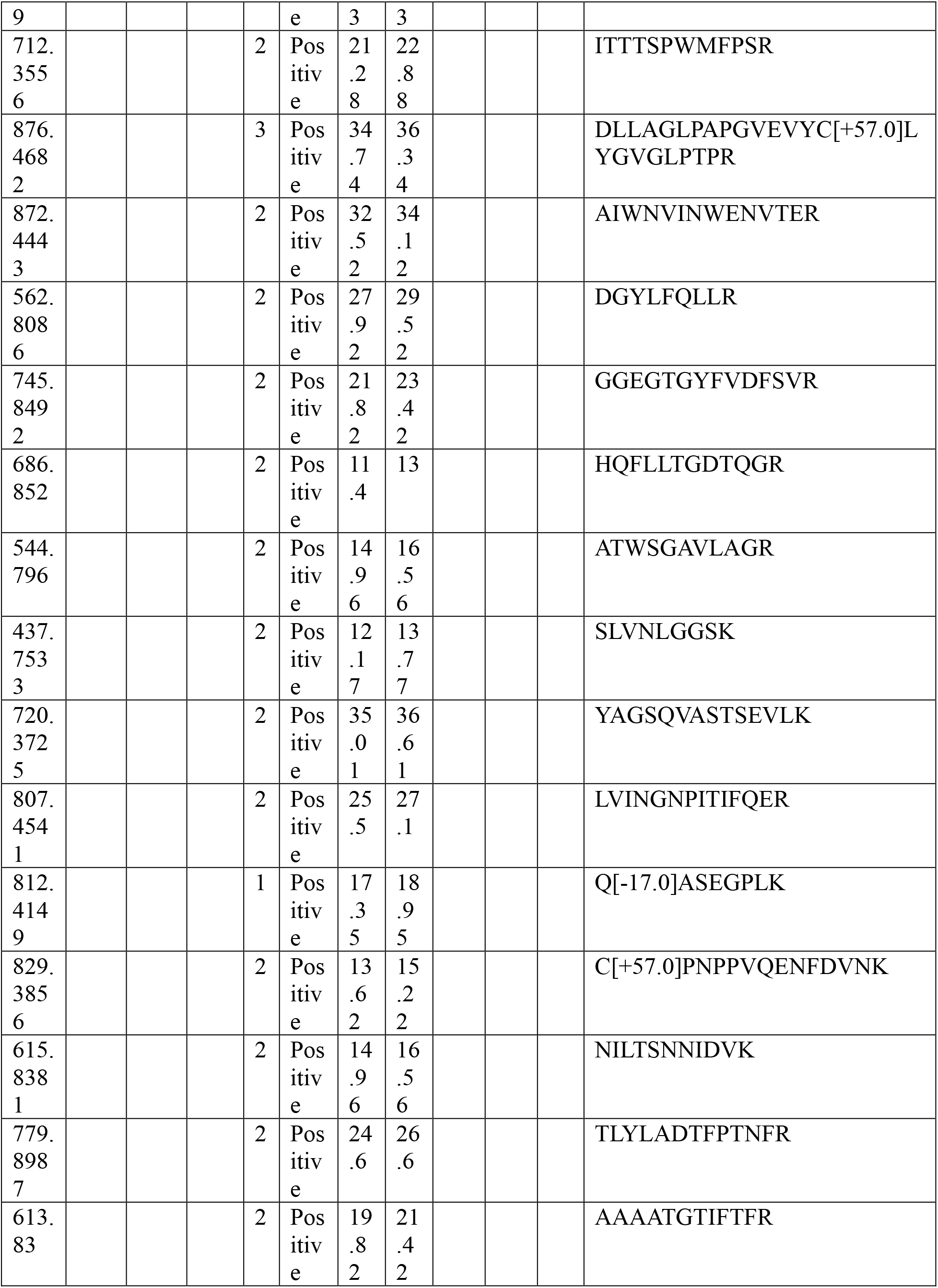
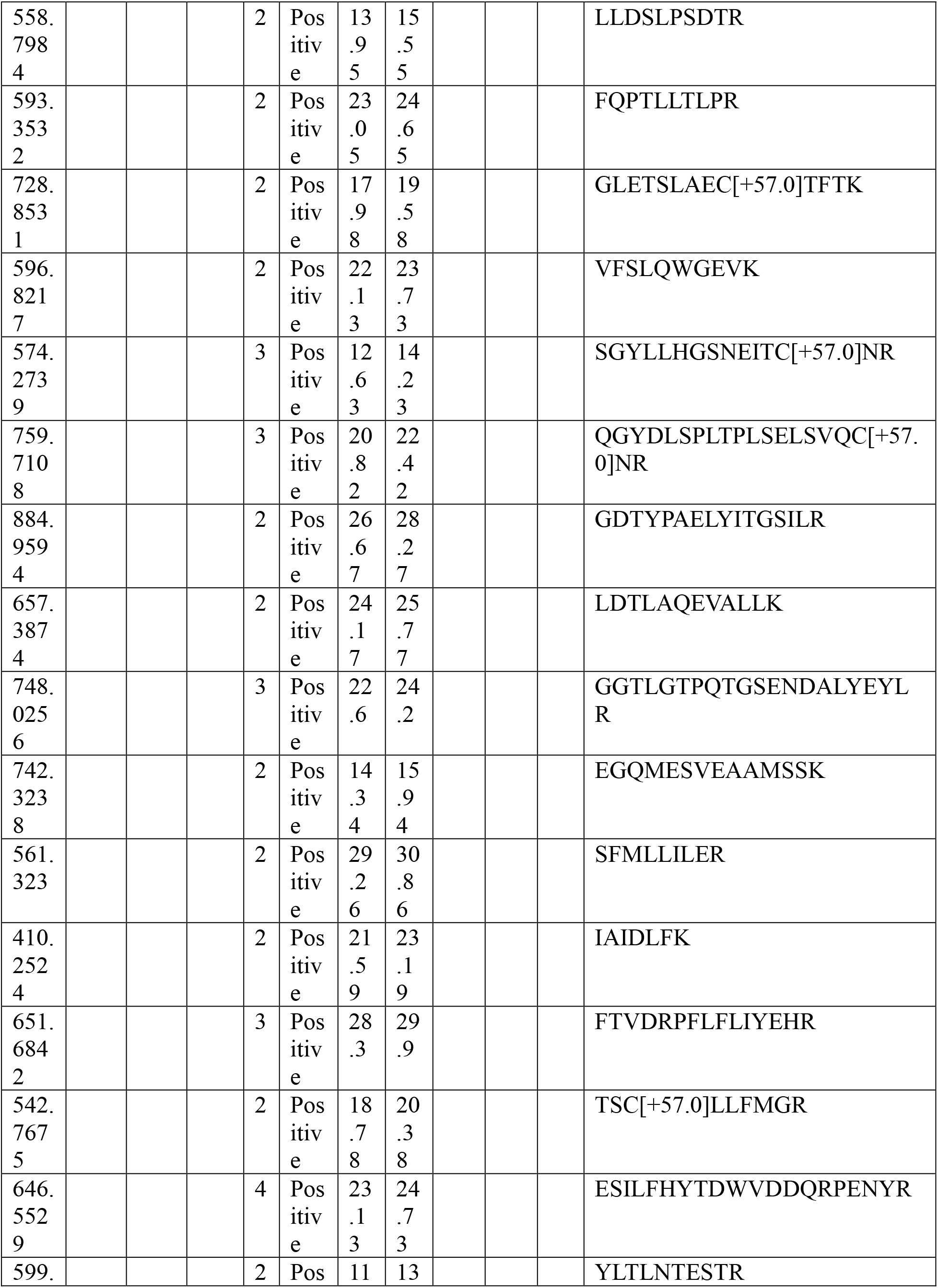
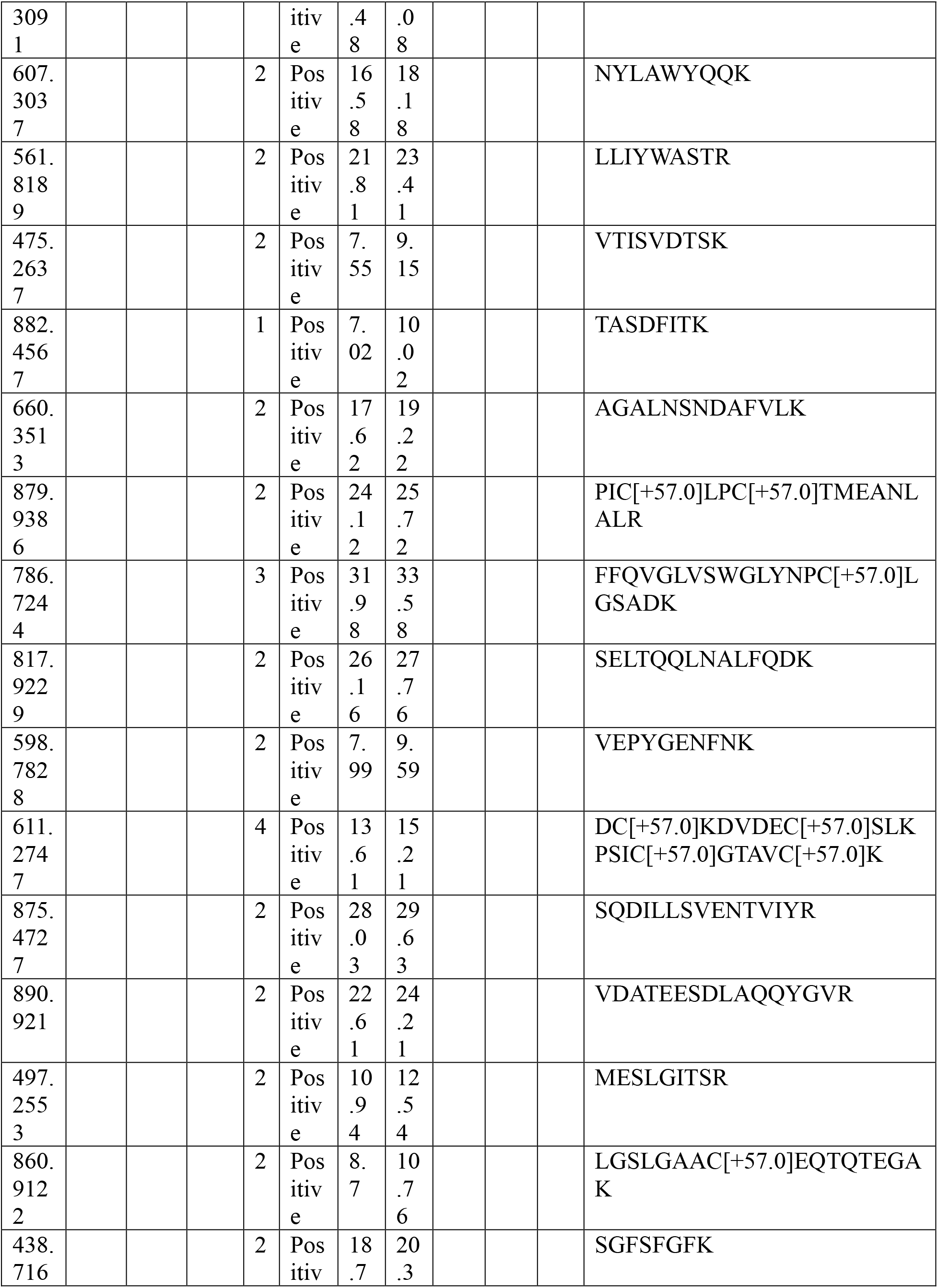
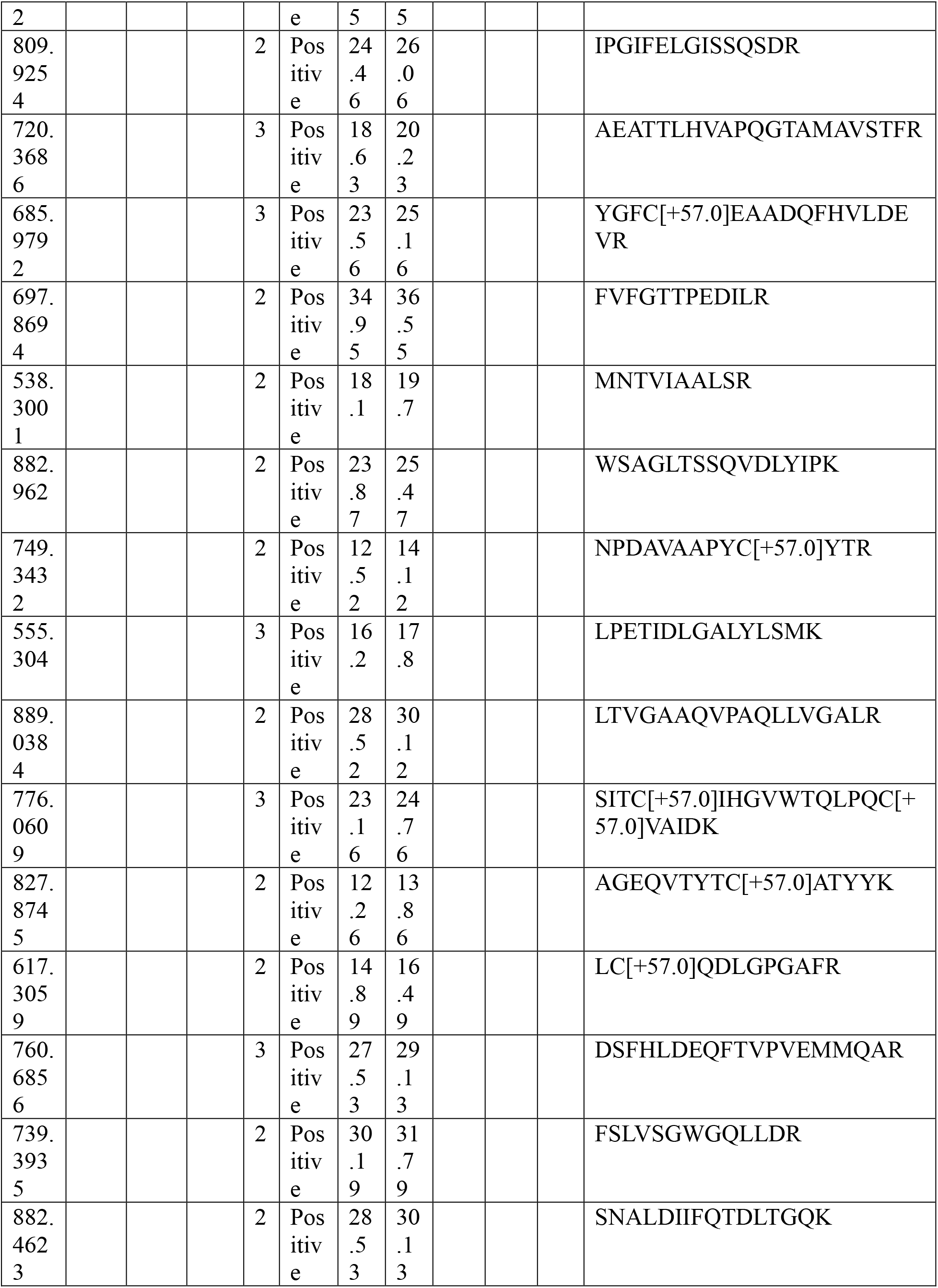
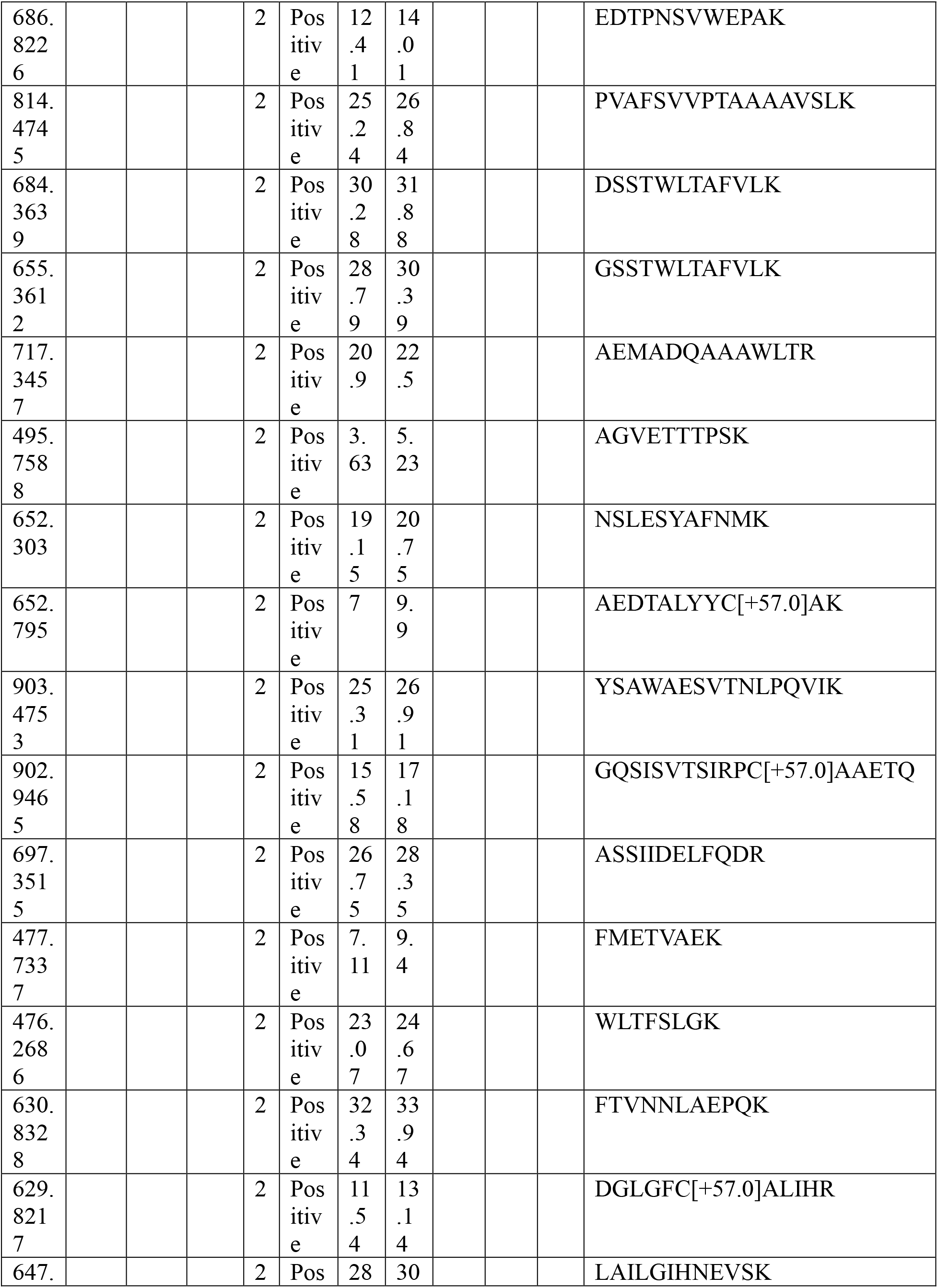
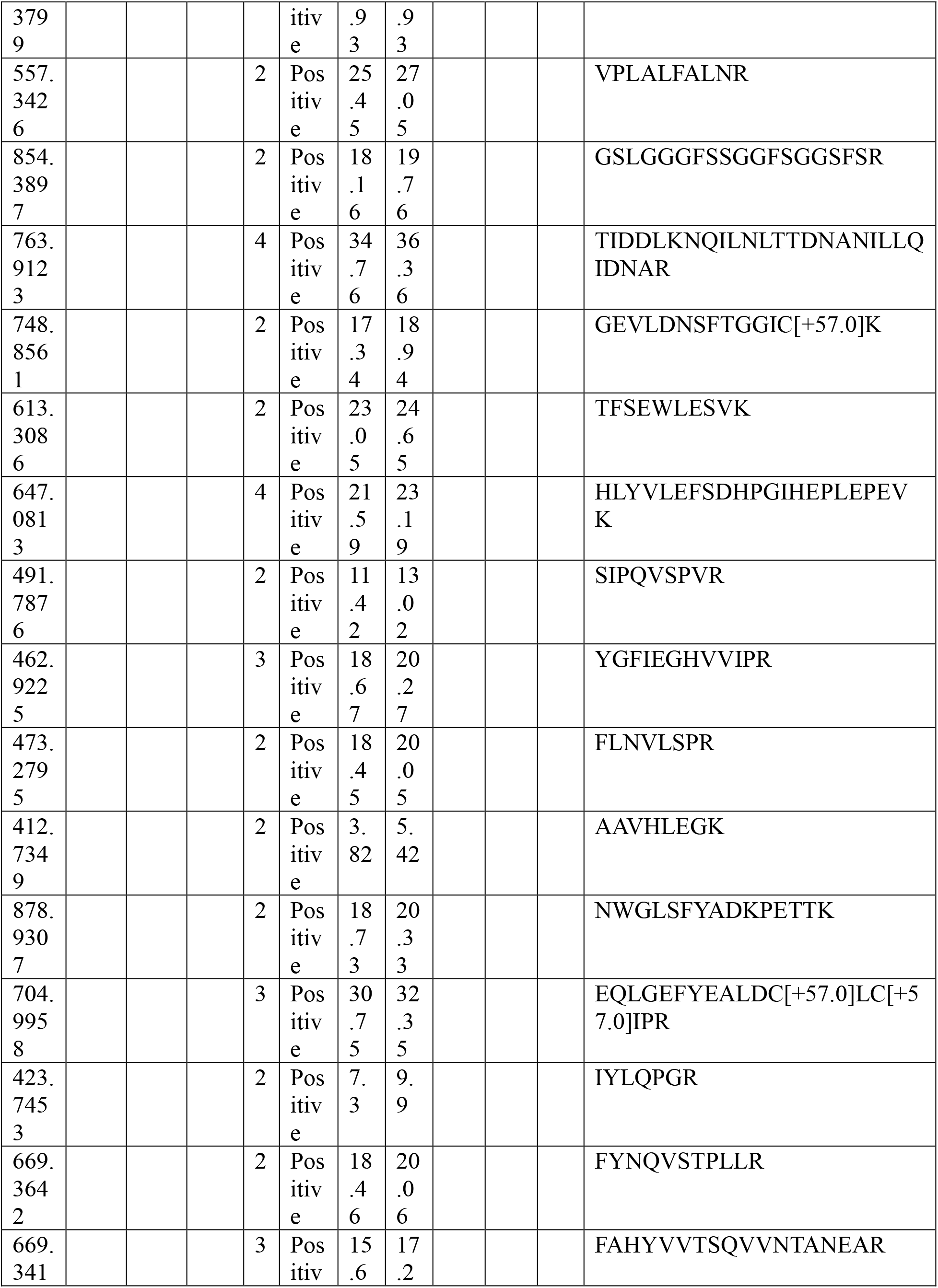
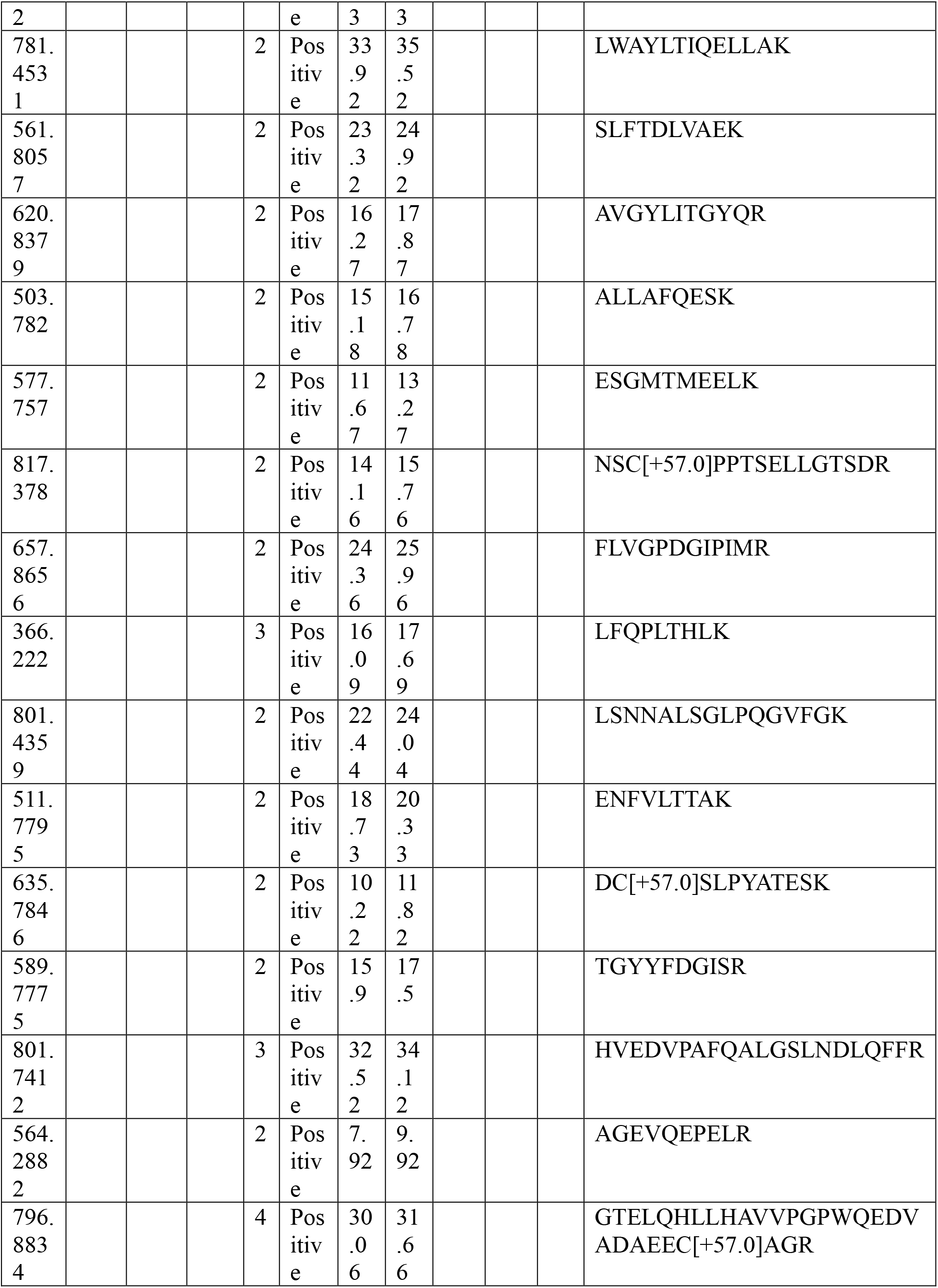
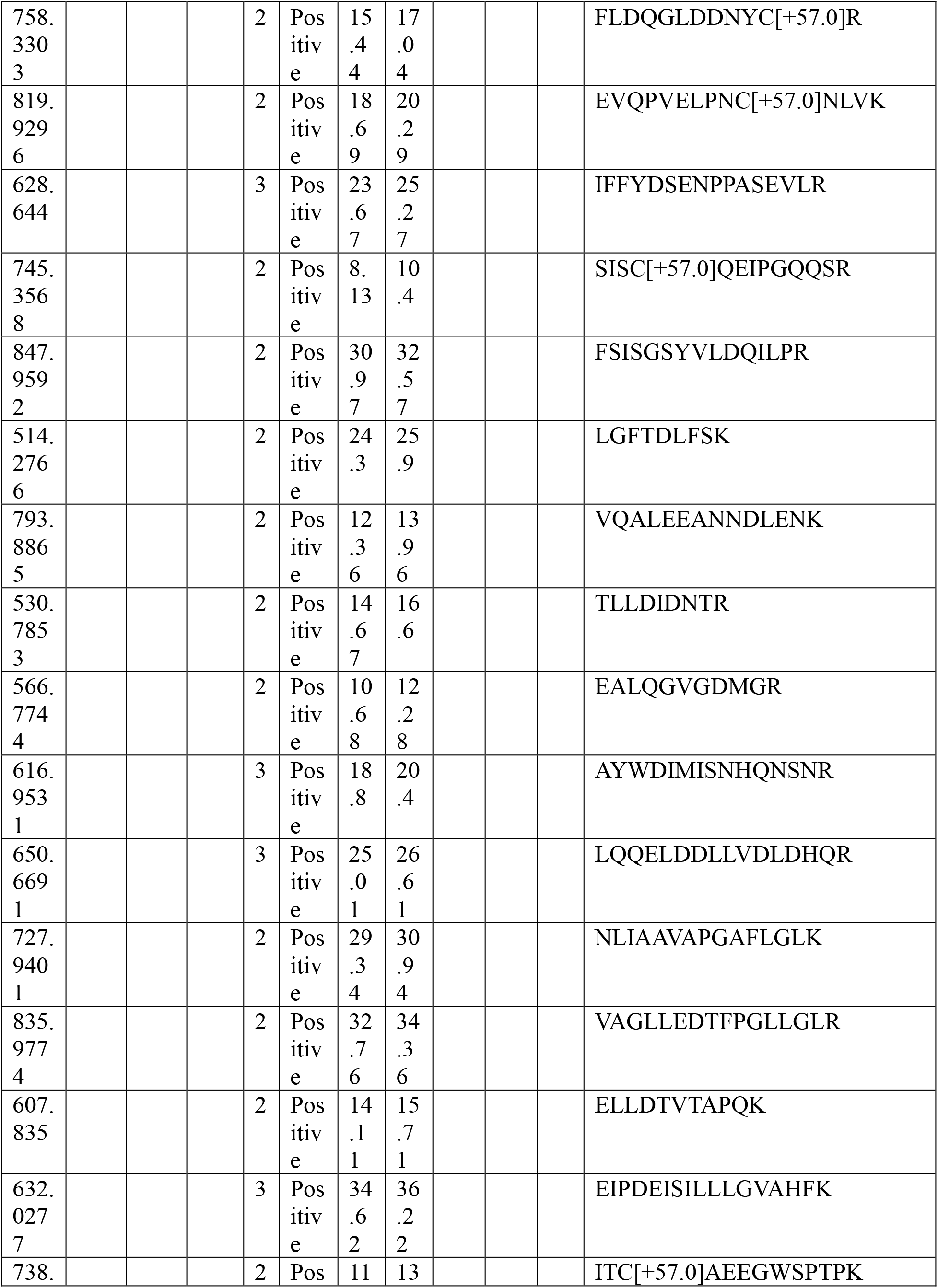
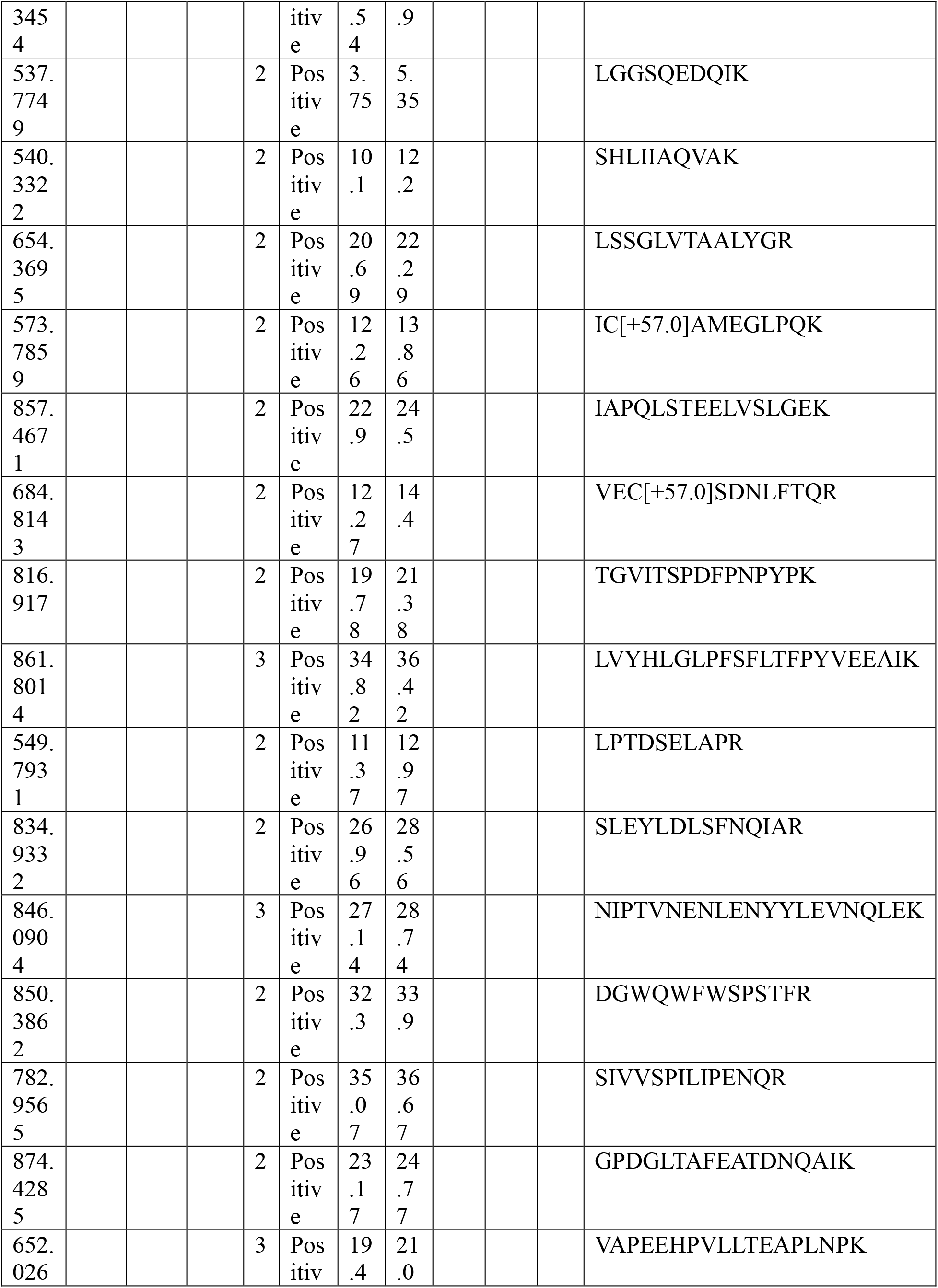
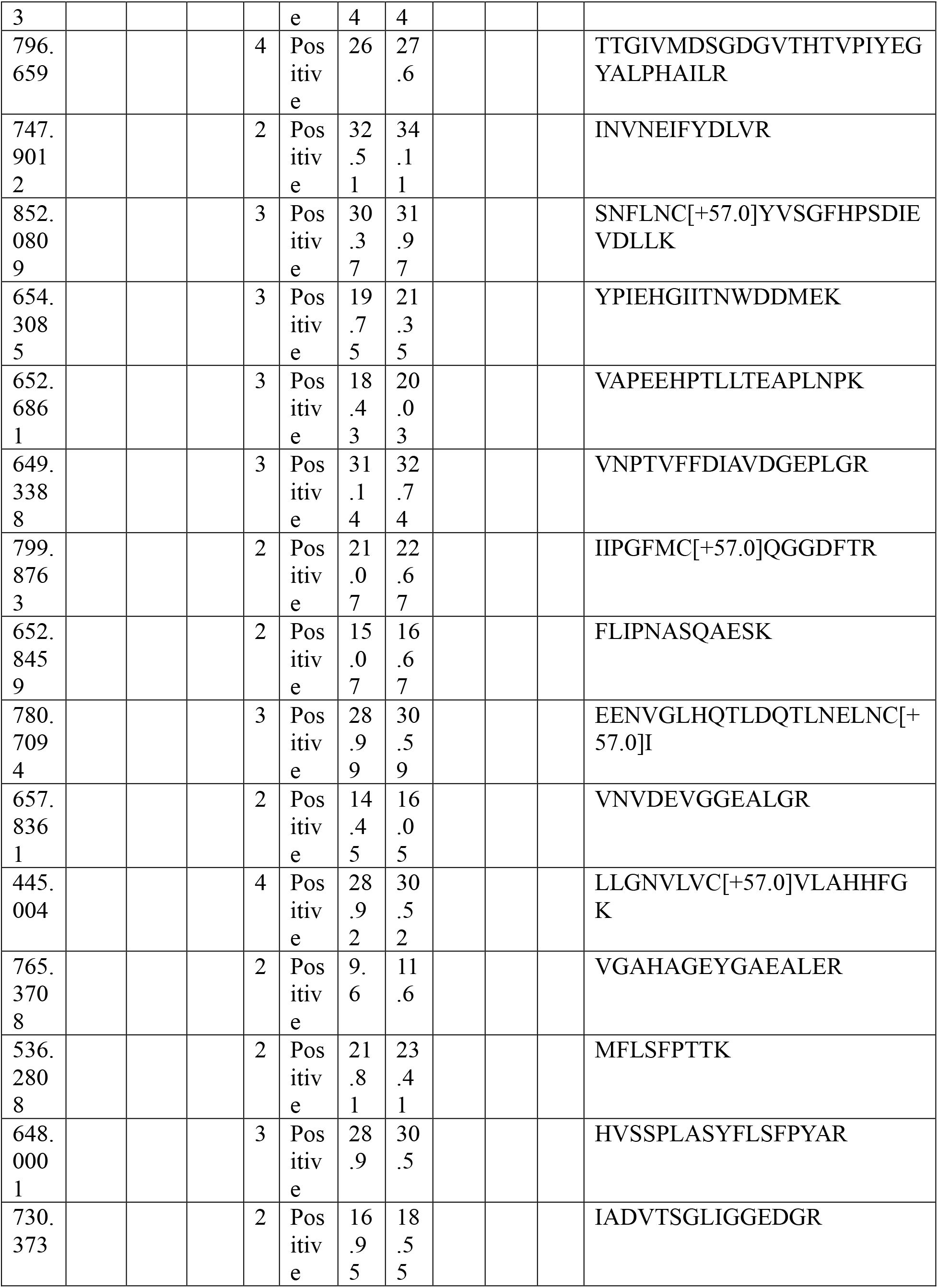
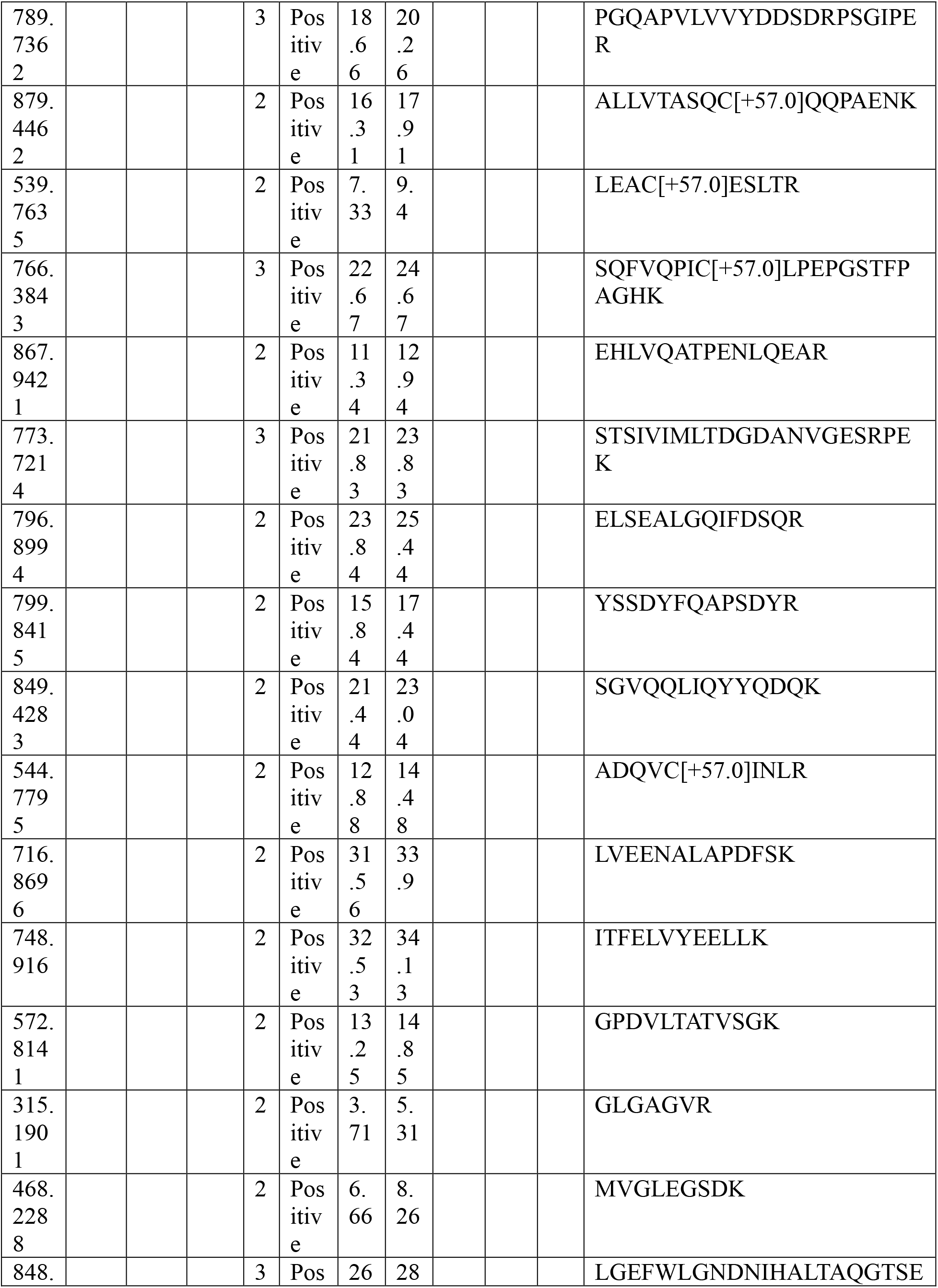
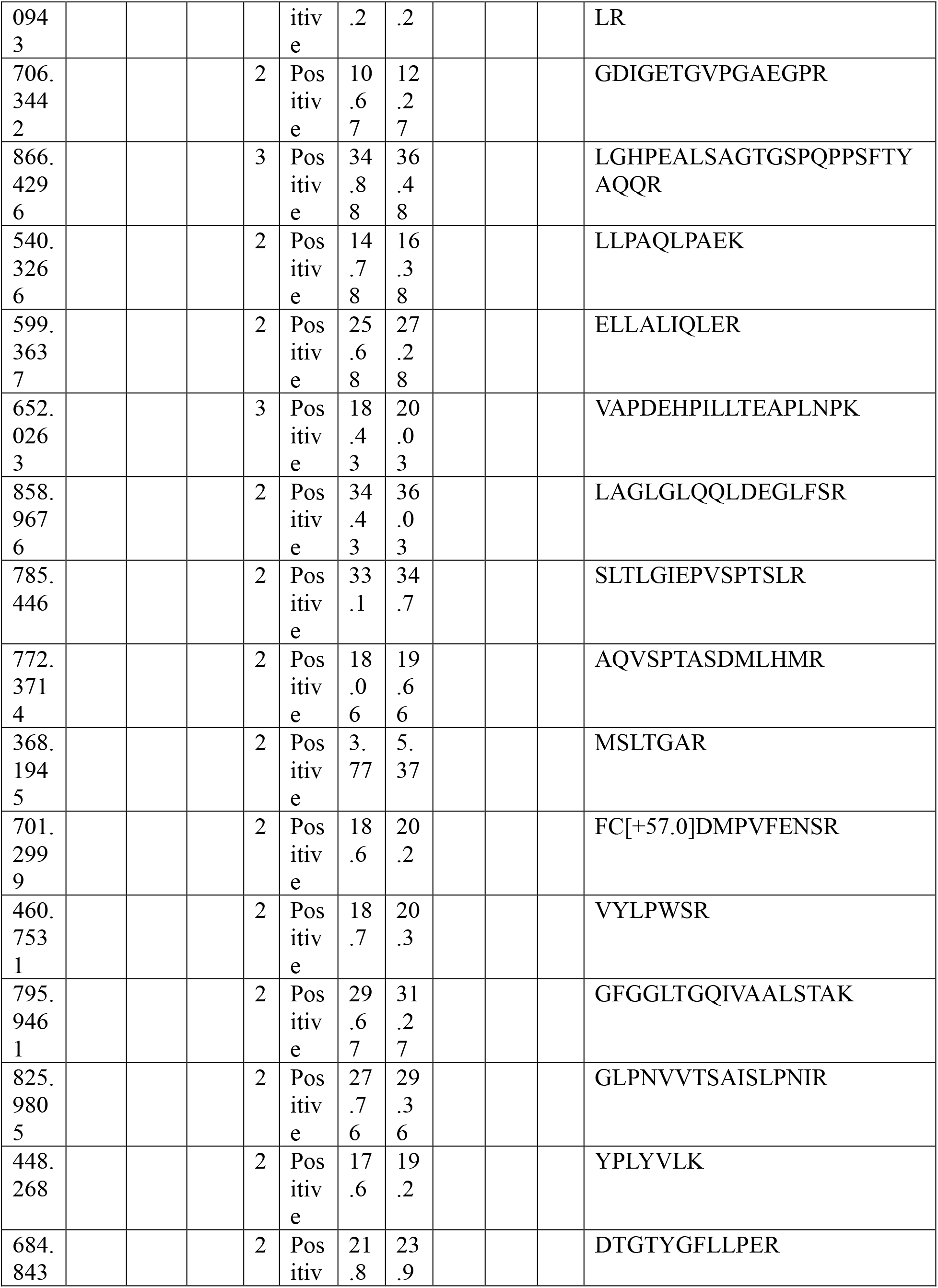
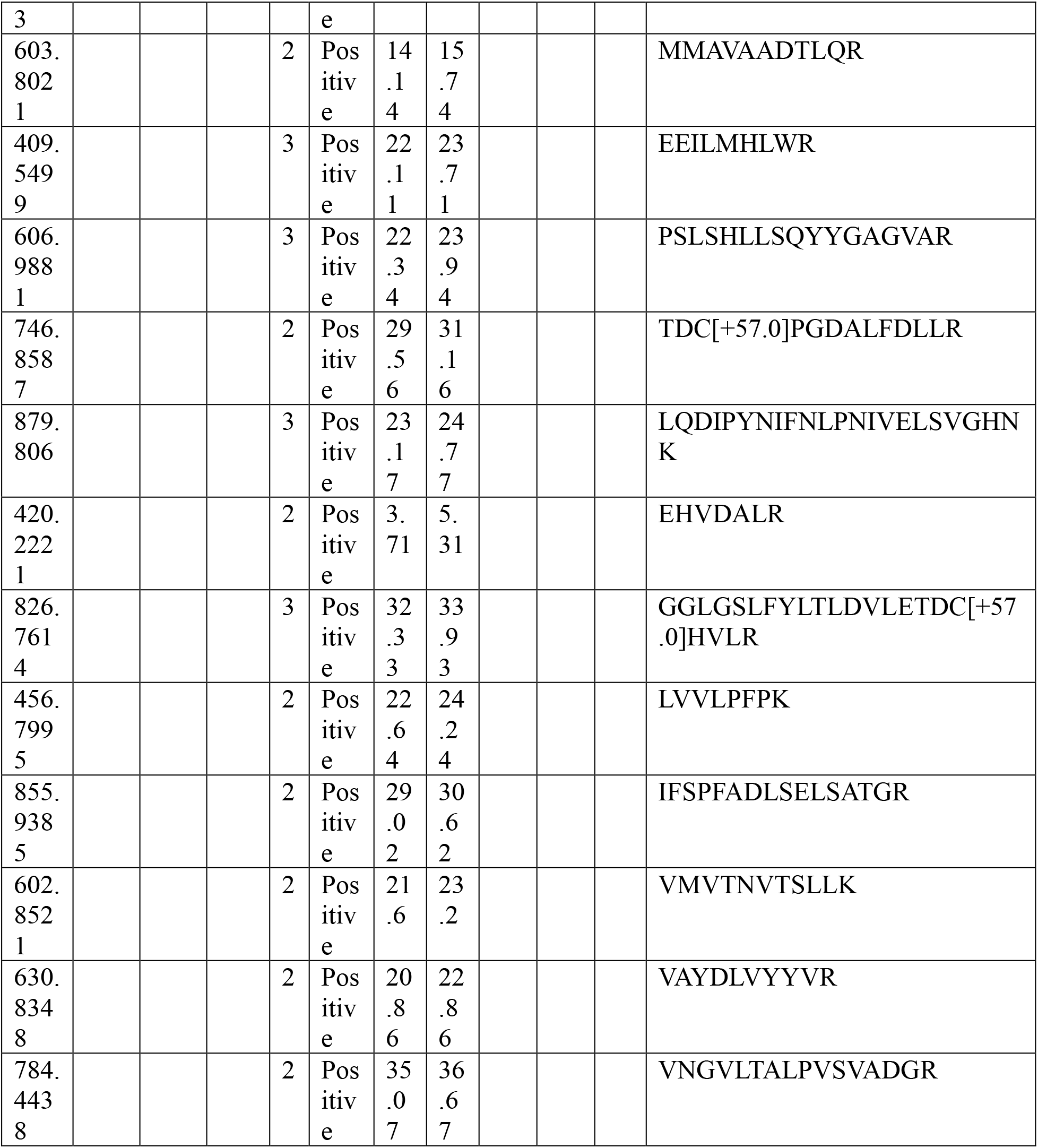
Inclusion list for targeted **proteomics** (PRM).

## Notes

### Competing Interest Statement

The authors have declared no competing interest.

### Funding Statement

National Institutes of Health grant R21NS116315 (CM)
National Institutes of Health grant P30ES013508 (CM)
Friedreich Ataxia Research Alliance (FARA) Postdoctoral Research Award (DW)

### Author Declarations

The study was approved by the Institutional Review Board (IRB) of the Children Hospital of Philadelphia (IRB Protocol 01 002609).

